# AI-driven fusion of neurological work-up for assessment of biological Alzheimer’s disease

**DOI:** 10.1101/2025.03.12.25323862

**Authors:** Varuna H. Jasodanand, Sahana S. Kowshik, Shreyas Puducheri, Michael F. Romano, Lingyi Xu, Rhoda Au, Vijaya B. Kolachalama

**Author notes:** Corresponding author: Vijaya B. Kolachalama, PhD.

## Abstract

Alzheimer’s disease (AD) diagnosis hinges on detecting amyloid beta (A*β*) plaques and neurofibrillary tau (*τ*) tangles. While amyloid PET imaging is now clinically approved, tau PET remains largely restricted to research settings. These imaging techniques, though valuable, are expensive and often difficult to access, limiting their widespread use in routine clinical practice. Here, we introduce a computational framework that leverages multimodal data from seven distinct cohorts comprising 12, 185 participants to estimate indi-vidual PET profiles, both global and regional, using more accessible data modalities, such as demographics, medical history, medication use, fluid measurements, functional and neuropsychological assessments, and structural MRIs. Our approach achieved an area under the receiver operating characteristic curve of 0.79 and 0.84 in classifying persons with positive A*β* and *τ* status, respectively. Model predictions were consistent with various biomarker and cognitive profiles, as well as with different degrees of protein abnormalities observed in post-mortem examinations. Furthermore, the regional volumes identified by the model as important aligned with the spatial distributions of the standardized uptake value ratio for regional *τ* labels. Our model offers a practical approach to identify potential candidates for newly approved anti-amyloid treatments and AD clinical trials for combined amyloid and tau therapies by utilizing standard neurological evaluation data.

## Introduction

Alzheimer’s disease (AD) is biologically defined by the progressive accumulation of amyloid beta (A*β*) plaques and neurofibrillary tau (*τ*) tangles ^1^. These proteinopathies develop years before symptom onset, presenting a window for early therapeutic interventions ^2^. The temporal progression of these biomarkers also facilitates biological staging of AD, guiding treatment strategies and timing ^3^. While amyloid positron emission tomography (PET) imaging is clinically approved for detecting A*β*, *τ* PET remains largely restricted to research settings ^4^. These imaging modalities provide critical insights into disease progression but are expensive and not widely accessible, limiting their routine clinical use compared to conventional modalities such as structural magnetic resonance imaging (MRI) and neurocognitive assessments. Cerebrospinal fluid (CSF) testing offers high sensitivity for amyloid detection but lacks the ability to stage disease progression, which tau PET imaging currently provides ^4^. PET imaging influences clinical decision-making ^5^ and remains integral to identifying candidates for disease-modifying therapies and clinical trials ^6–8^. However, its restricted accessibility in routine care settings underscores the need for cost-effective, scalable screening methods that preserve PET’s staging precision while overcoming logistical barriers.

The escalating costs associated with AD drug development underscore the necessity for precise disease staging. From 1995 to 2021, AD research and development incurred an estimated $42.5 billion expenditures, with a staggering 95% failure rate ^9^. A large portion of these costs stems from the screening process required to determine patient eligibility based on A*β* PET positivity status ^9^. However, emerging evidence suggests that *τ* pathology is more strongly linked with cognitive decline and disease progression ^10^. The TRAILBLAZER-ALZ 2 clinical trial demonstrated that Donanemab, an amyloid-lowering therapy, was most effective in patients with lower *τ* PET burden ^6^, highlighting the critical role of *τ* staging in determining therapeutic response. These findings highlight the urgent need for scalable approaches to estimate *τ* burden, bridging the gap between research and clinical practice to optimize patient selection for emerging AD therapies ^11,12^.

Emerging technologies and frameworks, including plasma biomarkers such as p-tau 217, offer potential for early AD detection ^13,14^. While these biomarkers can predict A*β* PET status with performance comparable to cerebrospinal fluid (CSF) analyses in certain cohorts ^15^, their ability to accurately predict tau PET status across diverse populations is less established ^14,16^. Further, these biomarkers lack the ability to capture the spatial distribution of tau pathology in the brain, which is essential for accurate biological assessment of AD ^4,17,18^. Variability due to non-neurological factors such as body mass index, cardiovascular and renal health can also affect their clinical efficacy ^19,20^. Finally, the generalizability and accuracy of cut-off points in racially and ethnically diverse samples have not yet been demonstrated, raising questions about their readiness to enter standard clinical workflows ^21^. Therefore, while promising, plasma biomarkers are not yet a standalone solution, and there is a need for integrated and accessible models capable of accurately pre-screening and stratifying patients based on their A*β* and *τ* status, as well as disease stage ^4,11^.

Machine learning (ML) models have shown promise in addressing some of the logistical challenges of PET scans by predicting A*β* or *τ* PET status using less invasive data such as demographics, MRIs and cognitive assessments ^22–30^. However, these models often face limitations, including development on relatively small cohorts, reliance on emerging plasma biomarkers, lack of external validation and adaptability to real-world settings where data availability varies. By leveraging standard-of-care data, there is an opportunity to develop a cost-effective pre-screening process that reliably estimates both amyloid and tau pathology, enabling broader access to advanced diagnostics and targeted treatments.

Here, we propose a transformer-based ML framework designed to integrate multimodal data and predict global A*β*, tau burden in a pre-defined meta-temporal region (meta-*τ*) encompassing medial and neocortical temporal regions ^31^, and regional tau PET statuses. By incorporating demographic information, medical history, neuropsychological assessments, genetic markers, neuroimaging and other relevant clinically obtained data, we sought to create a flexible computational framework that could be implemented in real-world settings, where complete feature sets are rarely available. Additionally, since tau pathology alone is not specific to AD ^32^, we aimed to output a combined prediction of both A*β* and *τ* accumulation to enhance specificity for AD. This approach not only fills the gaps in existing research by providing integrated models for concurrent A*β* on *τ* pathology prediction, but also facilitates a cost-effective participant selection process that can streamline clinical trials and research. Finally, by outputting probabilities that align with established biological stages, our modeling framework could concurrently identify and stage AD.

## Results

Our modeling framework was developed through training on a large, diverse dataset with multimodal features (Fig. 1 & Tables S1-S9), and rigorously tested on an external dataset (Table 1). We evaluated our framework’s alignment with PET-estimated A*β* and *τ* burden and biomarker profiles, and assessed its ability to capture the synergistic relationship between A*β* and *τ*. Additionally, we constructed a graph network using Shapley values of brain volumes for each regional tau label and validated the model’s regional tau predictions against tau PET SUVr values in the same regions. Finally, we compared the model predictions with postmortem findings, ensuring that the predicted probabilities reflected the severity of the underlying pathology.

**Figure 1:**
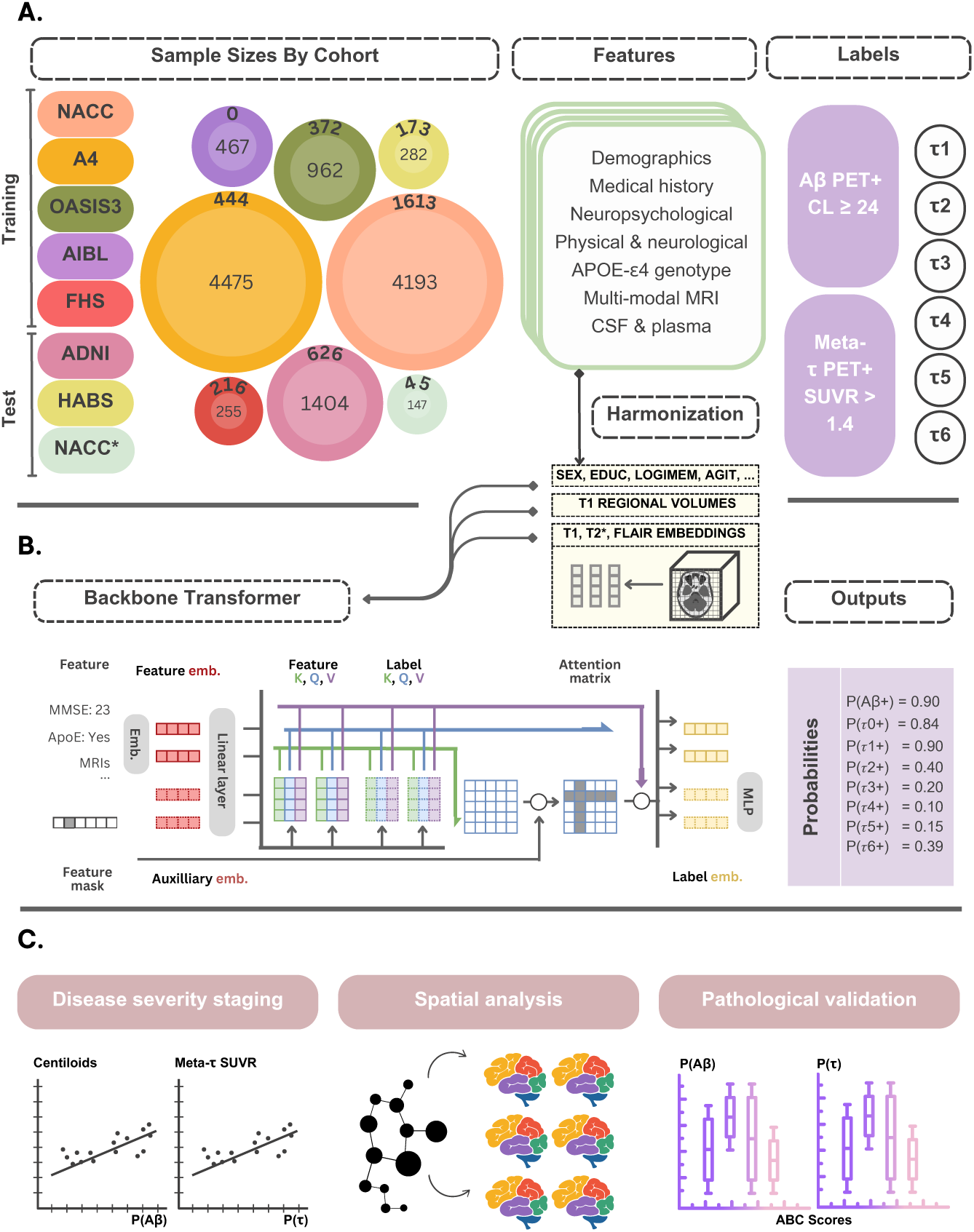
Data, model development and validation strategy. (a) Our model for assessing amyloid and tau status was developed using diverse data modalities, including individual-level demographics, health history, genetic information, neuropsychological testing, physical/neurological exams, and multi-sequence MRI scans. These data sources were aggregated from eight independent cohorts: NACC, A4, OASIS3, AIBL, FHS, ADNI and HABS. All features were harmonized to the UDS3 format and embeddings were extracted from multi-modal MRI scans. Inner concentric circles provide the sample size of the cases with A*β* PET data and outer circles denote the sample size with *τ* PET data. (b) Each feature was transformed into a set length vector through a modality-specific embedding approach before being input into the main transformer. A linear layer then linked the transformer to the output prediction layer. (c) The external ADNI and HABS datasets, as well as a held-out set of NACC* data, were selected to compare pathology-specific model predicted probabilities with functional and biological outcomes, as well as alignment with neuropathology grades. Shapley analysis was run on the regional *τ* model, and a graphical network analysis was performed to detect clusters of important brain regions using the Shapley values of the T1-weighted derived volumes. A similar community detection algorithm was run on the raw regional tau PET SUVrs and we compared communities derived from Shapley values with those derived from the regional SUVrs with statistical testing. The model architecture schematic in (b) was reproduced from our previous work ^34^.

**Table 1:**
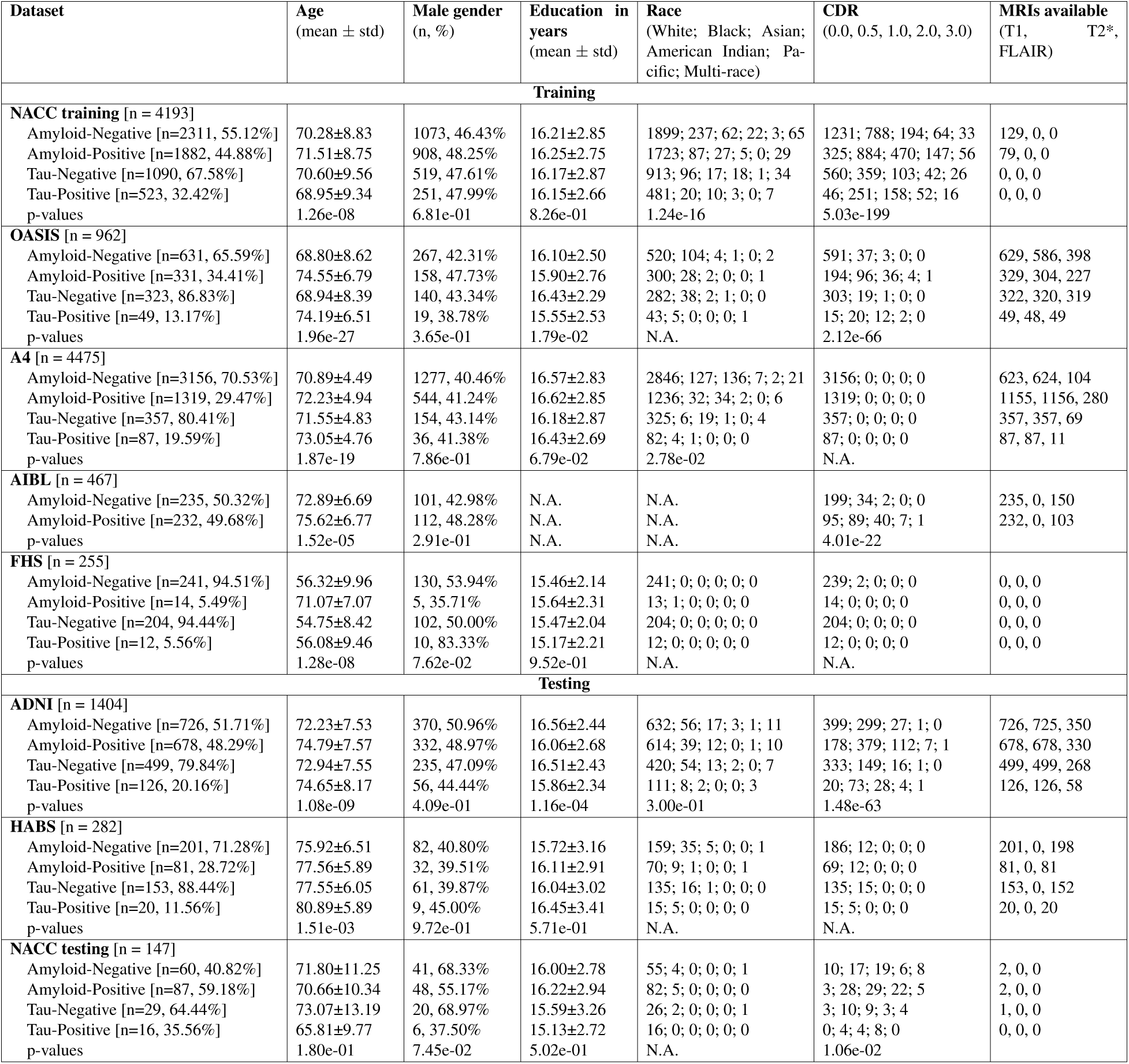
Study population. Summary of demographic and clinical attributes for the training and testing cohorts used in the study. Each cohort section details the mean age (with standard deviation), male participant count (with percentage), average years of education (with standard deviation), racial demographics (with counts for each racial category), clinical dementia rating (CDR) breakdown (with counts for each category), and the availability of MRI types (T1, T2*, and FLAIR). Statistical significance for between-group comparisons was assessed using one-way ANOVA for continuous variables (age, education), and chi-squared tests for categorical variables (gender, race, CDR), with p-values reported in scientific notation for each comparison. Statistical comparisons were not concluded for differences in MRI availability. The N.A. entries in the table signify scenarios when statistical tests could not be concluded and when data were not available.

### Model performance on A*β* and *τ* status

We first evaluated our model’s performance in predicting global A*β* and meta-*τ* status. The receiver operating characteristic (ROC) and precision-recall (PR) curves illustrate the model’s performance in predicting A*β* and tau positivity (Figs. 2a-b). The ROC curves show that the model achieves slightly higher sensitivity and specificity for *τ* (AUROC = 0.84) compared to A*β* (AUROC = 0.79). However, the PR curves indicate greater reliability in identifying true positive cases for A*β* (AUPR = 0.78) than for tau (AUPR = 0.60), despite the higher ROC for tau. This could be attributed to class imbalance or lower prevalence of *τ* positivity in the dataset, leading to a higher rate of false positives in *τ* predictions. Additional performance metrics are provided in Extended Table 1a. Supplementary Tables S10 and S11 detail the performance metrics for the internal validation set (NACC) and the combined ADNI-HABS external set, respectively. Notably, the ADNI dataset had 54% fewer features than the heldout NACC* test set, and the HABS dataset had 72% fewer features. Despite these constraints in feature availability, our model maintained robust performance, highlighting its flexibility and ability to handle incomplete feature sets without significant loss of accuracy. In Extended Data Fig. 1, we reported AUROC and AUPR metrics stratified by age, gender and race. The consistent performance across these subgroups indicates that our model is potentially applicable to diverse populations.

**Figure 2:**
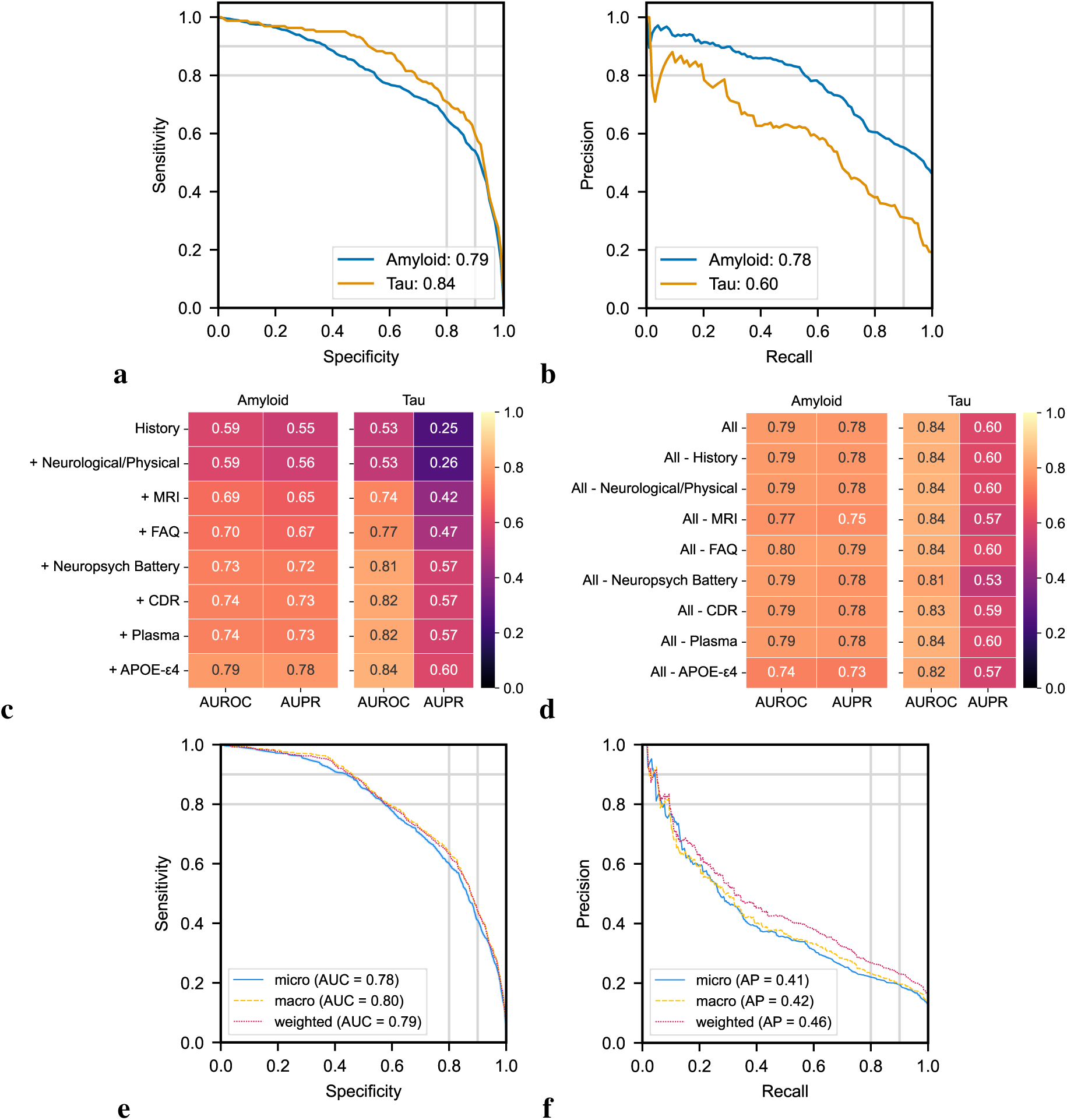
Model performance in predicting amyloid and tau positivity. (a, b) Receiver operating characteristic (ROC) and precision-recall (PR) curves for A*β* and meta-*τ* predictions are shown. The area under the ROC curve (AUC) and the area under the PR curve (AUPR) values for A*β* and meta-*τ* are displayed in the legends, respectively. (c) Heatmap presenting the ROC and PR values for A*β* and meta-*τ* predictions using various combinations of clinical features, starting with person-level history alone and incrementally adding features such as MRI, neuropsychological battery, and plasma data. (d) Heatmap displaying the AUC and AUPR values for A*β* and meta-*τ* predictions when specific feature sets are removed from the full model. Each row represents the model performance after excluding one feature set, showing how the absence of that data impacts prediction accuracy. (e, f) ROC and PR curves showing micro-average, macro-average, and weighted-average calculations based on the regional *τ* labels. A portion of the NACC dataset used for internal testing, along with data from the ADNI and HABS cohorts for external validation, contributed to generating these results.

To assess the impact of different types of clinical features on model performance, we evaluated the model’s predictions for A*β* and meta-temporal tau (meta-*τ*) status by successively adding different feature groups. Following the typical order of assessments in neurological work-up protocols for cognitive impairment, our analyses aimed to identify incremental gains, if any, when each new test is added to the work-up process (Figs. 2c-d). The plasma biomarker available at testing, the A*β* 42/40 ratio, and the APOE-*ɛ*4 tests are included last due to their relatively limited availability in clinical settings. For A*β* prediction, the AUROC improved from 0.59 with only person-level history to 0.79 when all features were included, with the AUPR values increasing in parallel from 0.55 to 0.78. Tau prediction models showed a comparable increase in AU-ROC from 0.53 with only patient history to 0.84 with all features. Notably, the addition of MRI data led to a substantial improvement in meta-*τ* AUROC from 0.53 to 0.74. Subsequent additions of neuropsychological battery scores provided additional improvements, highlighting that the integration of multiple modalities of data leads to better overall performance.

To evaluate our model’s robustness to the absence of specific feature sets, we systematically removed groups of features from the full model. For A*β* predictions, removing any single feature set had minimal impact on AUROC values, which remained between 0.74 and 0.80. This highlights the strength of our random feature masking strategy, which allowed the model to make meaningful predictions even in the absence of certain data types. Similarly, *τ* predictions were robust across feature exclusions, with the removal of the neuropsychological battery resulting in the most significant drop in AUPR to 0.53. While our modeling strategy afforded the flexibility in achieving high accuracy despite the absence of certain feature sets, the importance of neuropsychological testing is underscored by the sensitivity of *τ* AUPR values to the removal of these features. The results of our Shapley analysis (Extended Data Fig. 2a-b) provide additional support for this interpretation, with neuropsychological testing, neuroimaging and APOE-*ɛ*4 status having, on average, the greatest impact on model output.

We quantified our model’s performance on regional *τ* predictions and found that it achieved a macro-average AUROC and AUPR of 0.80 and 0.42, respectively (Fig. 2e-f). Individual AUROC scores ranged from 0.71 to 0.84, indicating robust discriminative ability across different regions of interest (ROIs). The medial temporal *τ* label achieved the highest AUPR of 0.60, suggesting that the model is particularly effective in identifying true positive cases in this critical region (Extended Table 1b). These results suggest that our transformer-based model effectively predicts regional tau accumulation, particularly excelling in the medial and lateral temporal regions, where the combined AUROC and AUPR values were the highest.

We conducted a comparative analysis of our transformer-based model against CatBoost, a robust machine learning approach, to evaluate performance in predicting A*β* and *τ* pathology. For this purpose, we tested our model without MRI embeddings, with the results detailed in Table S12. On the combined test set from ADNI, HABS, and NACC*, CatBoost achieved an AUROC of 0.81 for A*β* predictions and 0.83 for *τ* predictions. The corresponding AUPR values were 0.79 for A*β* and 0.53 for *τ*. In comparison, our model demonstrated slightly lower AUROC for A*β* predictions (0.79 vs. 0.81) but superior AUPR for *τ* predictions (0.60 vs. 0.53), indicating more effective identification of true positive *τ* cases. Additionally, CatBoost’s balanced accuracy for A*β* prediction stood at 0.64, while ours was 0.68, indicating a more effective balance between sensitivity and specificity in our model. Further performance metrics for CatBoost are provided in Table S13a. To deepen our analysis, we incrementally added features from clinical assessments in the order typically collected during neurological work-ups to the CatBoost model. This step-by-step addition is visualized in Extended Data Fig. 3, contrasting the performance of our model without MRI embeddings (panel a) to that of CatBoost (panel b). Although CatBoost initially shows higher AUROC and AUPR upon integrating medical history and neurological/physical examination data, our model surpasses these metrics upon adding brain regional volumes, functional assessments, and neuropsychological tests. When MRI embeddings are incorporated into our model (Fig.2c), it achieves an AUROC comparable to CatBoost’s upon the addition of CDR scores and plasma A*β* 42/40 ratios, with a marginally better AUPR. Overall, our transformer-based architecture, with its attention mechanism and random feature masking, provides an endto-end framework that flexibly handles multi-modal inputs and performs effectively on imbalanced datasets. This is especially evident in its superior performance for meta-*τ* and regional *τ* predictions, where CatBoost exhibits a macro-average AUROC and AUPR of 0.77 and 0.38, respectively (Extended Data Fig.3, Fig.2c, and Supplementary Tables S12, S13).

### Validation with biological outcomes

Even though our model was trained on binary classifications, we aimed to assess its alignment with PET-based gradients of A*β* and *τ* accumulation Fig. 3. As an additional step towards facilitating clinical interpretability of our model outputs, we visualized how well the model’s predictions aligned with a commonly used clinical endpoint in AD trials, the Alzheimer’s Disease Assessment Scale-Cognitive Subscale (ADAS-Cog_13_ or ADAS13). We observed a positive correlation between P(A*β*) and centiloid values (Pearson’s r = 0.58, *p <* 0.0001; Fig. 3a), indicating that higher predicted A*β* levels are associated with increased A*β* plaque deposition, as confirmed by centiloid measurements. This relationship aligned with more severe cognitive impairment, evidenced by higher scores on the ADAS13. Similarly, we found a positive correlation between P(*τ*) and the log of meta-*τ* SUVr (Pearson’s r = 0.59, *p <* 0.0001; Fig. 3b), suggesting that higher model-predicted tau levels correlated with greater tau PET estimated pathology. An associated increase in ADAS-Cog_13_ was again visible, indicating more pronounced cognitive impairment at higher P(*τ*) values (Supplementary Table S14). We ran a similar analysis comparing the regional *τ* probabilities to the log of the corresponding regional *τ* SUVr values and found the strongest alignment for the medial temporal (Pearson’s r = 0.56, *p <* 0.0001, Extended Data Fig. 4a) and lateral temporal predictions (Pearson’s r = 0.52, *p <* 0.0001, Extended Data Fig. 4b). Further statistical results are reported in Supplementary Table S15.

**Figure 3:**
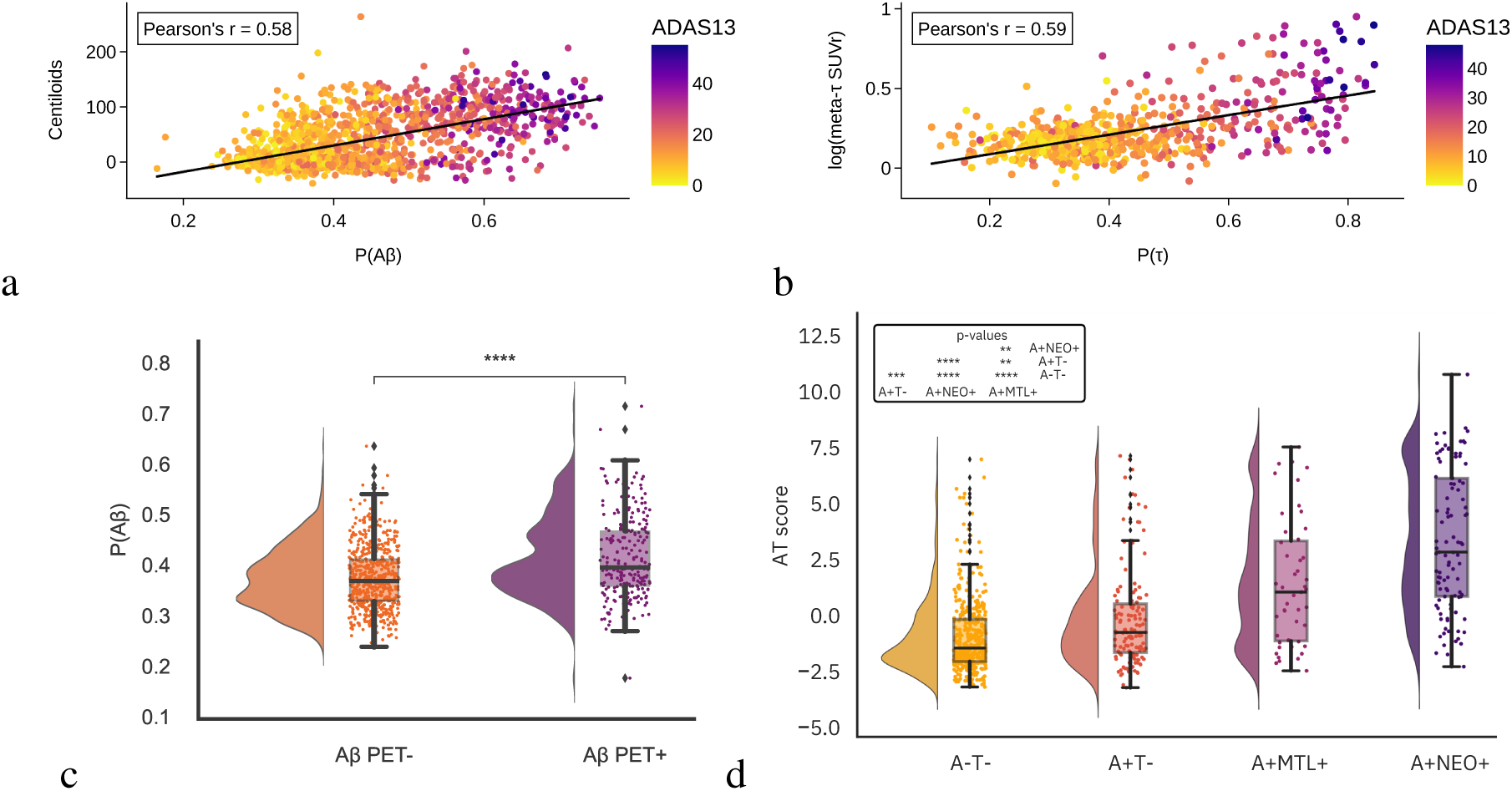
Model alignment with biological outcomes. (a) The bubble plot illustrates the model-predicted probabilities of amyloid PET positivity, P(A*β*), over centiloid values. The Pearson’s correlation coefficient was used to assess the strength of the relationship between the model’s probabilities and centiloids (*n* = 1392*, r* = 0.58*, p* = 4.04 *×* 10*^−^*^124^). The color scale indicates participants’ scores on the ADAS-Cog 13 task, a clinical tool for staging AD symptoms that was not provided as input to the model. (b) Model-predicted probabilities of tau PET positivity, P(*τ*), are shown over log-transformed SUVr values in the meta-temporal region. The Pearson’s correlation test was used to assess the relationship between the model probabilities and the log of meta-*τ* SUVr values (*n* = 619*, r* = 0.59*, p* = 2.35 *×* 10*^−^*^58^). Similarly, each data point is colored by ADAS-Cog 13 scores. Detailed statistical results can be found in Tables S14. (c) In a subset of cognitively unimpaired cases, we compared P(A*β*) between true A*β* PET negative (*n* = 590) and A*β* positive (*n* = 245) groups. A one-sided Mann-Whitney test indicated that model predicted probabilities were significantly lower for A*β* PET negative cases (*U* = 50727*, p* = 5.60 *×* 10*^−^*^12^). (d) The rainclouds plot illustrates the relationship between a composite score of model-predicted probabilities for A*β* and regional *τ* positivity against PET-defined disease stages. A Kruskal-Wallis H test, followed by post hoc Dunn’s testing with the Holm-Bonferonni correction revealed significant differences among cases in the A-T-(*n* = 411), A+T-(*n* = 139), A+MTL+ (*n* = 47), and A+NEO+ (*n* = 101) groups (*H* = 180.73*, p* = 6.15 *×* 10*^−^*^39^). Pair-wise post hoc results are provided in Supplementary Table S16. Cases from the ADNI cohort were used to generate the results shown in panels a-b and cases from both ADNI and HABS were used to generate results in panels c-d.

We sought to evaluate our model’s sensitivity for detecting A*β* positivity in preclinical AD by comparing P(A*β*) between A*β* PET-negative (*n* = 590) and A*β* PET-positive (*n* = 245) cognitively unimpaired individuals from the ADNI and HABS cohorts. A Mann-Whitney U test revealed significantly lower P(A*β*) values in A*β* PET-negative cases compared to PET-positive cases (*U* = 50727*, p* = 5.60 × 10*^−^*^12^, Fig. 3c), demonstrating the model’s ability to distinguish between amyloid status groups even in the absence of cognitive symptoms.

Finally, we aimed to evaluate the alignment of our model probabilities with established biomarker-defined disease stages (A-T-, A+T-, A+MTL+, and A+NEO+) ^4^. A Kruskal-Wallis H test revealed that our composite “AT” score derived from our models’ amyloid and regional tau probabilities significantly differed across disease stages (*H* = 180.73*, p* = 6.15 × 10*^−^*^39^; Fig. 3d). Post-hoc analysis using Dunn’s test with Holm-Bonferroni correction for multiple comparisons demonstrated significant differences between all pairwise stage comparisons, with AT scores progressively increasing from A-T-to A+NEO+ stages. This relationship suggests that our model-derived probabilities capture the biological progression of AD pathology as defined by recently proposed staging systems ^4^. Detailed statistical results are provided in Supplementary Table S16.

### Model ability to capture the synergistic relationship between A*β* and *τ*

To demonstrate the effectiveness of our model for pre-screening in AD clinical trials, we designed a validation approach that aligns with the emerging interest in dual targeting of A*β* and tau pathology, and in stratifying patients by disease burden. Specifically, we assessed the sensitivity of the model outputs to the co-occurring core pathological burden in amyloid PET positive cases. First, we examined how the model’s predicted probability of A*β* positivity, P(A*β*) varied across different levels of tau PET defined pathology. Participants were categorized into two groups based on their meta-*τ* SUVr values: a ‘low/medium’ group (below the 67th percentile) and a ‘high’ group (at or above the 67th percentile). In Fig. 4a, the left panel serves as a reference on the relationship we expect when comparing centiloids and tau PET quantiles in our testing set, showing that centiloid values significantly increased with higher *τ* burden. The one-sided Mann-Whitney U test test confirmed this trend, showing a significant difference in centiloid values across the *τ* tertiles (*U* = 5047, *p* = 1.92×10*^−^*^13^). The right panel presents P(A*β*) between these same quantiles, and similar statistically significant increases in P(A*β*) were seen between the low/medium and high groups (*U* = 3707, *p* = 4.01 × 10*^−^*^20^). These results indicate that the model’s A*β* predictions are sensitive to varying levels of tau burden. Similarly, we assessed how well our model’s *τ* probabilities related to centiloid levels in A*β* PET positive cases. First, we tested the relationship between tau SUVr in the meta-temporal region across tertiles of A*β* centiloids to obtain a reference for the quantitative relationship between A*β* and tau pathologies, as shown in the left panel of Fig. 4b.

**Figure 4:**
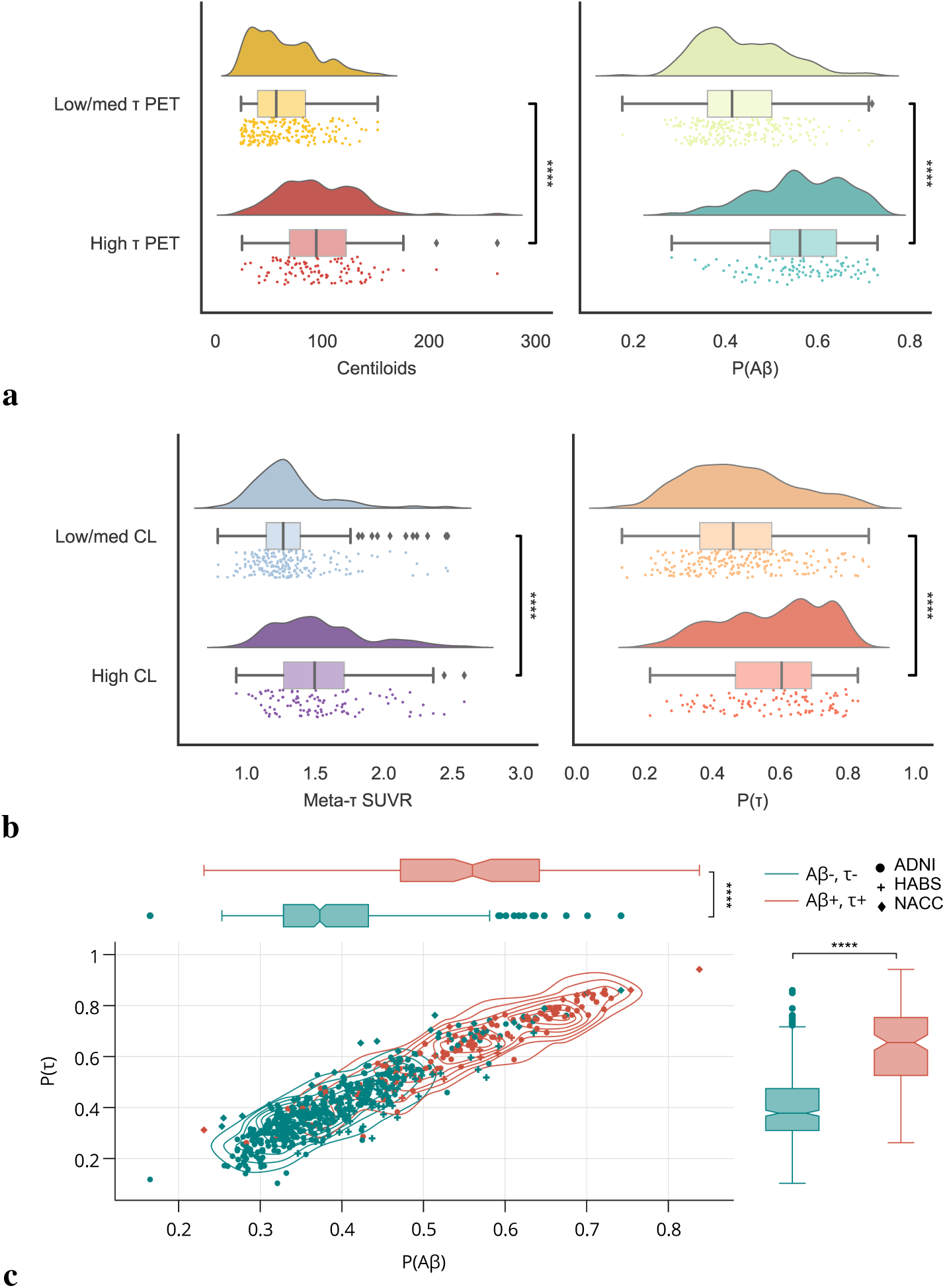
Model ability to capture the synergistic relationship between. **A***β* **and** *τ* **pathologies.** (a) The panel on the left serves as a reference and shows the differences in centiloid distribution in PET-estimated A*β* + individuals across low/medium (*n* = 202) and high (*n* = 102) *τ* PET groups, with the one-sided Mann-Whitney U test indicating significant differences across *τ* groups (*U* = 5047*, p* = 1.92 *×* 10*^−^*^13^). The panel on the right shows the comparisons of our model’s predicted probabilities of A*β* + cases across the same *τ* PET groups (*U* = 3707*, p* = 4.01 *×* 10*^−^*^20^). (b) The left panel shows the comparison of meta-temporal tau SUVr (meta-*τ* SUVr) across low/medium (*n* = 203) and high (*n* = 101) centiloid (CL) groups in A*β* + cases, with the one-sided Mann-Whitney U test pointing to significant differences across CL groups (*U* = 5876*, p* = 6.78*×*10*^−^*^10^). The right panel illustrates the differences in model predicted probabilities across the same CL groups (*U* = 6655.5*, p* = 3.17 *×* 10*^−^*^7^). Cases from the ADNI (*n* = 252) and HABS (*n* = 52) test set were used for rainclouds plots a and b. Detailed statistical tests and results for the data presented in panels a and b can be found in Supplementary Table S17. (c) Kernel density plots comparing model-predicted probabilities of A*β* and *τ* in two distinct A/T profiles (A*β* +,*τ* + and A*β*-,*τ* -), are shown in cases from ADNI, denoted by circles, HABS, denoted by cross symbols, and the held-out NACC* set, denoted by diamond symbols. The color of the points, contour lines and boxplots indicates PET-estimated A*β* +,*τ* + (in red, *n* = 139) and A*β*-,*τ* - (in green, *n* = 500) groups. A one-sided Mann-Whitney U test indicated significant differences in P(A*β*) between negative and positive groups (*n* = 639*, U* = 61430*, p* = 5.71 *×* 10*^−^*^44^) and similarly in P(*τ*) between negative and positive groups (*n* = 639*, U* = 60963*, p* = 1.63 *×* 10*^−^*^42^). All boxplots include a box presenting the median value and interquartile range (IQR), with whiskers extending from the box to the maxima and minima no further than a distance of 1.5 times the IQR. In all the panels, significance levels are denoted as ** for *p <* 0.01; *** for *p <* 0.001; and **** for *p <* 0.0001.

A one-sided Mann-Whitney test indicated that meta-*τ* SUVr was significantly higher in the high CL group relative to the low/medium CL group (*U* = 5876, 6.78 × 10*^−^*^10^). In the right panel, the model’s predictions for tau positivity, P(*τ*), captured similar biological gradients, with a one-sided Mann-Whitney test showing significant differences in P(*τ*) across the same centiloid quantiles (6655.5, *p* = 3.17 × 10*^−^*^07^). Detailed statistical results are reported in Supplementary Table S17. Overall, these results demonstrate our model’s ability to capture the synergistic relationship between A*β* and tau pathologies, reinforcing its potential utility in patient stratification for clinical trials targeting both pathologies individually or together.

We further compared the distributions of our model-predicted probabilities, P(A*β*) and P(*τ*), between participants with the following PET-confirmed biomarker profiles: A*β*-,*τ* - and A*β* +,*τ* + (Fig. 4c). The Mann-Whitney U test revealed significant differences in both P(A*β*) and P(*τ*) between biomarker-positive and biomarker-negative groups (*U* = 61430*, p* = 5.71 × 10*^−^*^44^; *U* = 60963*, p* = 1.63 × 10*^−^*^42^, for A*β* and *τ* respectively). The scatter plots indicate that A*β* +,*τ* + individuals consistently exhibited higher predicted probabilities for both A*β* and *τ* compared to those in the A*β*-,*τ* - group. The associated boxplots and contour plots collectively highlight key differences between the two groups, revealing higher concentrations and a broader distribution of A*β* and *τ* in the A*β* +,*τ* + group compared to the negative group. The results also reveal a greater variability in tau levels for the A*β* +,*τ* + group, with the data extending to higher probabilities. In contrast, the A*β*-,*τ* - group showed a tighter distribution and lower biomarker values.

### Spatial analysis

The accumulation and spatial progression of tau pathology in AD generally follows a stereotypical pattern, beginning in the transentorhinal region, progressing into the limbic system, and eventually spreading to the neocortical associative areas and, ultimately, the primary sensory cortices ^33^. We created a visualization of mean Shapley values for regional volumes across predictions of regional *τ* positivity (Extended Data Fig. 5), ordering them following this stereotypical progression. This visualization underscores the importance of the MTL, which consistently shows high Shapley values, highlighting its role as the initial site of tau deposition and volumetric changes. To further evaluate the model’s decision-making processes when provided with brain regional volumes data, we conducted a graphical analysis to investigate the relative importance attributed to community structures in our model. We then compared the SHAP-derived community structures with tau PET-estimated graphs to assess the alignment between them. The analysis revealed a statistically significant degree of concordance, particularly in the lateral temporal and parietal lobes, suggesting that model-based representations capture meaningful regional distinctions consistent with tau pathology (Fig. 5). Specifically, for the medial temporal *τ* positivity prediction, model-based and reference community structures showed moderate agreement (AMI = 0.219, *p* = 0.0014). The lateral temporal region prediction demonstrated a similar pattern (AMI = 0.176, *p* = 0.0056), while the medial parietal (AMI = 0.134, *p* = 0.0484) and frontal (AMI = 0.138, *p* = 0.0216) predictions exhibited modest similarity. The lateral parietal region achieved the highest agreement (AMI = 0.288, *p* = 0.0016), and the occipital region showed moderate alignment (AMI = 0.233, *p* = 0.0010). Overall, while the partitions in the model-based graphs are not identical to that of the SUVr graphs, there is a non-random correspondence between the two. This supports the idea that the model’s network of regional interactions is reflecting aspects of true tau pathology networks, rather than arbitrary groupings. These findings underscore the interpretability of our approach and its potential to bridge the gap between predictive modeling and biological markers of disease progression.

**Figure 5:**
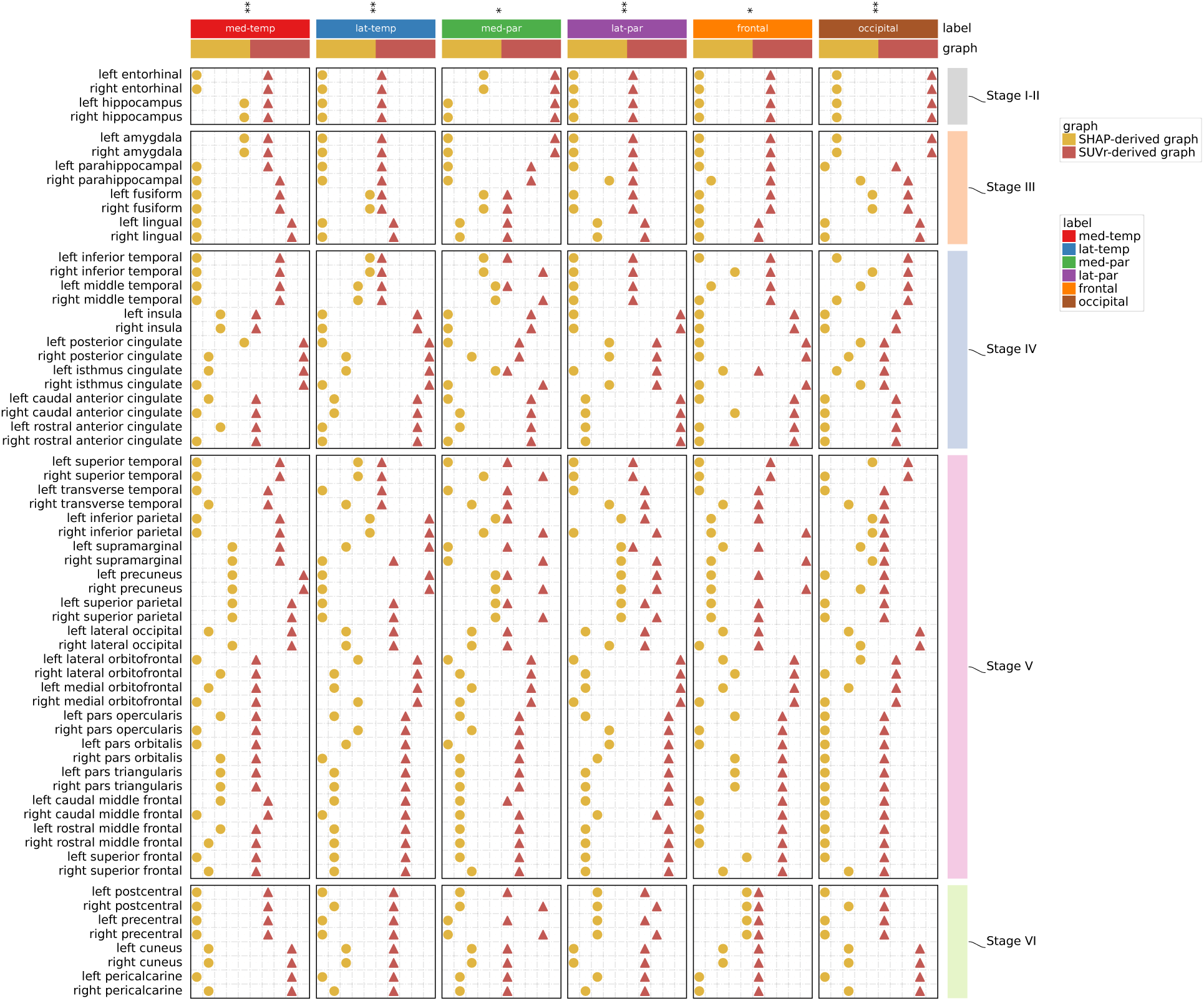
Communities detected from model predictions-driven and tau SUVr-derived graph networks. The dot heatmap visualizes the detected communities within the graph network constructed from normalized mutual information (NMI): one based on Shapley values of T1-weighted regional volumetric features, and the other based on tau PET SUVr within true positive cases. For each of the six regional labels (medial temporal (med-temp), lateral temporal (lat-temp), medial parietal (med-par), lateral parietal (lat-par), frontal, and occipital), communities derived from Shapley values of volumetric features are denoted by yellow dots, and communities derived from tau SUVr are denoted by red up-pointing triangles, with all dots or triangles in the same column as one detected community within the associated graph. Communities are order-invariant. Brain regions are grouped into pre-defined Braak stages (I-II, III, IV, V, and VI) shown on the right for visualization purposes. Statistical annotations denote the results of spatial permutation *t*-tests on the similarity between model-based and tau SUVr-derived communities evaluated by the adjusted mutual information (AMI). Significance levels are denoted as * for *p <* 0.05 and ** for *p <* 0.01

### Neuropathological validation

We validated our model’s predictions of A*β* and tau positivity by comparing them with neuropathological markers of AD. The mean time difference between age at death and age of assessments was 3.05 years. We observed a general increasing trend in the model probabilities with the severity of the pathological markers. Figs. 6a-d illustrate this relationship by comparing the model’s probability scores, P(A*β*) and P(*τ*), against key pathological markers across progressive AD stages: Thal phases of A*β* plaques, Braak stages of neurofibrillary degeneration, and CERAD (Consortium to Establish a Registry for Alzheimer’s Disease) scores for neuritic and diffuse plaques. These markers, denoted as A0-A3 (Thal phases), B0-B3 (Braak stages), and C0-C3 (CERAD scores for neuritic and diffuse plaques) all exhibited a statistically significant upward trend in the median probability of P(A*β*) and P(*τ*) as the stages advanced (*p <* 0.0001 for Thal, Braak, and CERAD stages) (Tables S18 & S19). We also evaluated the model’s predictions in relation to cerebral amyloid angiopathy (CAA) (Fig. 6e), which is commonly ob-served in postmortem AD cases. The model predicted significantly higher P(A*β*) and P(*τ*) in individuals with mild, moderate, or severe CAA compared to those without CAA (*p <* 0.05) (Table S19). These find-ings indicate that our model’s predicted probabilities for A*β* and *τ* positivity closely align with the severity of neuropathological markers, reinforcing the model’s validity in reflecting underlying disease pathology.

**Figure 6:**
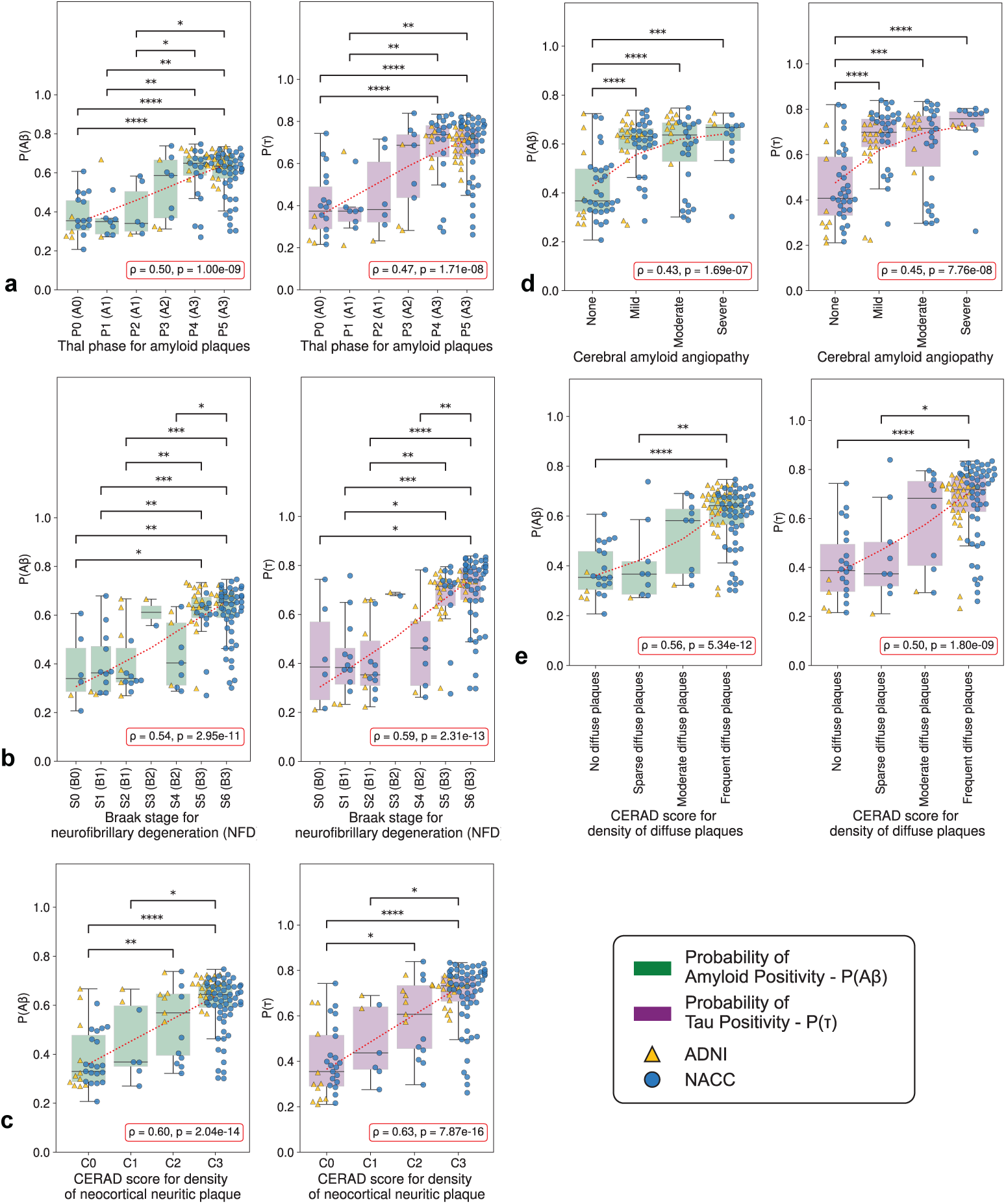
Model alignment with postmortem findings. The swarm and box plots display predicted probabilities of amyloid-beta positivity (P(A*β*)) in green and tau positivity (P(*τ*)) in magenta with respect to various neuropathological grades of Alzheimer’s disease: (a) Thal phase for amyloid plaques, (b) Braak stage for neurofibrillary degeneration (NFD) (c) CERAD score for density of neocortical neuritic plaque, (d) Cerebral amyloid angiopathy, and (e) CERAD score for density of diffuse plaques. Pairwise statistical annotations denote the results of post hoc Dunn tests with Holm-Bonferroni corrections following Kruskal-Wallis test, with significance levels denoted as * for *p <* 0.05; ** for *p <* 0.01; *** for *p <* 0.001; and **** for *p <* 0.0001. Additionally, red trend lines and text boxes in the bottom right of each subplot indicate the Spearman correlation coefficient *ρ* and associated p-value for the overall strength of the correlation between model probabilities and neuropathological grades. The figure legend specifies the magenta and green shades representing P(A*β*) and P(*τ*), respectively, as well as the yellow triangle marker and blue circle marker indicating patients from the ADNI (*n* = 41) and NACC (*n* = 147) neuropathological validation cohorts, respectively. Each boxplot includes a box presenting the median value and interquartile range (IQR), with whiskers extending from the box to the maxima and minima no further than a distance of 1.5 times the IQR. Detailed statistics regarding median values and IQRs can be found in S18 and specific statistics and p-values for Spearman correlation and Kruskal-Wallis tests can be found in S19.

## Discussion

In this work, we present a transformer-based machine learning model that uses multimodal data to predict individual-level A*β* and *τ* PET positivity status in a meta-temporal ROI and in regions associated with progressing disease. Our model achieved strong performance on external data not used for model training, with predictions closely matching postmortem findings. We showed that our model predictions aligned with biological and cognitive outcomes, as well as with disease severity staging, underscoring its clinical relevance. Additionally, the model’s predictions of *τ* pathology in specific ROIs aligned with *τ* burdens derived from regional SUVr observed on PET scans.

Our modeling framework demonstrates flexibility in handling cases with missing features through the use of random feature masking. This approach allows the model to generate predictions and maintain accuracy even when some features are unavailable, which is valuable in settings where all tests are not always accessible for every individual. However, our findings also highlight that certain data inputs, such as neuroimaging and APOE status provide critical information on the underlying pathology, given the improvement in performance upon adding these features (Fig. 2c). For tau predictions, the removal of neuropsychological battery scores significantly reduced the AUPR, underscoring its importance in accurate predictions. On the other hand, our analysis suggests that certain features, such as clinical dementia rating (CDR) scores, could potentially be excluded without significantly compromising the model’s predictive power. This is likely because our framework was developed by fine-tuning a model that already excels at classifying cognitive status^34^. This finding has practical implications for clinical settings, as CDR assessments require a trained clinician to conduct in-depth interviews and additional testing, which can be time-consuming and costly.

Our results indicate that AI models can potentially enhance biomarker-guided assessment of biological AD and facilitate participant selection in clinical trials targeting A*β* and *τ*, either individually or in combination. For example, in AD drug trials, models with high positive predictive values (PPV), can ensure that a higher proportion of individuals flagged as likely to have positive A*β* or tau PET scans are true positives. This could reduce the number of false positives that would need to be excluded later, improving the efficiency and cost-effectiveness of the trial. Additionally, models with a high negative predictive value (NPV) are clinically desirable as they accurately rule out individuals without the condition, reducing the need for unnecessary PET scans and alleviating patient anxiety, thereby lowering both healthcare costs and patient burden. In practice, our AI-based strategy could be integrated into AD screening as follows: persons undergoing neurological evaluation would first be assessed using our AI model, which utilizes clinical and imaging data to predict A*β* and *τ* status. The primary objective of this initial step would be to identify persons who are unlikely to have A*β* or *τ* pathology, thereby ruling out low-risk cases. For individuals whom the AI model does not confidently rule out as being A*β* or *τ* positive, PET imaging would then be recommended. This approach ensures that PET scans are focused on cases where they are most likely to provide a diagnostic benefit. In our testing cohort of 1, 833 individuals with known A*β* PET status, our model predictions demonstrate significant potential for cost savings. With an NPV of 75.35%, we can rule out 587 cases from undergoing unnecessary A*β* PET scans. Similarly, in the test cohort of 844 individuals with known tau PET status, our tau PET model achieved an NPV of 91.65%, suggesting to exclude 582 cases from requiring tau PET scans. Together, these models help avoid unnecessary imaging in low-risk individuals, potentially saving over $3.5 million, based on a $3, 000 cost per scan. Additionally, leveraging the PPV of these models can enhance efficiency by identifying high-risk cases. Our A*β* PET model, with a PPV of 62.05%, can ensure that 654 individuals receive the necessary scans, while the tau PET model, with a PPV of 52.40%, can prioritize 109 high-risk cases for *τ* imaging.

In addition to predicting probabilities for A*β* and tau status, our model provides spatial characterization of the disease, which correlated with disease stage. Our findings further demonstrate that the model-derived volumetric regions of importance align with local patterns of tau deposition observed in PET imaging, thereby validating the model’s predictive capability (Fig. 5). This alignment can aid in differential diagnosis, enable more precise identification of disease stages and subtypes, and support personalized treatment approaches based on regional tau pathology. While neurofibrillary tau tangles are a hallmark of AD, other dementias such as frontotemporal dementia and chronic traumatic encephalopathy can also exhibit tau accumulation ^32,35^. The presence of A*β* and the distribution of tau pathology, however, vary by type of dementia, contributing to diverse clinical presentations and progression patterns ^36,37^. Through providing concurrent predictions of A*β* and *τ* status, our model can aid in increasing specificity to biological AD. In a second stage, our regional tau model could further enhance differential diagnosis by allowing comparison of predicted regional tau profiles with known tau patterns of other dementias. In typical AD, tau burden gradually increases in the medial and neocortical temporal lobes before spreading to the parietal, frontal, and occipital lobes ^33^. We have shown that our model’s composite AT score effectively differentiates between disease stages, distinguishing A+T-cases from A+MTL+ cases, thereby identifying tau pathology in regions that are affected early in the disease course ^4,38^. Because tau PET is closely associated with biological disease stage as well as cognitive decline, it has been proposed as a potential clinical endpoint for disease-modifying treatments ^39^. Our model could thus serve as a pre-screening tool to not only identify the presence of disease but also to delineate the stage of disease, refining the selection of candidates for potential clinical trials or treatments. While our current dataset lacked sufficient data to fully validate the subtyping potential of our model, the comprehensive regional profile of tau pathology it provides could eventually enable clinicians to determine disease stage and subtype based on established tau deposition patterns in AD ^40^. This capability offers promising directions for future research and clinical practice, potentially transforming how AD and related disorders are diagnosed and managed.

Our study has a few limitations. Our model was developed and validated on seven distinct cohorts; however, its generalizability across diverse populations and clinical settings remains to be determined, as the dataset was predominantly composed of White participants. Additionally, we used a binary thresholding technique to define A*β* and tau PET positivity, despite the variability in these definitions across different studies. Various studies have adopted their own criteria for PET positivity, influenced by multiple factors. Nevertheless, our modeling framework is flexible and can be adapted to different definitions of PET positivity (Figs. 3a-b). Future work could extend this binary classification to an ordinal regression task with multiple categories, providing a more quantitative approach to predicting PET status. Moreover, due to the limited number of cases with blood-based biomarker data in our training dataset (*n* = 255), we were unable to fully leverage these data to enhance the model’s predictive accuracy. As novel plasma biomarkers become more widely available, we anticipate that integrating them with existing medical data and neurocognitive evaluations will likely enhance the accuracy of predicting AD pathology than relying on any single modality of data. While our model could help identify individuals likely to have pathology associated with biological AD, extending this framework to select participants for clinical trials is more complex than merely identifying those who are A*β* and *τ* positive. Key barriers include limited awareness, fear of diagnosis, overstretched healthcare systems, poor physician awareness, lack of effective treatments, lack of fast diagnostics, and low awareness of clinical trials, causing many eligible participants to be lost before enrollment. Nevertheless, our framework can provide an important first step in identifying individuals likely to have the disease, thereby enabling more effective targeting of community outreach programs. Additionally, given preliminary evidence that tau PET status and severity may impact treatment response in anti-amyloid therapies ^6^, our model could serve as a tool to predict which patients might benefit most from specific disease-modifying drugs. By stratifying patients based on pathology severity subgroups, clinical trials can be more efficiently designed to assess treatment efficacy in targeted subgroups, potentially improving outcomes and accelerating the development of effective therapies.

In conclusion, by leveraging multimodal data from standard neurological work-up, our model shows promise in identifying individuals with biological AD cost-effectively, reducing the reliance on expensive imaging techniques like PET scans. Such frameworks highlight the potential of advanced machine learning techniques to reduce the burden associated with participant selection for AD clinical trials. Future prospective studies are needed to assess the accuracy of our approach in identifying biological AD and quantify the economic benefits of using this method in selecting participants for clinical trials.

## Methods

### Study population

This study involved a total of 12, 185 participants drawn from seven different cohorts. Written informed consents were obtained from all participants or their proxies, and approval was secured from each cohort’s respective institutional ethical review boards. The training set, consisting of 10, 352 participants, included individuals from the A4 study ^41^, the National Alzheimer’s Coordinating Center (NACC)^42^, the Open Access Series of Imaging Studies (OASIS) ^43^, the Australian Imaging, Biomarkers and Lifestyle (AIBL) study of aging ^44^, and the Framingham Heart Study (FHS ^45^). All subjects in this study had an amyloid PET scan, but only 3, 488 of these participants also underwent tau PET imaging. The training set was further split into training (8281 participants) and validation (2071 participants) subsets using stratified splitting across all labels, ensuring the label distribution remained consistent with the original dataset. The test set comprised 1, 833 participants from the Alzheimer’s Disease Neuroimaging Initiative (ADNI) ^46^, the Harvard Aging Brain Study (HABS) ^47^, and a subset of NACC subjects with neuropathological data.

Data collected included demographics, medical history, neuropsychological scores, physical and neurological examinations, APOE e4 genotype, neuroimaging data, as well as CSF and blood biomarkers for model training. All model evaluations at testing were performed without using CSF. In the study sample, 7, 561 participants were A*β* PET negative and 4, 624 were A*β* PET positive. Among those who underwent tau PET assessments (n=3, 488), 2655 were tau PET negative and 833 were tau PET positive on a meta-temporal region of interest (ROI). Table 1 provides a detailed overview of the study population across all cohorts. Single visits were included for each participant.

### Selection criterion

Participants were eligible for inclusion in the study if they had undergone at least one A*β* PET scan and had clinical or neuroimaging visits within one year of the PET scan. For cohorts with multiple eligible visits such as ADNI, HABS, NACC, OASIS, and AIBL, visits were selected to minimize the time difference between PET scan and clinical or MRI visits. Because OASIS may share participants with ADNI and NACC, we conducted pairwise comparisons between participants in OASIS and ADNI as well as OASIS and NACC - we searched for similar characteristics across demographics, physical characteristics, medical history and comorbidities, functional assessment scores, neuropsychiatric symptoms, and cognitive statuses, with an error tolerance of 2 units in numerical features and excluded any such potentially duplicated participants. All subjects in the A4 cohort with a A*β* PET scan were included. In the FHS cohort, participants with a A*β* PET scan performed within one year after a clinical visit were retained. To ensure consistency across the diverse cohorts, all data were renamed according to the Uniform Data Set Researchers Data Dictionary 3. Despite the unique sets of variables between cohorts, which did not always overlap, no cases were excluded due to missing data. This was facilitated by our model training approach, which incorporated random feature and label masking, as previously reported ^34^.

### PET image processing

Cortical amyloid positivity was quantified using various PET imaging agents in the cohorts: Dynamic 11C-PiB for FHS, late-frame 18F-florbetapen and 18F-florbetapir for ADNI, 18F-florbetapir for A4 and OASIS3, 18F-flutemetamol for AIBL, and 11C-PiB for AIBL, OASIS3, and HABS.

Centiloid (CL) values were provided directly by ADNI, A4, OASIS, and a subset of NACC (*n* = 334) while for AIBL and HABS, an internal pipeline was used to process standard uptake value (SUV) images, following the methodology established by Klunk and colleagues ^48^. Briefly, A*β* PET and T1 weighted (T1w) images were automatically realigned to match the orientation of the MNI152 template. We then coregistered the A*β* PET and T1w MRI images to the MNI152 template, normalized to standard space, and calculated global cortical SUV ratios (SUVr) using the Global Alzheimer’s Association Interactive Network (GAAIN) masks. Our pipeline, which uses SPM12 for image realignment and normalization, differs slightly from the standard Klunk method ^48^, which requires us to process GAAIN data and regress our calculated SUVrs against Klunk’s published values to derive a scaling equation to convert SUVrs to CL for each tracer. For the FHS cohort, mean cortical 11C-PiB distribution volume ratios (DVR) images were estimated using the Logan method ^49^ and these were subsequently processed as described above to calculate global cortical DVR values. DVR images and T1w scans were realigned to the MNI152 orientation, before being co-registered and normalized to standard space. GAAIN masks were finally used to estimate the global cortical DVR. For tau PET, standardized uptake value ratios (SUVr) in Freesurfer-defined regions were made available by the A4, OASIS, FHS, ADNI, HABS and a subset of the NACC cohorts (*n* = 344).

### PET data harmonization

Tau PET data from the various cohorts were processed using different image processing pipelines ^18,50–52^. Therefore, we employed the ComBat tool to harmonize tau PET SUVr values to account for variation across cohorts ^53^. A batch variable for cohort and several covariates were used, including age, sex, amyloid status and diagnosis. We used an analysis of covariance (ANCOVA) framework to assess the main effects of cohorts on tau SUVr measurements across brain regions before and after ComBat harmonization, adjusting for covariates age, sex, diagnosis, and amyloid status. Raw p-values from the ANOVA results were adjusted using the Benjamini-Hochberg procedure to control for the false discovery rate across multiple comparisons. ROIs with an adjusted p-value below 0.05 were considered significant. For SUVr regions where the ANOVA indicated a significant cohort effect post harmonization, post hoc pairwise comparisons were conducted using estimated marginal means. Pairwise contrasts between cohorts were computed with Tukey’s adjustment for multiple comparisons. Please refer to Extended Data Fig. 6 and Supplementary Tables S20 and S21 for more detail on the effect of harmonization.

### PET positivity thresholding and tau profiling

For A*β* PET, a pre-established threshold of 24 CL ^14^ was applied to define positivity in A4, OASIS3, AIBL, HABS, ADNI and the subset of NACC with available CL data. For FHS, a pre-established threshold of 1.2 DVR was used to define A*β* PET positivity ^14^. Most of the NACC subjects included in this study (*n* = 4, 006) were assessed using a binary UDS variable indicating A*β* positivity, and no information was available regarding site-specific thresholding. For tau PET, a meta-temporal region of interest (ROI) was constructed following established standards ^31^. A Gaussian mixture model (GMM) with two components was run on the training dataset and tau PET positivity was defined as SUVr values greater than 1.37. In addition to the meta-temporal ROI, we also defined tau ROIs associated with various AD stages and subtypes: medial temporal, lateral temporal, medial parietal, lateral parietal, frontal and occipital ^17,18^. GMM analyses set the positivity thresholds at 1.32, 1.33, 1.38, 1.29, 1.30 and 1.23, respectively. Table S9 provides an overview of the study population broken down by regional tau positivity status.

### MRI processing

T1-weighted (T1w), FLAIR, and T2*-weighted (T2*w) MRI sequences were collected from various cohorts. Table 1 details the MRI counts for each sequence across these cohorts. T1w images were segmented with Fastsurfer ^54^, and regional volumes were estimated. A Swin UNETR architecture ^55,56^ was further leveraged to extract features from bias field corrected volumetric T1 scans, as well as FLAIR and T2* images that were resampled to 1mm resolution. FLAIR and T2* images were additionally padded to 256 × 256 × 256 before being input to the Swin UNETR architecture. All resulting embeddings were of length 768 × 8 × 8 × 8.

### Modeling framework

We utilized the framework detailed in Xue et. al.,^34^ to analyze 443 distinct clinical features encompassing personal demographics, medical history, functional assessments, neuropsychological test scores, neuroimaging data, and fluid biomarkers (Fig. 1). Each feature was first encoded into a fixed length vector via a modality-specific embedding technique that served as input to the transformer. The transformer then integrated these inputs to generate predictions. A key feature of this model is the implementation of a feature masking mechanism within the transformer, which is designed to handle missing data effectively. The framework also incorporated a label masking strategy to leverage datasets with missing labels. The task was formulated as a multilabel classification problem, with separate binary heads assigned for predicting each label. To account for missing labels, the loss associated with samples lacking specific labels was masked before backpropagation. This approach significantly enhanced the model’s robustness and accuracy in real-world scenarios with incomplete datasets. We fine-tuned this model, originally trained on a 13-label classification task ^34^, using a two-stage process. In the first stage, we trained the model to predict A*β* and meta-*τ* labels by transferring the weights of the transformer encoder module and the embedding modules corresponding to overlapping features. During the initial 15 epochs, only the newly initialized weights were trained, while the transferred weights remained frozen. Subsequently, we unfroze the transferred weights and included them in the training process. In the second stage, we further fine-tuned the model to predict regional *τ* labels. To prevent label leakage, we maintained the same training and testing splits for the NACC dataset as in the original transformer protocol ^34^, ensuring no subject overlap between the two sets.

### Loss function

Our model was trained by minimizing the “Focal Loss (FL)” ^57^ (L), along with the standard L2 regularization term. FL is a variant of standard cross-entropy loss that addresses the issue of class imbalance. It assigns low weight to easy (well-classified) instances and high weight to hard-to-classify examples. This loss function was used for each of the biomarker categories. Therefore, our L_FL_ term was:

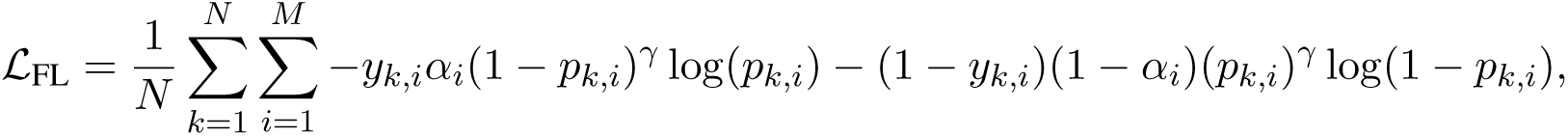

where *N* was the batch size and *M* was the number of biomarker categories (2 for the first stage and 6 for the second), and other parameters and variables were as defined. The focusing parameter *γ* was set to 2, which had been reported to work well in previous studies ^34,57^. Moreover, *α_i_* ∈ [0, 1] was the balancing parameter that influenced the weights of positive and negative instances. It was set as the square of the complement of the fraction of samples labeled as 1, varying for each *i* due to the differing level of class imbalance across biomarker categories. After combining all terms, the overall loss function (L) was:

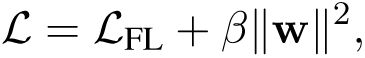

where *β* controlled the importance of the L2 regularization term. For both stages of training, the maximum epochs was set to 128 epochs, with early stopping applied if no improvement was observed on the validation split for 15 epochs in the first stage and 30 epochs in the second. The L2 regularization parameter *β* was set to 0.01 for the first stage and 0.005 for the second. Mini-batch training was performed using the AdamW optimizer ^58^, with learning rates of 0.001 and 0.0001 for the respective stages. Additionally, a cosine scheduler was employed to dynamically adjust the learning rate during training.

### Interpretability analysis

To interpret the model predictions, we conducted Shapley analysis ^59^ on the outputs for A*β*, meta-*τ*, and regional *τ* models. Shapley values quantify the contribution of each feature to the model’s predictions, effectively providing a measure of feature importance. We employed a permutation sampling strategy ^34,60^ to efficiently estimate Shapley values across the high-dimensional feature space. This approach involves permuting feature values and measuring changes in the model’s output to approximate each feature’s impact. For each label prediction, Shapley values were calculated for all input features, including imaging-derived measures, whole brain image embeddings and clinical variables. Missing features were assigned a Shapley value of zero, indicating no contribution to the prediction. The features were then ranked by their mean Shapley values across true positive samples, identifying the most influential features driving the model’s decisions.

### Traditional machine learning model

We sought to compare the performance of our model with that of a traditional machine learning framework, CatBoost ^61^, to provide a benchmark for our approach. As a tree-based classification framework, CatBoost effectively handles missing features by assigning designated missing values when an input is absent at inference. However, CatBoost lacks support for incorporating learned embeddings from imaging data, limiting its ability to leverage spatial patterns captured in MRI scans. To address this, we used regional volumes derived from FastSurfer as the imaging-related inputs for CatBoost. Additionally, unlike our transformer-based model, which performs multi-label classification in a unified manner, CatBoost requires training separate models for each output variable. As a result, we trained eight independent CatBoost models, one for each label, while our deep learning approach benefited from joint optimization across multiple tasks.

### Model validation on biological outcomes

We sought to validate predicted probabilities of the model against PET estimates of amyloid and tau burden, as well as evaluate its alignment with a common clinical endpoint in AD clinical trials, the Alzheimer’s Disease Assessment Scale-Cognitive Subscale (ADAS-Cog_13_). Importantly, ADAS-Cog_13_ scores were not incorporated as input during the model’s training, ensuring independent validation of the model’s predictive capabilities. Participants from the ADNI cohort were selected for this analysis, as they both underwent amyloid and tau PET imaging and completed the ADAS-Cog_13_ assessment. To further evaluate our model performance in preclinical AD, we included a subset of cases from the ADNI and HABS cohort who were cognitively unimpaired. We then compared model predicted probabilities for amyloid, P(A*β*), between cases who were A*β* PET negative vs A*β* PET positive. Finally, we aimed to validate our model predictions of regional tau positivity and investigate its potential for disease staging. To derive a unified quantification of AD pathology, we employed principal component analysis (PCA). This dimensionality reduction technique allowed us to capture the shared variance across different regional tau and amyloid probabilities into a single composite score. We applied PCA and used the first principal component (PC1), which explained 97.5% of the variance, as our composite measure of AD pathology, termed the amyloid-tau (AT) score. Based on the PET binary labels, we classified participants and compared the AT scores across four distinct disease stages. These included cases who are A*β* PET negative and tau PET negative in all regions (A-T-), A*β* positive but tau negative in all regions (A+T-), A*β* positive with tau PET positivity restricted to the medial temporal lobe (A+MTL+), and A*β* positive cases with tau PET positivity in the medial temporal and neocortical regions (A+NEO+).

### Subgroup analysis on biomarker profiles

We selected a subset of cases from the testing set with PET-confirmed A*β* positivity, mirroring the inclusion criteria for amyloid presence used in recent clinical trials [REF]. Participants were then stratified into tertiles (low, medium, and high) based on their meta-*τ* SUVr values to evaluate the model’s predictive accuracy across a spectrum of tau burdens. We further assessed the relationship between tertile groups and centiloids to evaluate whether the model’s output is consistent with empirically measured amyloid levels. Similarly, we conducted an analysis of the model-predicted tau probabilities, P(*τ*), in A*β* + cases, this time stratifying participants into tertiles based on their centiloid values. Because the NACC* testing cohort did not have continuous PET data available, only ADNI and HABS were included in these analyses. Finally, to further validate our model’s ability to differentiate those who are positive on both biomarkers from those who are negative on both, we compared the distributions of P(A*β*) and P(*τ*) in the combined ADNI, HABS and NACC* test set between A*β* +, *τ* + and A*β*-, *τ* - cases.

### Spatial analysis

Cases with positive regional *τ* labels and predictions were selected for this data-driven analysis. A fully-connected graph network was constructed with nodes representing individual brain regions and edges connecting the nodes. Edge weights were determined by computing pairwise normalized mutual information (NMI) ^62–64^ on the Shapley values of T1-derived regional volumetric features. This quantifies the mutual dependence between two brain regions in their contribution to the model. We identified non-overlapping communities of brain regions that the model deemed important for positive predictions on each regional label using the Louvain method for community detection ^65^. We preset the number of communities in each graph to five, corresponding to the established Braak staging of tau pathology progression, combining regions from stages 1 and 2 ^33^. To address the randomness inherent in the Louvain algorithm, we employed consensus clustering with 100 draws ^63^. Using the same set of cases, we established another graph network on the same brain regions, but with edges defined by the NMI of the tau PET SUVr values.

We identified communities of brain regions in this network using the same methodology as before. To compare the T1-derived communities identified as important by the model against the communities identified in the tau PET scan, we evaluated the similarity between these two clusterings using the adjusted mutual information (AMI) ^66^. The AMI measures the level of agreement between two clusterings with correction for random clustering agreement, and is preferred over adjusted Rand index (ARI) when the reference clustering is unbalanced and there exist small clusters ^67^, which aligns well with our results (Table S22). A *t*-test on 5, 000 spatial permutation draws was conducted on AMI for assessing statistical significance ^68^,^69^. Spatial permutations were applied to maintain the brain’s contralateral symmetry through rotating spherically projected brain region coordinates extracted from the Desikan-Killiany atlas by a random angle along each of the x, y, and z axes. New labels were assigned by mapping the original region centroids to the closest permuted region centroid based on Euclidean distance.

### Postmortem validation

To assess the alignment of our model with neuropathological evidence, we utilized a subset of cases from the ADNI database (*n* = 41) for which postmortem evaluations were available. We supplemented this sample with an additional subset of cases from the NACC database (*n* = 147) for which neuropathological data was available, excluding these cases from the training set. Of note, this subset of NACC cases was also in the testing set of the original transformer model ^34^ that we are finetuning in this study, thus preventing potential label leakage. The average time lag between the visit selected for model development and the time of death was 3 years on average. On these cases, we examined the Thal phase for amyloid plaques (A score), Braak stage for neurofibrillary degeneration (B score), density of neocortical neuritic plaques (CERAD score) (C score), density of diffuse plaques (CERAD semi-quantitative score), and cerebral amyloid angiopathy, and investigated the correlation between the model-generated probability scores of A*β* and *τ* positivity and the grades of these neuropathological features.

### Statistical analyses

We conducted a series of statistical analyses to rigorously evaluate our AI model’s alignment with PET burden, biomarker profiles, and post-mortem neuropathological grades. A Shapiro–Wilk test was performed prior to each analysis to assess normality. To evaluate the alignment between our model-predicted probabilities and continuous PET values, we computed both the Spearman’s *ρ* and Pearson’s *r* coefficients, log-transforming regional *τ* SUVr values to improve linearity. We then sought to validate our model’s predictive accuracy across quantiles of disease severity. We used a one-sided Mann-Whitney U test to compare predicted probabilities (P(A*β*) and P(*τ*)) and PET measures (centiloids and meta-T SUVr) between cases with low/medium vs. high disease burden. Similarly, we applied a one-sided Mann-Whitney U test to compare P(A*β*) and P(*τ*) between cases who are PET-confimed biomarker positive and negative. Additionally, we evaluated the model’s ability to detect preclinical AD by comparing amyloid probability outputs between A*β* PET negative and positive cognitively unimpaired cases using a one-sided Mann-Whitney U test. We then aimed to validate our model’s ability to distinguish disease stages. A Kruskal-Wallis H test, followed by post hoc Dunn’s test with Holm-Bonferroni adjustments for multiple comparisons was performed to assess the alignment of our model’s AT score with PET-defined disease stages. To evaluate differences in model probability outputs across various stages of neuropathological scores, we employed the Kruskal-Wallis test, followed by post hoc Dunn’s tests to conduct pairwise comparisons between groups, with adjustments for multiple comparisons using the Holm-Bonferroni correction method. Additionally, to assess the overall correlation between the model-generated probabilities and each neuropathological feature, we computed the Spearman correlation coefficient, assessing the strength and direction of association between the ranked neuropathological grades and model probabilities.

### Performance metrics

Receiver operating characteristic (ROC) and precision-recall (PR) curves were created based on the predictions on the combined ADNI and HABS external datasets, as well as on the NACC test set. Additional performance metrics including balanced accuracy, sensitivity, specificity, precision (positive predictive value), F1 score, Matthews correlation coefficient, and negative predictive value (NPV) were computed by determining the optimal threshold for each label using Youden’s J statistic, based on the performance of the validation split.

### Computational hardware and software

Our software development utilized Python (version 3.11.9) and the models were developed using PyTorch (version 2.4.0). We used several other Python libraries to support data analysis, including pandas (version 2.2.2), numpy (version 1.26.3), matplotlib (version 3.9.1), monai (version 1.3.2), scipy (version 1.14.0), and scikit-learn (version 1.5.1). Several R packages were also used for data analysis and visualization, including dplyr, emmeans, and ggseg3D. Training the model on a single Tesla V100 GPU on a shared computing cluster had an average runtime of 2 minutes per epoch, while the inference task took less than a minute per instance. All figures were prepared using Canva and Adobe Illustrator.

## Data availability

Data from A4, AIBL and ADNI are available to download from the LONI website at https://ida.loni.usc.edu/. NACC and OASIS-3 data can be requested and downloaded at https://naccdata.org and https://sites.wustl.edu/oasisbrains/, respectively. FHS data (https://www.framinghamheartstudy.org/fhs-for-researchers/data-available-overvie can be requested by emailing, and access conditions include completing the steps outlined at https://www.framinghamheartstudy.org/fhs-for-researchers/, as well as approval from the FHS Research Committee. HABS data can be requested at https://habs.mgh.harvard.edu/researchers/request-data/. All data used in this study should be available free of charge upon request from the specific cohorts.

## Data Availability

Data from A4, AIBL and ADNI are available to download from the LONI website at \url{https://ida.loni.usc.edu}. NACC and OASIS-3 data can be requested and downloaded at \url{https://naccdata.org} and \url{https://sites.wustl.edu/oasisbrains/}, respectively. FHS data (\url{https://www.framinghamheartstudy.org/fhs-for-researchers/data-available-overview/}) can be requested by emailing \url{}, and access conditions include completing the steps outlined at \url{https://www.framinghamheartstudy.org/fhs-for-researchers/}, as well as approval from the FHS Research Committee. HABS data can be requested at \url{https://habs.mgh.harvard.edu/researchers/request-data/}. All data used in this study should be available free of charge upon request from the specific cohorts.

## Acknowledgements

This project was supported by grants from the National Institute on Aging’s Artificial Intelligence and Technology Collaboratories (P30-AG073105, VBK), the American Heart Association (20SFRN35460031, VBK & RA), Gates Ventures (RA & VBK), and the National Institutes of Health (R01-HL159620 [VBK], R01-AG062109 [RA & VBK], and R01-AG083735 [RA & VBK]). We acknowledge the efforts of several individuals from the A4, ADNI, AIBL, FHS, HABS, NACC, and OASIS for providing access to data. We thank Mr. Chonghua Xue, Mr. Osman B. Guney and Dr. Derek Archer for several useful discussions. We also thank Dr. Christina Young for sharing processed tau PET data for the FHS cohort, and for several valuable discussions.

The NACC database is funded by NIA grant U24-AG072122. NACC data are contributed by the NIA-funded ADRCs: P30 AG062429 (PI James Brewer, MD, PhD), P30 AG066468 (PI Oscar Lopez, MD), P30 AG062421 (PI Bradley Hyman, MD, PhD), P30 AG066509 (PI Thomas Grabowski, MD), P30 AG066514 (PI Mary Sano, PhD), P30 AG066530 (PI Helena Chui, MD), P30 AG066507 (PI Marilyn Albert, PhD), P30 AG066444 (PI John Morris, MD), P30 AG066518 (PI Jeffrey Kaye, MD), P30 AG066512 (PI Thomas Wisniewski, MD), P30 AG066462 (PI Scott Small, MD), P30 AG072979 (PI David Wolk, MD), P30 AG072972 (PI Charles DeCarli, MD), P30 AG072976 (PI Andrew Saykin, PsyD), P30 AG072975 (PI David Bennett, MD), P30 AG072978 (PI Neil Kowall, MD), P30 AG072977 (PI Robert Vassar, PhD), P30 AG066519 (PI Frank LaFerla, PhD), P30 AG062677 (PI Ronald Petersen, MD, PhD), P30 AG079280 (PI Eric Reiman, MD), P30 AG062422 (PI Gil Rabinovici, MD), P30 AG066511 (PI Allan Levey, MD, PhD), P30 AG072946 (PI Linda Van Eldik, PhD), P30 AG062715 (PI Sanjay Asthana, MD, FRCP), P30 AG072973 (PI Russell Swerdlow, MD), P30 AG066506 (PI Todd Golde, MD, PhD), P30 AG066508 (PI Stephen Strittmatter, MD, PhD), P30 AG066515 (PI Victor Henderson, MD, MS), P30 AG072947 (PI Suzanne Craft, PhD), P30 AG072931 (PI Henry Paulson, MD, PhD), P30 AG066546 (PI Sudha Seshadri, MD), P20 AG068024 (PI Erik Roberson, MD, PhD), P20 AG068053 (PI Justin Miller, PhD), P20 AG068077 (PI Gary Rosenberg, MD), P20 AG068082 (PI Angela Jefferson, PhD), P30 AG072958 (PI Heather Whitson, MD), P30 AG072959 (PI James Leverenz, MD).

The ADNI database is funded by NIA grant U01-AG024904. ADNI is funded by the National Institute on Aging, the National Institute of Biomedical Imaging and Bioengineering, and through generous contributions from the following: AbbVie, Alzheimer’s Association; Alzheimer’s Drug Discovery Foundation; Araclon Biotech; BioClinica, Inc.; Biogen; Bristol-Myers Squibb Company; CereSpir, Inc.; Cogstate; Eisai Inc.; Elan Pharmaceuticals, Inc.; Eli Lilly and Company; EuroImmun; F. Hoffmann-La Roche Ltd and its affiliated company Genentech, Inc.; Fujirebio; GE Healthcare; IXICO Ltd.; Janssen Alzheimer Immunotherapy Research & Development, LLC.; Johnson & Johnson Pharmaceutical Research & Development LLC.; Lumosity; Lundbeck; Merck & Co., Inc.; Meso Scale Diagnostics, LLC.; NeuroRx Research; Neurotrack Technologies; Novartis Pharmaceuticals Corporation; Pfizer Inc.; Piramal Imaging; Servier; Takeda Pharmaceutical Company; and Transition Therapeutics. The Canadian Institutes of Health Research is providing funds to support ADNI clinical sites in Canada. Private sector contributions are facilitated by the Foundation for the National Institutes of Health http://www.fnih.org/(www.fnih.org). The grantee organization is the Northern California Institute for Research and Education, and the study is coordinated by the Alzheimer’s Therapeutic Research Institute at the University of Southern California. ADNI data are disseminated by the Laboratory for Neuro Imaging at the University of Southern California. Data used in the preparation of this article was obtained from the Australian Imaging Biomarkers and Lifestyle flagship study of ageing (AIBL) funded by the Commonwealth Scientific and Industrial Research Organisation (CSIRO) which was made available at the ADNI database (www.loni.usc.edu/ADNI). The AIBL researchers contributed data but did not participate in analysis or writing of this report. AIBL re-searchers are listed at www.aibl.csiro.au.

Data were provided in part by OASIS-3 Longitudinal Multimodal Neuroimaging (Principal Investigators: T. Benzinger, D. Marcus, J. Morris) funded by NIH P30 AG066444, P50 AG00561, P30 NS09857781, P01 AG026276, P01 AG003991, R01 AG043434, UL1 TR000448, R01 EB009352. AV-45 doses were provided by Avid Radiopharmaceuticals, a wholly owned subsidiary of Eli Lilly. Data were also provided in part by OASIS-3 AV1451 (Principal Investigators: T. Benzinger, J. Morris) P30 AG066444, AW00006993. AV-1451 doses were provided by Avid Radiopharmaceuticals, a wholly owned subsidiary of Eli Lilly.

The A4 study is a secondary prevention trial for preclinical Alzheimer’s disease that aims to slow cognitive decline by targeting brain amyloid buildup in clinically unimpaired elderly individuals. It is supported by a diverse funding coalition that includes the National Institutes of Health–National Institute on Aging, Eli Lilly and Company, the Alzheimer’s Association, the Accelerating Medicines Partnership, GHR Foundation, an unnamed foundation, and other private contributors, along with in-kind contributions from Avid and Cogstate. Additionally, the related observational Longitudinal Evaluation of Amyloid Risk and Neurodegeneration (LEARN) Study is funded by the Alzheimer’s Association and GHR Foundation. Dr. Sperling from Brigham and Women’s Hospital and Harvard Medical School, along with Dr. Paul Aisen from the Alzheimer’s Therapeutic Research Institute (ATRI) at the University of Southern California, lead the A4 and LEARN Studies. ATRI coordinates these studies, and the data is accessible through the Laboratory for Neuro Imaging at the University of Southern California.

The Harvard Aging Brain Study (HABS), initiated in 2010, provided data for this manuscript (P01AG036694; https://habs.mgh.harvard.edu). The study, supported by the National Institute on Aging, is directed by principal investigators R. A. Sperling and K. A. Johnson at the Massachusetts General Hospital/Harvard Medical School in Boston.

## Author contributions

V.H.J. and S.P. performed data collection. S.S.K. designed and developed the machine learning framework. V.H.J., S.S.K., S.P., M.F.R. and L.X. performed model training and validation. V.H.J., S.P., and L.X. performed statistical analysis. V.H.J., S.S.K., S.P., M.F.R. and L.X. generated figures and tables. R.A. provided access to data. V.B.K. and V.H.J. wrote the manuscript. All authors reviewed, edited and approved the manuscript. V.B.K. conceived, designed and directed the study.

## Ethics declarations

V.B.K. is a co-founder and equity holder of deepPath Inc., and CogniScreen, Inc. He also serves on the scientific advisory board of Altoida Inc. R.A. is a scientific advisor to Signant Health and NovoNordisk. The remaining authors declare no competing interests.

## Extended data

**Extended Figure 1:**
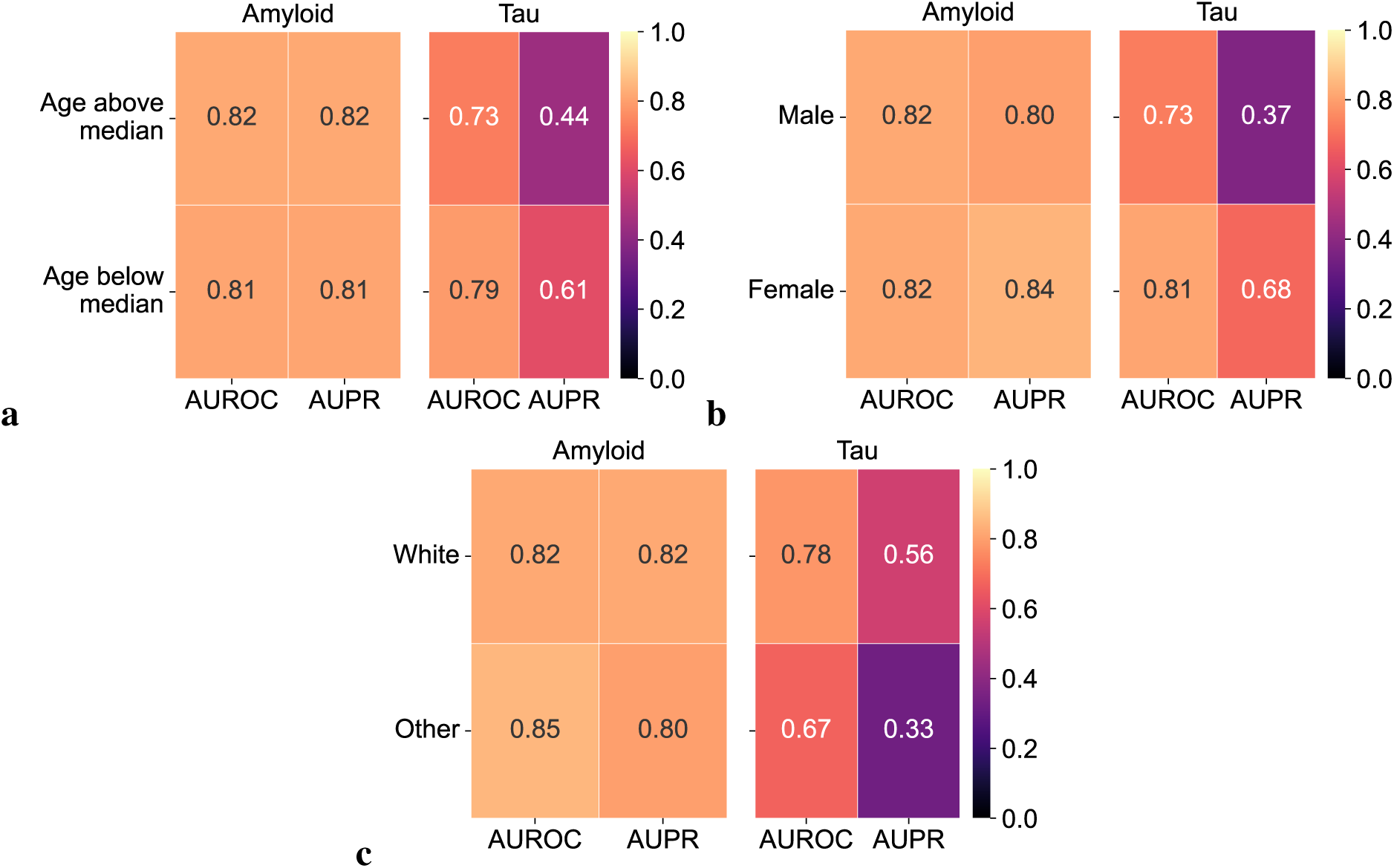
Model performance across demographic subgroups. (a) Area under the receiver operating characteristic (ROC) and precision-recall (PR) curves for A*β* and meta-*τ* predictions in individuals younger than 73 years (A*β*: *n* = 917, *τ*: *n* = 415) and those aged 73 years and older (A*β*: *n* = 916, *τ*: *n* = 428). The median age of the testing population is 73 years. (b) Area under the ROC and PR curves for A*β* and meta-*τ* predictions stratified by age groups: individuals younger than 73 years (A*β*: *n* = 917, *τ*: *n* = 415) and those aged 73 years and older (A*β*: *n* = 916, *τ*: *n* = 428). The median age of the testing population is 73 years. (c) Area under the ROC and PR curves for A*β* and meta-*τ* predictions stratified by sex: (a, b) individuals identified as male (A*β*: *n* = 905, *τ*: *n* = 387) and (c, d) individuals identified as female (A*β*: *n* = 928, *τ*: *n* = 456).

**Extended Figure 2:**
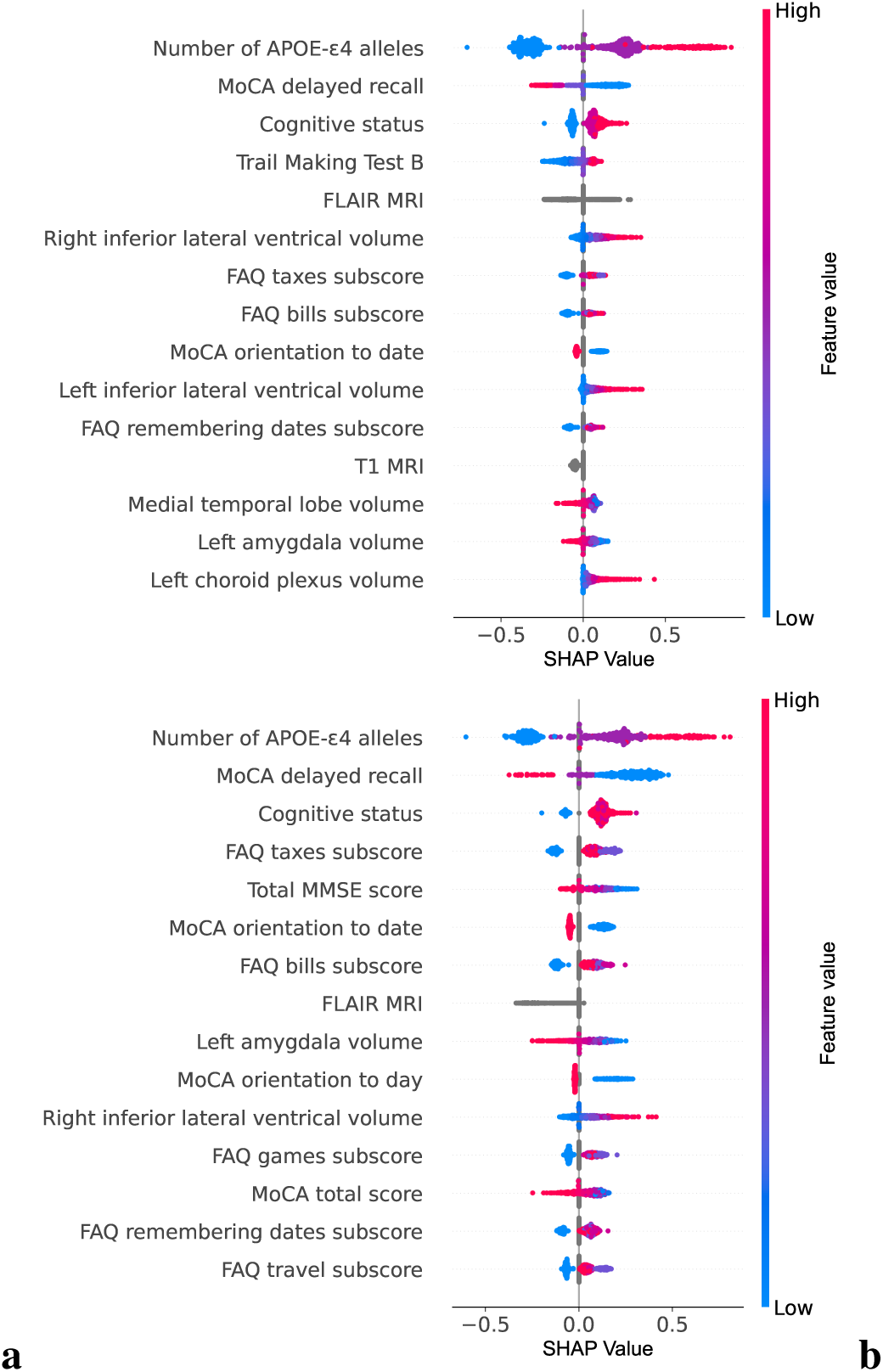
Shapley analysis. The figure presents the top fifteen contributing features for the model’s positive predictions of (a) A*β* and (b) *τ* labels, ranked by their mean Shapley values. These values, which represent the average contribution of each feature to the model’s decision, were used to rank the features from highest to lowest impact.

**Extended Figure 3:**
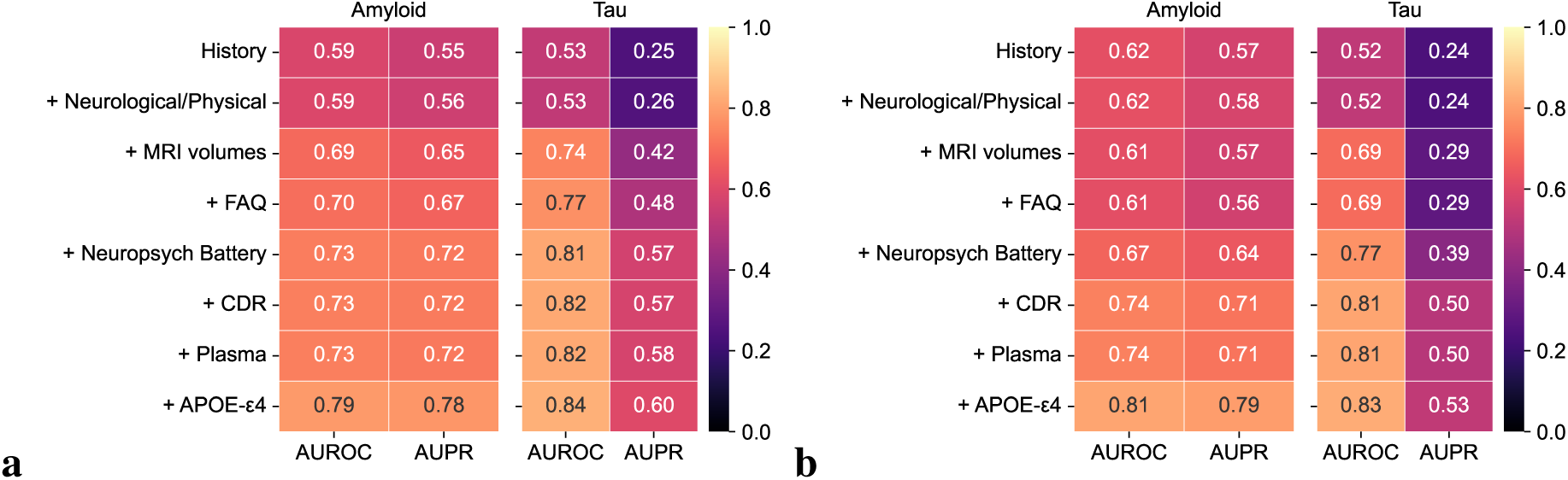
AUROC and AUPR performance using incremental clinical features. Heatmap presenting the AUROC and AUPR values for A*β* and meta-*τ* predictions using various combinations of clinical features for (a) our model without image embeddings (b) CatBoost.

**Extended Figure 4:**
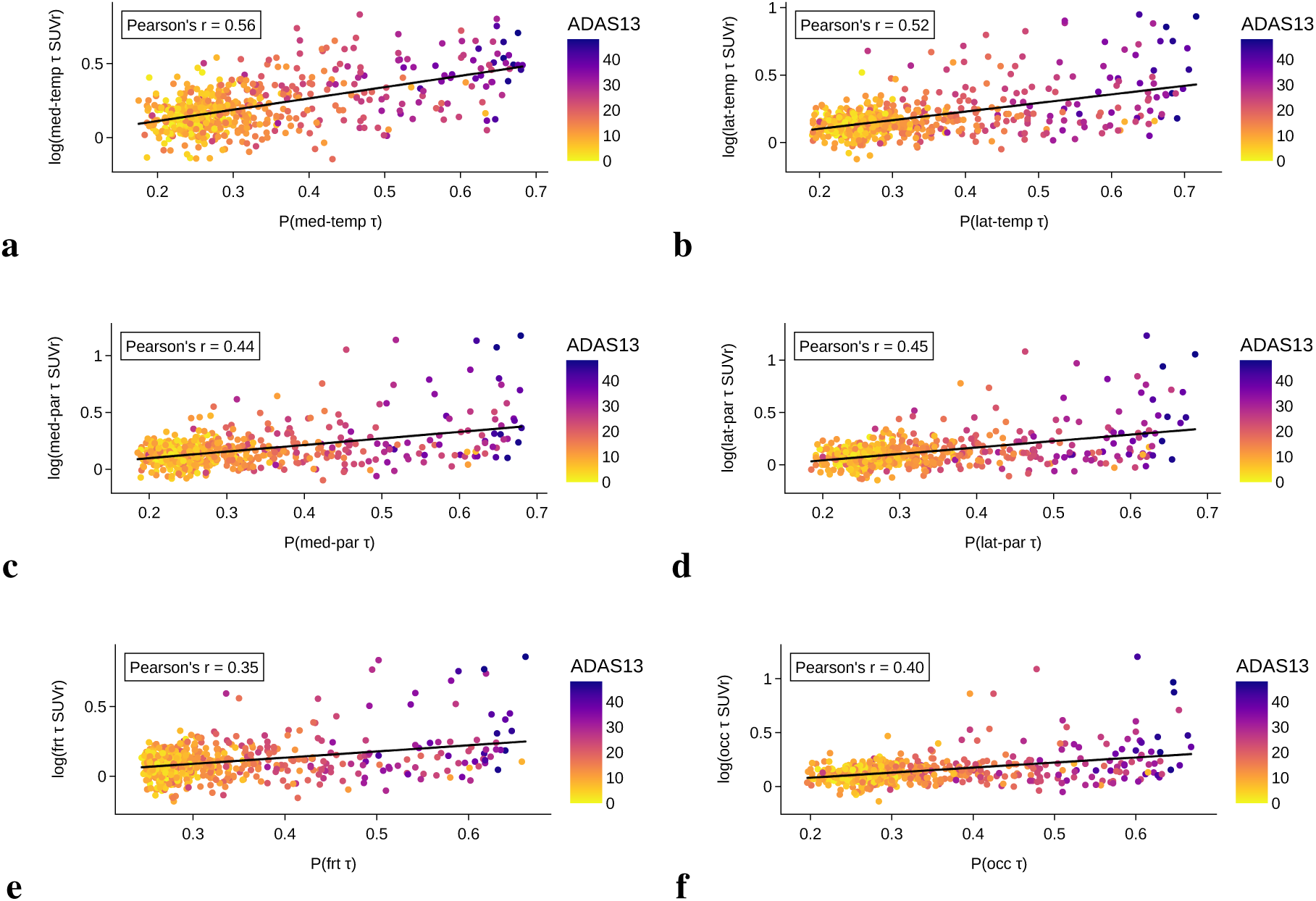
Regional tau model’s alignment with biological outcomes. The bubble plots illustrate model-predicted probabilities of regional *τ* positivity with the respective regional *τ* PET SUVr values. SUVr values were log transformed before computing Pearson’s r correlations. Detailed statistics are reported in Table S15, including Pearson’s and Spearman’s correlation coefficients with p-values.

**Extended Figure 5:**
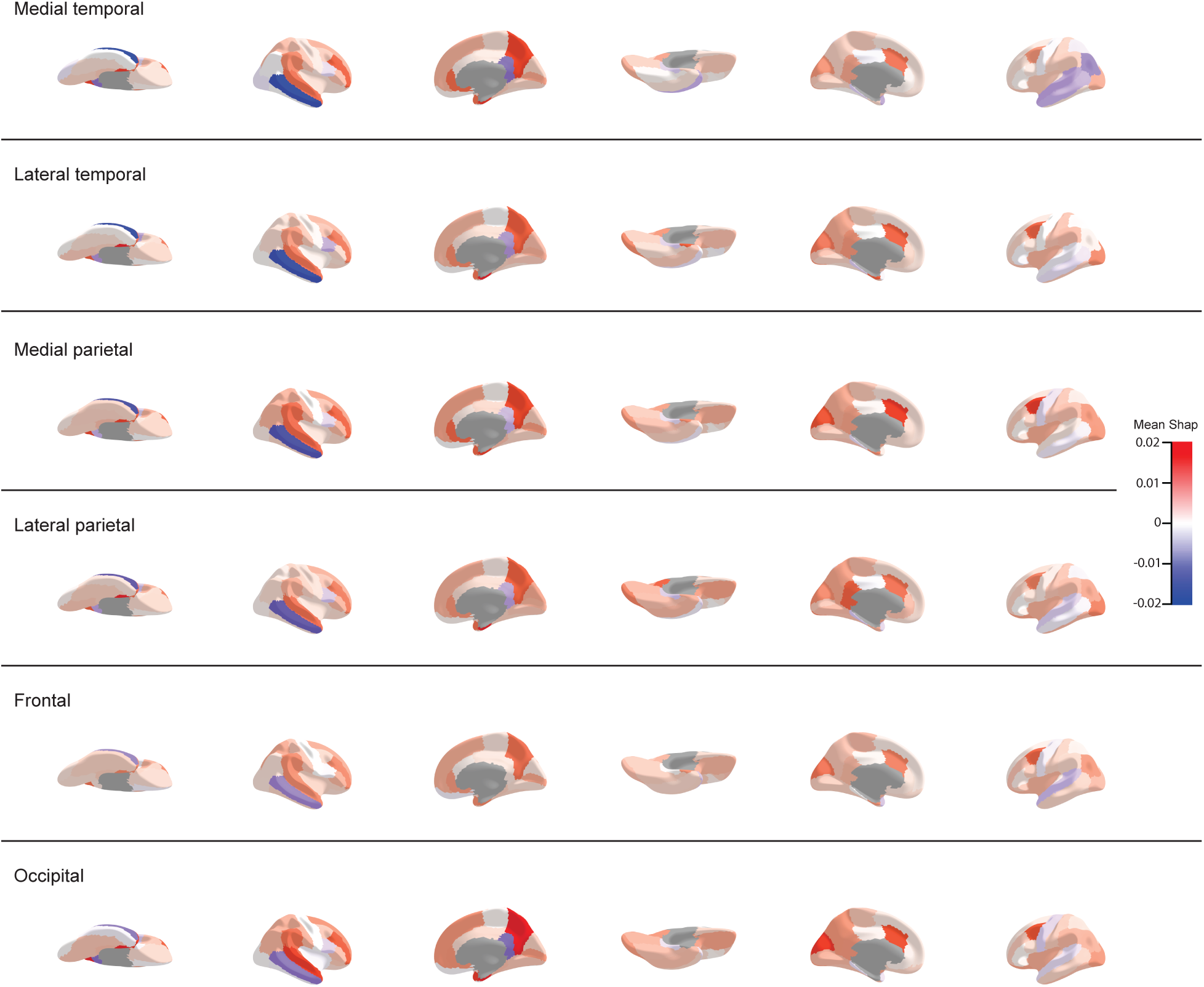
Inflated cortical surfaces showing Shapley values of regional brain volumes for regional tau predictions. Shapley values are overlaid on the Desikan-Killiany-Tourville atlas to illustrate the relative importance of different cortical volumes in predicting regional tau positivity. The first panel displays the mean Shapley values for the medial temporal predictions and subsequent panels are ordered according to Braak stages.

**Extended Figure 6:**
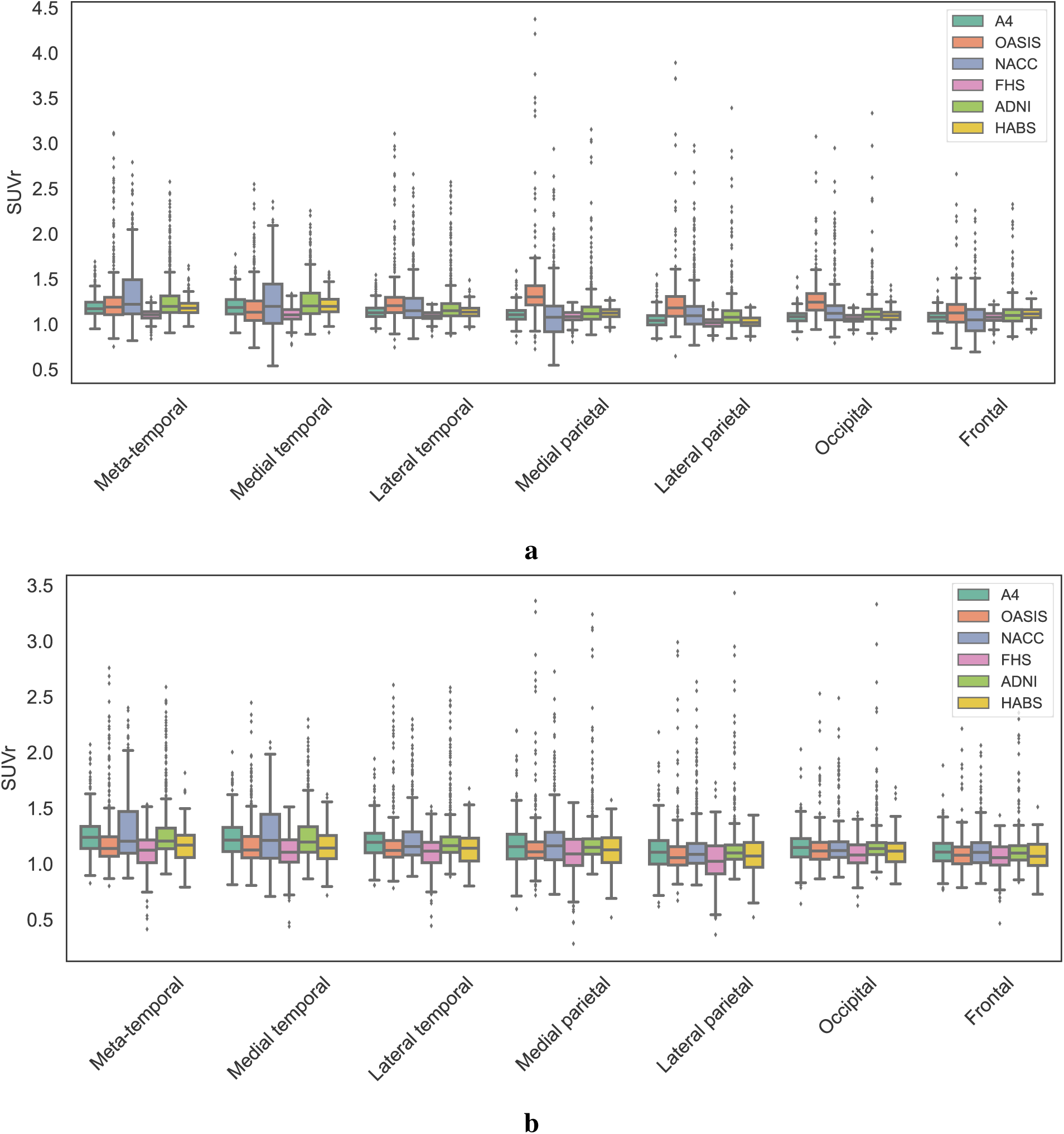
Impact of harmonization on SUVr distributions across cohorts. The top panel presents boxplots of unharmonized SUVr values for each *τ* PET ROI, while the bottom panel displays SUVr values after harmonization. Each box represents the distribution of SUVr within a cohort, with different colors indicating different cohorts. Harmonization reduced the batch effect of cohort-specific variability, while controlling for meaningful covariates. Detailed statistics are reported in Supplementary Tables S20 and S21.

---

**Extended Table 1:**
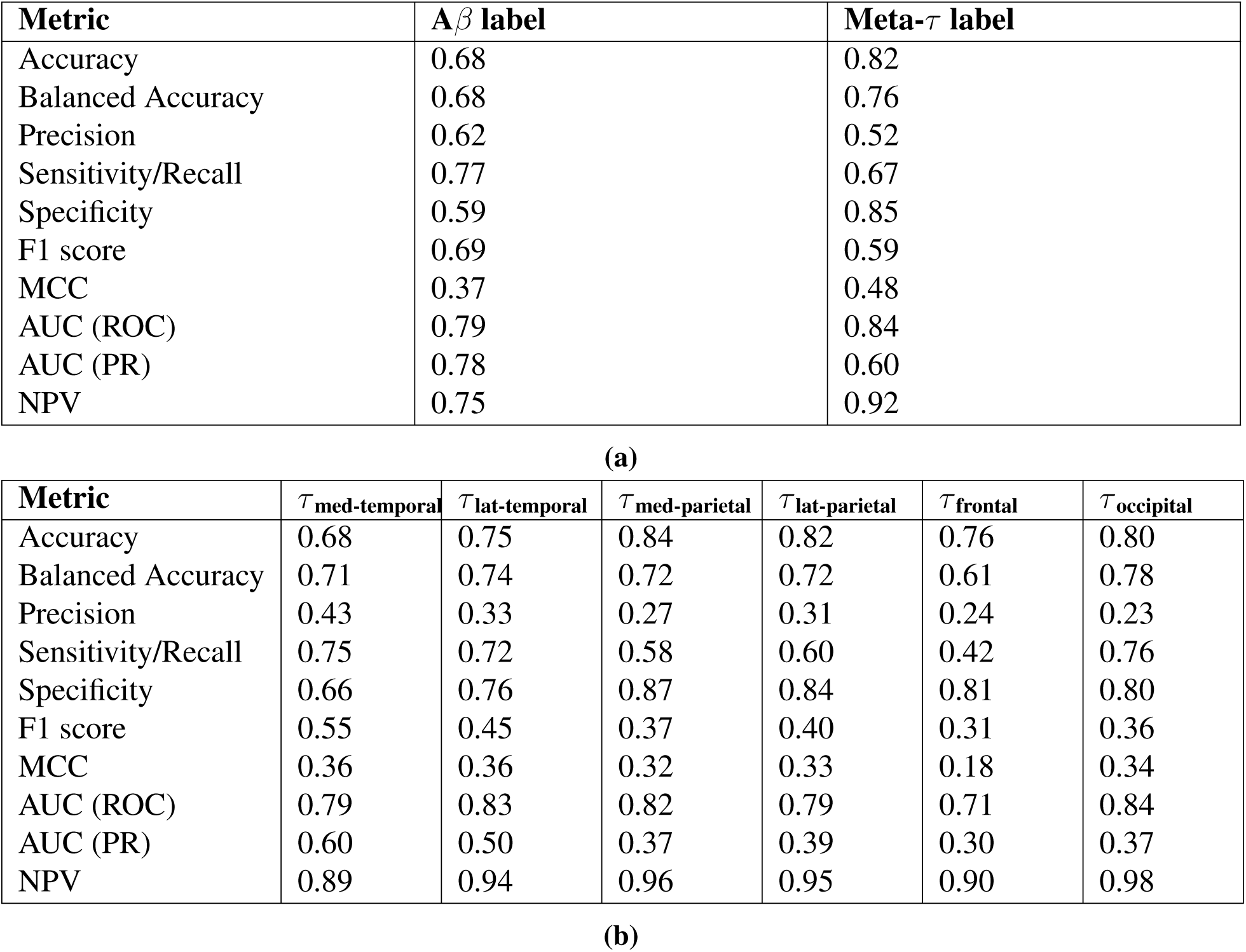
Model performance. Performance metrics for A*β* and *τ* labels on the combined ADNI (A*β*: *n* = 1404, *τ*: *n* = 625), HABS (A*β*: *n* = 282, *τ*: *n* = 173) and NACC* (A*β*: *n* = 147, *τ*: *n* = 45) cohorts are shown. Table (a) presents the performance metrics for A*β* and meta-*τ* labels, and table (b) presents the performance metrics for the regional *τ* labels. Note that all available data from ADNI and HABS, and a held-out set from NACC were used for model testing.

Extended data
| **Metric** | **$A\beta$ label** | **Meta-τ label** |
| --- | --- | --- |
| Accuracy | 0.68 | 0.82 |
| Balanced Accuracy | 0.68 | 0.76 |
| Precision | 0.62 | 0.52 |
| Sensitivity/Recall | 0.77 | 0.67 |
| Specificity | 0.59 | 0.85 |
| F1 score | 0.69 | 0.59 |
| MCC | 0.37 | 0.48 |
| AUC (ROC) | 0.79 | 0.84 |
| AUC (PR) | 0.78 | 0.60 |
| NPV | 0.75 | 0.92 |

(a)
| **Metric** | $\tau_{\text{med-temporal}}$ | $\tau_{\text{lat-temporal}}$ | $\tau_{\text{med-parietal}}$ | $\tau_{\text{lat-parietal}}$ | $\tau_{\text{frontal}}$ | $\tau_{\text{occipital}}$ |
| --- | --- | --- | --- | --- | --- | --- |
| Accuracy | 0.68 | 0.75 | 0.84 | 0.82 | 0.76 | 0.80 |
| Balanced Accuracy | 0.71 | 0.74 | 0.72 | 0.72 | 0.61 | 0.78 |
| Precision | 0.43 | 0.33 | 0.27 | 0.31 | 0.24 | 0.23 |
| Sensitivity/Recall | 0.75 | 0.72 | 0.58 | 0.60 | 0.42 | 0.76 |
| Specificity | 0.66 | 0.76 | 0.87 | 0.84 | 0.81 | 0.80 |
| F1 score | 0.55 | 0.45 | 0.37 | 0.40 | 0.31 | 0.36 |
| MCC | 0.36 | 0.36 | 0.32 | 0.33 | 0.18 | 0.34 |
| AUC (ROC) | 0.79 | 0.83 | 0.82 | 0.79 | 0.71 | 0.84 |
| AUC (PR) | 0.60 | 0.50 | 0.37 | 0.39 | 0.30 | 0.37 |
| NPV | 0.89 | 0.94 | 0.96 | 0.95 | 0.90 | 0.98 |

## References

1. Jack Jr, C. R., et al. NIA-AA research framework: toward a biological definition of Alzheimer’s disease. Alzheimer’s & dementia 14, 535–562 (2018).

2. Dubois, B., von Arnim, C. A., Burnie, N., Bozeat, S. & Cummings, J. Biomarkers in alzheimer’s disease: role in early and differential diagnosis and recognition of atypical variants. Alzheimer’s Research & Therapy 15, 175 (2023).

3. Jia, J. et al. Biomarker changes during 20 years preceding Alzheimer’s disease. New England Journal of Medicine 390, 712–722 (2024).

4. Jack Jr, C. R., et al. Revised criteria for diagnosis and staging of alzheimer’s disease: Alzheimer’s association workgroup. Alzheimer’s & Dementia (2024).

5. Rabinovici, G. D. et al. Association of amyloid positron emission tomography with subsequent change in clinical management among medicare beneficiaries with mild cognitive impairment or dementia. Jama 321, 1286–1294 (2019).

6. Sims, J. R. et al. Donanemab in early symptomatic alzheimer disease: the trailblazer-alz 2 randomized clinical trial. Jama 330, 512–527 (2023).

7. Van Dyck, C. H. et al. Lecanemab in early Alzheimer’s disease. New England Journal of Medicine 388, 9–21 (2023).

8. Sperling, R. A. et al. Trial of solanezumab in preclinical Alzheimer’s disease. New England Journal of Medicine 389, 1096–1107 (2023).

9. Cummings, J. L., Goldman, D. P., Simmons-Stern, N. R. & Ponton, E. The costs of developing treatments for Alzheimer’s disease: A retrospective exploration. Alzheimer’s & Dementia 18, 469–477 (2022). URL https://onlinelibrary.wiley.com/doi/abs/10.1002/alz.12450.

10. Groot, C. et al. Tau positron emission tomography for predicting dementia in individuals with mild cognitive impairment. JAMA neurology 81, 845–856 (2024).

11. Ossenkoppele, R., van der Kant, R. & Hansson, O. Tau biomarkers in Alzheimer’s disease: towards implementation in clinical practice and trials. The Lancet Neurology 21, 726–734 (2022).

12. Cummings, J. et al. Alzheimer’s disease drug development pipeline: 2023. Alzheimer’s & Dementia: Translational Research & Clinical Interventions 9, e12385 (2023).

13. Lantero Rodriguez, J., et al. Plasma p-tau181 accurately predicts Alzheimer’s disease pathology at least 8 years prior to post-mortem and improves the clinical characterisation of cognitive decline. Acta neuropathologica 140, 267–278 (2020).

14. Ashton, N. J. et al. Diagnostic accuracy of a plasma phosphorylated tau 217 immunoassay for Alzheimer disease pathology. JAMA neurology (2024).

15. Palmqvist, S. et al. Performance of fully automated plasma assays as screening tests for alzheimer disease–related beta-amyloid status. JAMA neurology 76, 1060–1069 (2019).

16. Shen, X.-N. et al. Plasma phosphorylated-tau181 as a predictive biomarker for Alzheimer’s amyloid, tau and fdg pet status. Translational psychiatry 11, 585 (2021).

17. Vogel, J. W. et al. Four distinct trajectories of tau deposition identified in Alzheimer’s disease. Nature medicine 27, 871–881 (2021).

18. Young, C. B. et al. Divergent cortical tau positron emission tomography patterns among patients with preclinical Alzheimer disease. JAMA neurology 79, 592–603 (2022).

19. Mielke, M. M. et al. Performance of plasma phosphorylated tau 181 and 217 in the community. Nature medicine 28, 1398–1405 (2022).

20. Blood biomarkers promise to revolutionize alzheimer’s diagnosis (2024). URL https://www.nature.com/articles/d42473-024-00030-5.

21. Mielke, M. M. & Fowler, N. R. Alzheimer disease blood biomarkers: considerations for population-level use. Nature Reviews Neurology 1–10 (2024).

22. Palmqvist, S., et al. Accurate risk estimation of β-amyloid positivity to identify prodromal Alzheimer’s disease: Cross-validation study of practical algorithms.Alzheimer’s & Dementia 15, 194– 204 (2019). URL https://onlinelibrary.wiley.com/doi/abs/10.1016/j.jalz.2018.08.014.

23. Tosun, D. et al. Detection of *β*-amyloid positivity in alzheimer’s disease neuroimaging initiative participants with demographics, cognition, mri and plasma biomarkers. Brain communications 3, fcab008 (2021).

24. Kang, S. H. et al. Machine learning for the prediction of amyloid positivity in amnestic mild cognitive impairment. Journal of Alzheimer’s Disease 80, 143–157 (2021).

25. Pekkala, T. et al. Detecting amyloid positivity in elderly with increased risk of cognitive decline. Frontiers in Aging Neuroscience 12, 228 (2020).

26. Shan, G., Bernick, C., Caldwell, J. Z. K. & Ritter, A. Machine learning methods to predict amyloid positivity using domain scores from cognitive tests. Scientific Reports 11, 4822 (2021). URL https://www.nature.com/articles/s41598-021-84101-4.

27. Petersen, K. K. et al. Predicting amyloid positivity in cognitively unimpaired older adults: A machine learning approach using a4 data. Neurology 98, e2425–e2435 (2022).

28. Wu, J. et al. Improved prediction of amyloid-*β* and tau burden using hippocampal surface multivariate morphometry statistics and sparse coding. Journal of Alzheimer’s Disease 91, 637–651 (2023).

29. Moradi, E. et al. Machine learning prediction of future amyloid beta positivity in amyloid-negative individuals. Alzheimer’s Research & Therapy 16, 46 (2024). URL https://alzres.biomedcentral.com/articles/10.1186/s13195-024-01127-0.

30. Karlsson, L. et al. Machine learning prediction of tau-pet in alzheimer’s disease using plasma, mri, and clinical data. Alzheimer’s & Dementia 21, e14600 (2025).

31. Villemagne, V. L. et al. CenTauR: Toward a universal scale and masks for standardizing tau imaging studies. Alzheimer’s & Dementia: Diagnosis, Assessment & Disease Monitoring 15, e12454 (2023). URL https://onlinelibrary.wiley.com/doi/abs/10.1002/dad2.12454.

32. McKee, A. C., Stein, T. D., Kiernan, P. T. & Alvarez, V. E. The neuropathology of chronic traumatic encephalopathy. Brain pathology 25, 350–364 (2015).

33. Braak, H. & Braak, E. Neuropathological stageing of Alzheimer-related changes. Acta neuropathologica 82, 239–259 (1991).

34. Xue, C. et al. AI-based differential diagnosis of dementia etiologies on multimodal data. Nature Medicine 1–13 (2024). URL https://www.nature.com/articles/s41591-024-03118-z.

35. Mackenzie, I. R. et al. Nomenclature and nosology for neuropathologic subtypes of frontotemporal lobar degeneration: an update. Acta neuropathologica 119, 1–4 (2010).

36. Tsai, R. M. et al. 18 f-flortaucipir (av-1451) tau pet in frontotemporal dementia syndromes. Alzheimer’s research & therapy 11, 1–18 (2019).

37. Stern, R. A. et al. Tau positron-emission tomography in former national football league players. New England journal of medicine 380, 1716–1725 (2019).

38. Ossenkoppele, R. et al. Amyloid and tau PET-positive cognitively unimpaired individuals are at high risk for future cognitive decline. Nature Medicine 28, 2381–2387 (2022). URL https://www.nature.com/articles/s41591-022-02049-x.

39. Insel, P. S. et al. Tau positron emission tomography in preclinical Alzheimer’s disease. Brain 146, 700– 711 (2022). URL https://www.ncbi.nlm.nih.gov/pmc/articles/PMC10169284/.

40. Ferreira, D., Nordberg, A. & Westman, E. Biological subtypes of Alzheimer disease: a systematic review and meta-analysis. Neurology 94, 436–448 (2020).

41. Sperling, R. A. et al. The a4 study: stopping ad before symptoms begin? Science translational medicine 6, 228fs13–228fs13 (2014).

42. Beekly, D. L., et al. The national Alzheimer’s coordinating center (nacc) database: an Alzheimer disease database. *Alzheimer Disease & Associated Disorders* 18, 270–277 (2004).

43. LaMontagne, P. J. et al. Oasis-3: longitudinal neuroimaging, clinical, and cognitive dataset for normal aging and Alzheimer’s disease. medrxiv 2019–12 (2019).

44. Ellis, K. A. et al. The australian imaging, biomarkers and lifestyle (aibl) study of aging: methodology and baseline characteristics of 1112 individuals recruited for a longitudinal study of alzheimer’s disease. International psychogeriatrics 21, 672–687 (2009).

45. Mahmood, S. S., Levy, D., Vasan, R. S. & Wang, T. J. The framingham heart study and the epidemiology of cardiovascular disease: a historical perspective. The lancet 383, 999–1008 (2014).

46. Petersen, R. C. et al. Alzheimer’s disease neuroimaging initiative (adni). Neurology 74, 201–209 (2010).

47. Dagley, A. et al. Harvard aging brain study: dataset and accessibility. Neuroimage 144, 255–258 (2017).

48. Klunk, W. E. et al. The Centiloid Project: standardizing quantitative amyloid plaque estimation by PET. Alzheimer’s & Dementia: The Journal of the Alzheimer’s Association 11, 1–15.e1–4 (2015).

49. Sanchez, J. S. et al. The cortical origin and initial spread of medial temporal tauopathy in Alzheimer’s disease assessed with positron emission tomography. Science translational medicine 13, eabc0655 (2021).

50. Young, C. B. et al. Speech patterns during memory recall relates to early tau burden across adulthood. Alzheimer’s & Dementia 20, 2552–2563 (2024).

51. Landau, S. M. et al. Positron emission tomography harmonization in the alzheimer’s disease neuroimaging initiative: A scalable and rigorous approach to multisite amyloid and tau quantification. Alzheimer’s & Dementia (2024).

52. Sperling, R. A. et al. The impact of ab and tau on prospective cognitive decline in older individuals. Ann. Neurol 85.

53. Orlhac, F. et al. A guide to combat harmonization of imaging biomarkers in multicenter studies. Journal of Nuclear Medicine 63, 172–179 (2022).

54. Henschel, L. et al. Fastsurfer-a fast and accurate deep learning based neuroimaging pipeline. NeuroImage 219, 117012 (2020).

55. Hatamizadeh, A. et al. Swin unetr: Swin transformers for semantic segmentation of brain tumors in mri images. In Crimi, A. & Bakas, S. (eds.) Brainlesion: Glioma, Multiple Sclerosis, Stroke and Traumatic Brain Injuries, 272–284 (Springer International Publishing, Cham, 2022).

56. Tang, Y. et al. Self-supervised pre-training of swin transformers for 3d medical image analysis. In 2022 IEEE/CVF Conference on Computer Vision and Pattern Recognition (CVPR), 20698–20708 (2022).

57. Lin, T.-Y., Goyal, P., Girshick, R., He, K. & Dollar, P. Focal loss for dense object detection (2018). URL https://arxiv.org/abs/1708.02002. 1708.02002.

58. Loshchilov, I. & Hutter, F. Decoupled weight decay regularization (2019). URL https://arxiv.org/abs/1711.05101.1711.05101.

59. Lundberg, S. A unified approach to interpreting model predictions. arXiv preprint arXiv:1705.07874 (2017).

60. Mitchell, R., Cooper, J., Frank, E. & Holmes, G. Sampling permutations for Shapley value estimation. Journal of Machine Learning Research 23, 1–46 (2022).

61. Dorogush, A. V., Ershov, V. & Gulin, A. Catboost: gradient boosting with categorical features support. arXiv preprint arXiv:1810.11363 (2018).

62. Cover, T. M. Elements of information theory (John Wiley & Sons, 1999).

63. Strehl, A. & Ghosh, J. Cluster ensembles—a knowledge reuse framework for combining multiple partitions. Journal of machine learning research 3, 583–617 (2002).

64. Danon, L., Diaz-Guilera, A., Duch, J. & Arenas, A. Comparing community structure identification. Journal of statistical mechanics: Theory and experiment 2005, P09008 (2005).

65. Blondel, V. D., Guillaume, J.-L., Lambiotte, R. & Lefebvre, E. Fast unfolding of communities in large networks. Journal of statistical mechanics: theory and experiment 2008, P10008 (2008).

66. Vinh, N. X., Epps, J. & Bailey, J. Information theoretic measures for clusterings comparison: is a correction for chance necessary? In Proceedings of the 26th annual international conference on machine learning, 1073–1080 (2009).

67. Romano, S., Vinh, N. X., Bailey, J. & Verspoor, K. Adjusting for chance clustering comparison measures. Journal of Machine Learning Research 17, 1–32 (2016).

68. Welch, W. J. Construction of permutation tests. Journal of the American Statistical Association 85, 693–698 (1990).

69. Alexander-Bloch, A. F. et al. On testing for spatial correspondence between maps of human brain structure and function. Neuroimage 178, 540–551 (2018).

