## Supplement for "AI-driven fusion of neurological work-up for assessment of biological Alzheimer’s disease"

### Supplementary material

| Category | Features |
|---|---|
| Patient History | Patient Sex; Hispanic/Latino ethnicity; Primary language; Years of education; Marital status; Type of residence; Subject's age at visit; Derived NIH race definitions; Indicator of mother with cognitive impairment; Indicator of father with cognitive impairment; Subject's height (inches); Subject's weight (lbs); Subject blood pressure (sitting), systolic; Subject blood pressure (sitting), diastolic; Subject resting heart rate (pulse) |
| Genetics | Number of APOE e4 alleles |
| Clinical Dementia Rating (CDR) | Memory; Orientation; Judgment and problem-solving; Community affairs; Home and hobbies; Personal care; CDR sum of boxes; Global CDR |
| Geriatric Depression Scale (GDS) | Are you basically satisfied with your life?; Have you dropped many of your activities and interests?; Do you feel that your life is empty?; Do you often get bored?; Are you in good spirits most of the time?; Are you afraid that something bad is going to happen to you?; Do you feel happy most of the time?; Do you often feel helpless?; Do you prefer to stay at home, rather than going out and doing new things?; Do you feel you have more problems with memory than most?; Do you think it is wonderful to be alive now?; Do you feel pretty worthless the way you are now?; Do you feel full of energy?; Do you feel that your situation is hopeless?; Do you think that most people are better off than you are?; Total GDS Score |
| Neuropsychological Battery | Orientation subscale score - Time; Orientation subscale score - Place; Total MMSE score (using D-L-R-O-W); Total number of story units recalled from this current test administration; Logical Memory IIA - Delayed - Total number of story units recalled; WAIS-R Digit Symbol |

**Table S1: Features available for A4.**

| Category | Features |
|---|---|
| Patient History | Patient Sex; Subject's age at visit; Smoked more than 100 cigarettes in life |
| Genetics | Number of APOE e4 alleles |
| Clinical Dementia Rating (CDR) | Global CDR |

| | |
|---|---|
| Neuropsychological Battery | Total MMSE score (using D-L-R-O-W); Total number of story units recalled from this current test administration; Logical Memory IIA - Delayed - Total number of story units recalled |

**Table S2: Features available for AIBL.**

| **Category** | **Features** |
|---|---|
| Patient History | Patient Sex; Hispanic/Latino ethnicity; Primary language; Years of education; Marital status; Is the subject left- or right-handed?; Subject's age at visit; Derived NIH race definitions; Smoked cigarettes in last 30 days; Subject's height (inches); Subject's weight (lbs); body mass index (BMI); Subject blood pressure (sitting), systolic; Subject blood pressure (sitting), diastolic |
| Medication | Reported current use of lipid lowering medication; Reported current use of a diabetes medication |
| Genetics | Number of APOE e4 alleles |
| Clinical Dementia Rating (CDR) | Global CDR |
| Neuropsychological Battery | Total MMSE score (using D-L-R-O-W); Total number of story units recalled from this current test administration; Logical Memory IIA - Delayed - Total number of story units recalled; Digit span forward length; Digit span backward length; Animals - Total number of animals named in 60 seconds; Trail Making Test Part A - Total number of seconds to complete; Trail Making Test Part b - Total number of seconds to complete; boston Naming Test (30) - Total score |
| Blood Tests | Plasma Tau 181 Measurements; Total Tau; AB42/AB40 Ratio |

**Table S3: Features available for FHS.**

| **Category** | **Features** |
|---|---|

| | |
|---|---|
| Patient History | Patient Sex; Hispanic/Latino ethnicity; Years of education; Marital status; Living situation; Level of independence; Type of residence; Is the subject left- or right-handed?; Subject's age at visit; Derived NIH race definitions; Indicator of mother with cognitive impairment; Indicator of father with cognitive impairment; Smoked cigarettes in last 30 days; Smoked more than 100 cigarettes in life; Total years smoked cigarettes; Average number of packs smoked per day; If the subject quit smoking, age at which he/she last smoked (i.e., quit); Heart attack/cardiac arrest; Atrial fibrillation; Angioplasty/endarterectomy/stent; Cardiac bypass procedure; Congestive heart failure; Other cardiovascular disease; Stroke; Transient ischemic attack (TIA); Parkinson's disease (PD); Other parkinsonian disorder; Seizures; Diabetes; Hypertension; Hypercholesterolemia; Vitamin b12 deficiency; Thyroid disease; Incontinence - urinary; Incontinence - bowel; Alcohol abuse - clinically significant occurring over a 12-month period manifested in one of the following areas: work, driving, legal, or social; Other abused substances - clinically significant impairment occurring over a 12-month period manifested in one of the following areas: work, driving, legal, or social; Other psychiatric disorder; Subject's height (inches); Subject's weight (lbs); Subject blood pressure (sitting), systolic; Subject blood pressure (sitting), diastolic; Subject resting heart rate (pulse); Without corrective lenses, is the subject's vision functionally normal?; Does the subject usually wear corrective lenses?; If the subject usually wears corrective lenses, is the subject's vision functionally normal with corrective lenses?; Without a hearing aid(s), is the subject's hearing functionally normal?; Does the subject usually wear a hearing aid(s)?; If the subject usually wears a hearing aid(s), is the subject's hearing functionally normal with a hearing aid(s)? |
| Genetics | Number of APOE e4 alleles |
| Clinical Dementia Rating (CDR) | Memory; Orientation; Judgment and problem-solving; Community affairs; Home and hobbies; Personal care; CDR sum of boxes; Global CDR |

| | |
|---|---|
| Geriatric Depression Scale (GDS) | Are you basically satisfied with your life?; Have you dropped many of your activities and interests?; Do you feel that your life is empty?; Do you often get bored?; Are you in good spirits most of the time?; Are you afraid that something bad is going to happen to you?; Do you feel happy most of the time?; Do you often feel helpless?; Do you prefer to stay at home, rather than going out and doing new things?; Do you feel you have more problems with memory than most?; Do you think it is wonderful to be alive now?; Do you feel pretty worthless the way you are now?; Do you feel full of energy?; Do you feel that your situation is hopeless?; Do you think that most people are better off than you are?; Total GDS Score |
| Neuropsychological Battery 1/2 | Total MMSE score (using D-L-R-O-W); Total number of story units recalled from this current test administration; Logical Memory IIA - Delayed - Total number of story units recalled; Total score for copy of benson figure; Total score for 10- to 15-minute delayed drawing of benson figure; Recognized original stimulus from among four options; Animals - Total number of animals named in 60 seconds; Vegetable - Total number of vegetables named in 60 seconds; Trail Making Test Part A - Total number of seconds to complete; Trail Making Test Part b - Total number of seconds to complete; WAIS-R Digit Symbol; Number of correct F-words generated in 1 minute; Number of F-words repeated in 1 minute; Number of non-F-words and rule violation errors in 1 minute; Number of correct L-words generated in 1 minute; Number of L-words repeated in 1 minute; Number of non-L-words and rule violation errors in 1 minute; Total number of correct F-words and L-words; Total number of F-word and L-word repetition errors; Total number of non-F/L-words and rule violation errors; MoCA Total Score - corrected for education; MoCA: Visuospatial/executive - Trails; MoCA: Visuospatial/executive - Cube; MoCA: Visuospatial/executive - Clock contour; MoCA: Visuospatial/executive - Clock numbers; MoCA: Visuospatial/executive - Clock hands; MoCA: Language - Naming; MoCA: Attention - Digits |

| | |
|---|---|
| Neuropsychological Battery (cont.) 2/2 | MoCA: Attention - Letter A; MoCA: Attention - Serial 7s; MoCA: Language - Repetition; MoCA: Language - Fluency; MoCA: Abstraction; MoCA: Delayed recall - No cue; MoCA: Orientation - Date; MoCA: Orientation - Month; MoCA: Orientation - Year; MoCA: Orientation - Day; MoCA: Orientation - Place; MoCA: Orientation - City; Craft Story 21 Recall (Immediate) - Total story units recalled, verbatim scoring; Craft Story 21 Recall (Immediate) - Total story units recalled, paraphrase scoring; Number Span Test: Forward - Number of correct trials; Number Span Test: Forward - Longest span forward; Number Span Test: backward - Number of correct trials; Number Span Test: backward - Longest span backward; Craft Story 21 Recall (Delayed) - Total story units recalled, verbatim scoring; Craft Story 21 Recall (Delayed) - Total story units recalled, paraphrase scoring; Craft Story 21 Recall (Delayed) - Delay time; Craft Story 21 Recall (Delayed) - Cue (boy) needed; Multilingual Naming Test (MINT) - Total score; Multilingual Naming Test (MINT) - Total correct without semantic cue; Multilingual Naming Test (MINT) - Semantic cues: Number given; Multilingual Naming Test (MINT) - Semantic cues: Number correct with cue; Multilingual Naming Test (MINT) - Phonemic cues: Number given; Multilingual Naming Test (MINT) - Phonemic cues: Number correct with cue |
| Neuropsychiatric Inventory (NPI) Questionnaire | Delusions in the last month; Hallucinations in the last month; Agitation or aggression in the last month; Depression or dysphoria in the last month; Anxiety in the last month; Elation or euphoria in the last month; Apathy or indifference in the last month; Disinhibition in the last month; Irritability or lability in the last month; Motor disturbance in the last month; Nighttime behaviors in the last month; Appetite and eating problems in the last month |

| | |
|---|---|
| Functional Activities Questionnaire (FAQ) | In the past four weeks, did the subject have any difficulty or need help with: Writing checks, paying bills, or balancing a checkbook; In the past four weeks, did the subject have any difficulty or need help with: Assembling tax records, business affairs, or other paper; In the past four weeks, did the subject have any difficulty or need help with: Shopping alone for clothes, household necessities, or groceries; In the past four weeks, did the subject have any difficulty or need help with: Playing a game of skill such as bridge or chess, working on a hobby; In the past four weeks, did the subject have any difficulty or need help with: Heating water, making a cup of coffee, turning off the stove; In the past four weeks, did the subject have any difficulty or need help with: Preparing a balanced meal; In the past four weeks, did the subject have any difficulty or need help with: Keeping track of current events; In the past four weeks, did the subject have any difficulty or need help with: Paying attention to and understanding a TV program, book, or magazine; In the past four weeks, did the subject have any difficulty or need help with: Remembering appointments, family occasions, holidays, medications; In the past four weeks, did the subject have any difficulty or need help with: Traveling out of the neighborhood, driving, or arranging to take public transportation |

**Table S4: Features available for OASIS.**

| Category | Features |
|---|---|

| | |
|---|---|
| Patient History 1/3 | Primary reason for coming to ADC; Principal referral source; Patient Sex; Hispanic/Latino ethnicity; Hispanic origins; Primary language; Years of education; Marital status; Living situation; Level of independence; Type of residence; Is the subject left- or right-handed?; Subject's age at visit; Derived NIH race definitions; Indicator of first-degree family member with cognitive impairment; Indicator of mother with cognitive impairment; Indicator of father with cognitive impairment; In this family, is there evidence of a dominantly inherited AD mutation?; In this family, is there evidence for an AD mutation (from list of specific mutations)?; Source of evidence for AD mutation; In this family, is there evidence for an FTLD mutation?; In this family, is there evidence for a mutation other than an AD or FTLD mutation?; Smoked cigarettes in last 30 days; Smoked more than 100 cigarettes in life; Total years smoked cigarettes; Average number of packs smoked per day; If the subject quit smoking, age at which he/she last smoked (i.e., quit); In the past three months, has the subject consumed any alcohol?; During the past three months, how often did the subject have at least one drink of any alcoholic beverage such as wine, beer, malt liquor, or spirits?; Heart attack/cardiac arrest; More than one heart attack/cardiac arrest?; Year of most recent heart attack |
| Patient History (cont.) 2/3 | Atrial fibrillation; Angioplasty/endarterectomy/stent; Cardiac bypass procedure; Pacemaker and/or defibrillator; Congestive heart failure; Angina; Heart valve replacement or repair; Other cardiovascular disease; Stroke; More than one stroke reported as of the Initial Visit; Most recently reported year of stroke as of the Initial Visit; Transient ischemic attack (TIA); More than one TIA reported as of the Initial Visit; Most recently reported year of TIA as of the Initial Visit; Parkinson's disease (PD); Year of PD diagnosis; Other parkinsonian disorder; Year of parkinsonian disorder diagnosis; Seizures; Traumatic brain injury (TBI); Traumatic brain injury (TBI) with brief loss of consciousness; TBI with extended loss of consciousness - 5 minutes of longer; TBI without loss of consciousness - as might result from military detonations or sports injury; Year of most recent TBI; Diabetes; If Recent/active or Remote/inactive diabetes, which type?; Hypertension; Hypercholesterolemia; Vitamin b12 deficiency; Thyroid disease; Arthritis; Type of arthritis |

| | |
|---|---|
| Patient History (cont.) 3/3 | Arthritis, region affected - upper extremity; Arthritis, region affected - lower extremity; Arthritis, region affected - spine; Region affected - unknown; Incontinence - urinary; Incontinence - bowel; Sleep apnea history reported at Initial Visit; REM sleep behavior disorder (RBD) history reported at Initial Visit; Hyposomnia/insomnia history reported at Initial Visit; Other sleep disorder history reported at Initial Visit; Alcohol abuse - clinically significant occurring over a 12-month period manifested in one of the following areas: work, driving, legal, or social; Other abused substances - clinically significant impairment occurring over a 12-month period manifested in one of the following areas: work, driving, legal, or social; Post-traumatic stress disorder (PTSD); bipolar disorder; Schizophrenia; Active depression in the last two years; Depression episodes more than two years ago; Anxiety; Obsessive-compulsive disorder (OCD); Developmental neuropsychiatric disorders (e.g., autism spectrum disorder [ASD], attention-deficit hyperactivity disorder [ADHD], dyslexia); Other psychiatric disorder; History of traumatic brain injury (TBI); Subject's height (inches); Subject's weight (lbs); body mass index (BMI); Subject blood pressure (sitting), systolic; Subject blood pressure (sitting), diastolic; Subject resting heart rate (pulse); Without corrective lenses, is the subject's vision functionally normal?; Does the subject usually wear corrective lenses?; If the subject usually wears corrective lenses, is the subject's vision functionally normal with corrective lenses?; Without a hearing aid(s), is the subject's hearing functionally normal?; Does the subject usually wear a hearing aid(s)?; If the subject usually wears a hearing aid(s), is the subject's hearing functionally normal with a hearing aid(s)? |

| | |
|---|---|
| Medication | Subject taking any medications; Total number of medications reported at each visit; Reported current use of any type of antihypertensive or blood pressure medication; Reported current use of an antihypertensive combination therapy; Reported current use of an angiotensin converting enzyme (ACE) inhibitor; Reported current use of an antiadrenergic agent; Reported current use of a beta-adrenergic blocking agent (beta-blocker); Reported current use of a calcium channel blocking agent; Reported current use of a diuretic; Reported current use of a vasodilator; Reported current use of an angiotensin II inhibitor; Reported current use of lipid lowering medication; Reported current use of nonsteroidal anti-inflammatory medication; Reported current use of an anticoagulant or antiplatelet agent; Reported current use of an antidepressant; Reported current use of an antipsychotic agent; Reported current use of an anxiolytic, sedative, or hypnotic agent; Reported current use of a FDA-approved medication for Alzheimer's disease symptoms; Reported current use of an antiparkinson agent; Reported current use of estrogen hormone therapy; Reported current use of estrogen + progestin hormone therapy; Reported current use of a diabetes medication |
| Genetics | Number of APOE e4 alleles |
| Clinical Dementia Rating (CDR) | Memory; Orientation; Judgment and problem-solving; Community affairs; Home and hobbies; Personal care; CDR sum of boxes; Global CDR |
| Geriatric Depression Scale (GDS) | Is the subject able to complete the GDS, based on the clinician's best judgment?; Are you basically satisfied with your life?; Have you dropped many of your activities and interests?; Do you feel that your life is empty?; Do you often get bored?; Are you in good spirits most of the time?; Are you afraid that something bad is going to happen to you?; Do you feel happy most of the time?; Do you often feel helpless?; Do you prefer to stay at home, rather than going out and doing new things?; Do you feel you have more problems with memory than most?; Do you think it is wonderful to be alive now?; Do you feel pretty worthless the way you are now?; Do you feel full of energy?; Do you feel that your situation is hopeless?; Do you think that most people are better off than you are?; Total GDS Score |

| | |
|---|---|
| Neuropsychological Battery 1/4 | Was any part of the MMSE completed?; Administration of the MMSE was:; Language of MMSE administration; Subject was unable to complete one or more sections due to visual impairment; Subject was unable to complete one or more sections due to hearing impairment; Orientation subscale score - Time; Orientation subscale score - Place; Intersecting pentagon subscale score; Total MMSE score (using D-L-R-O-W); The remainder of the battery was administered:; Language of test administration; Total score from the previous test administration; Total number of story units recalled from this current test administration; Logical Memory IIA - Delayed - Total number of story units recalled; Logical Memory IIA - Delayed - Time elapsed since Logical Memory IA - Immediate; Total score for copy of benson figure; Total score for 10- to 15-minute delayed drawing of benson figure; Recognized original stimulus from among four options; Digit span forward trials correct; Digit span forward length; Digit span backward trials correct; Digit span backward length; Animals - Total number of animals named in 60 seconds; Vegetable - Total number of vegetables named in 60 seconds; Trail Making Test Part A - Total number of seconds to complete; Part A - Number of commission errors; Part A - Number of correct lines; Trail Making Test Part b - Total number of seconds to complete; Part b - Number of commission errors; Part b - Number of correct lines |

| | |
|---|---|
| Neuropsychological Battery (cont.) 2/4 | WAIS-R Digit Symbol; boston Naming Test (30) - Total score; Number of correct F-words generated in 1 minute; Number of F-words repeated in 1 minute; Number of non-F-words and rule violation errors in 1 minute; Number of correct L-words generated in 1 minute; Number of L-words repeated in 1 minute; Number of non-L-words and rule violation errors in 1 minute; Total number of correct F-words and L-words; Total number of F-word and L-word repetition errors; Total number of non-F/L-words and rule violation errors; Per clinician, based on the neuropsychological examination, the subject's cognitive status is deemed; Modality of communication used to administer neuropsychological battery; Was any part of MoCA administered?; If no part of MoCA administered, reason code; Where was MoCA administered?; Language of MoCA administration; MoCA Total Score - corrected for education; MoCA: Visuospatial/executive - Trails; MoCA: Visuospatial/executive - Cube; MoCA: Visuospatial/executive - Clock contour; MoCA: Visuospatial/executive - Clock numbers; MoCA: Visuospatial/executive - Clock hands; MoCA: Language - Naming; MoCA: Memory - Registration (two trials); MoCA: Attention - Digits; MoCA: Attention - Letter A; MoCA: Attention - Serial 7s; MoCA: Language - Repetition; MoCA: Language - Fluency |

| | |
|---|---|
| Neuropsychological Battery (cont.) 3/4 | MoCA: Abstraction; MoCA: Delayed recall - No cue; MoCA: Delayed recall - Category cue; MoCA: Delayed recall - Recognition; MoCA: Orientation - Date; MoCA: Orientation - Month; MoCA: Orientation - Year; MoCA: Orientation - Day; MoCA: Orientation - Place; MoCA: Orientation - City; Craft Story 21 Recall (Immediate) - Total story units recalled, verbatim scoring; Craft Story 21 Recall (Immediate) - Total story units recalled, paraphrase scoring; Number Span Test: Forward - Number of correct trials; Number Span Test: Forward - Longest span forward; Number Span Test: backward - Number of correct trials; Number Span Test: backward - Longest span backward; Craft Story 21 Recall (Delayed) - Total story units recalled, verbatim scoring; Craft Story 21 Recall (Delayed) - Total story units recalled, paraphrase scoring; Craft Story 21 Recall (Delayed) - Delay time; Craft Story 21 Recall (Delayed) - Cue (boy) needed; Multilingual Naming Test (MINT) - Total score; Multilingual Naming Test (MINT) - Total correct without semantic cue; Multilingual Naming Test (MINT) - Semantic cues: Number given; Multilingual Naming Test (MINT) - Semantic cues: Number correct with cue; Multilingual Naming Test (MINT) - Phonemic cues: Number given; Multilingual Naming Test (MINT) - Phonemic cues: Number correct with cue; MoCA blind Total raw score - uncorrected; MoCA-blind Total Score - corrected for education; Rey Auditory Verbal Learning: Trial 1 total recall; Rey Auditory Verbal Learning: Trial 1 intrusions |

| | |
|---|---|
| Neuropsychological Battery (cont.) 4/4 | Rey Auditory Verbal Learning: Trial 2 total recall; Rey Auditory Verbal Learning: Trial 2 intrusions; Rey Auditory Verbal Learning: Trial 3 total recall; Rey Auditory Verbal Learning: Trial 3 intrusions; Rey Auditory Verbal Learning: Trial 4 total recall; Rey Auditory Verbal Learning: Trial 4 intrusions; Rey Auditory Verbal Learning: Trial 5 total recall; Rey Auditory Verbal Learning: Trial 5 intrusions; Rey Auditory Verbal Learning: Trial 6 total recall; Rey Auditory Verbal Learning: Trial 6 intrusions; Oral Trail Making Test - Part A: Total number of seconds to complete; Oral Trail Making Test - Part A: Number of commission errors; Oral Trail Making Test - Part A: Number of correct lines; Oral Trail Making Test Part b: Total number of seconds to complete; Oral Trail Making Test Part b: Number of commission errors; Oral Trail Making Test Part b: Number of correct lines; Rey Auditory Verbal Learning: total delayed recall; Rey Auditory Verbal Learning: delayed intrusions; Rey Auditory Verbal Learning: recognition total correct; Rey Auditory Verbal Learning: recognition total false positives; Verbal naming test: total correct without a cue; Verbal naming test: total correct with a phonemic cue; How valid do you think the participant's responses are?; What makes this participant's responses less valid? Hearing impairment; What makes this participant's responses less valid? Distractions; What makes this participant's responses less valid? Interruptions; What makes this participant's responses less valid? Lack of effort or disinterest; What makes this participant's responses less valid? Fatigue; What makes this participant's responses less valid? Emotional issues; What makes this participant's responses less valid? Unapproved assistance; What makes this participant's responses less valid? Other |
| Neuropsychiatric Inventory (NPI) Questionnaire | NPI-Q co-participant; Delusions in the last month; Hallucinations in the last month; Agitation or aggression in the last month; Depression or dysphoria in the last month; Anxiety in the last month; Elation or euphoria in the last month; Apathy or indifference in the last month; Disinhibition in the last month; Irritability or lability in the last month; Motor disturbance in the last month; Nighttime behaviors in the last month; Appetite and eating problems in the last month |

| | |
|---|---|
| Functional Activities Questionnaire (FAQ) | In the past four weeks, did the subject have any difficulty or need help with: Writing checks, paying bills, or balancing a checkbook; In the past four weeks, did the subject have any difficulty or need help with: Assembling tax records, business affairs, or other paper; In the past four weeks, did the subject have any difficulty or need help with: Shopping alone for clothes, household necessities, or groceries; In the past four weeks, did the subject have any difficulty or need help with: Playing a game of skill such as bridge or chess, working on a hobby; In the past four weeks, did the subject have any difficulty or need help with: Heating water, making a cup of coffee, turning off the stove; In the past four weeks, did the subject have any difficulty or need help with: Preparing a balanced meal; In the past four weeks, did the subject have any difficulty or need help with: Keeping track of current events; In the past four weeks, did the subject have any difficulty or need help with: Paying attention to and understanding a TV program, book, or magazine; In the past four weeks, did the subject have any difficulty or need help with: Remembering appointments, family occasions, holidays, medications; In the past four weeks, did the subject have any difficulty or need help with: Traveling out of the neighborhood, driving, or arranging to take public transportation |
| CSF Tests | Amyloid Beta in CSF; Abnormally elevated CSF Tau or pTau |

| | |
|---|---|
| Physical Exam 1/2 | Were there abnormal neurological exam findings?; Parkinsonian signs; Resting tremor - left arm; Resting tremor - right arm; Slowing of fine motor movements - left side; Slowing of fine motor movements - right side; Rigidity - left arm; Rigidity - right arm; bradykinesia; Parkinsonian gait disorder; Postural instability; Neurological sign considered by examiner to be most likely consistent with cerebrovascular disease; Cortical cognitive deficit (e.g., aphasia, apraxia, neglect); Focal or other neurological findings consistent with SIVD (subcortical ischemic vascular dementia); Motor (may include weakness of combination of face, arm, and leg; reflex changes, etc.) - left side; Motor (may include weakness of combination of face, arm, and leg; reflex changes, etc.) - right side; Cortical visual field loss - left side; Cortical visual field loss - right side; Somatosensory loss - left side; Somatosensory loss - right side; Higher cortical visual problem suggesting posterior cortical atrophy (e.g., prosopagnosia, simultagnosia, balint's syndrome) or apraxia of gaze |
| Physical Exam (cont.) 2/2 | Findings suggestive of progressive supranuclear palsy (PSP), corticobasal syndrome (CBS), or other related disorders; Eye movement changes consistent with PSP; Dysarthria consistent with PSP; Axial rigidity consistent with PSP; Gait disorder consistent with PSP; Apraxia of speech; Apraxia consistent with CBS - left side; Apraxia consistent with CBS - right side; Cortical sensory deficits consistent with CBS - left side; Cortical sensory deficits consistent with CBS - right side; Ataxia consistent with CBS - left side; Ataxia consistent with CBS - right side; Alien limb consistent with CBS - left side; Alien limb consistent with CBS - right side; Dystonia consistent with CBS, PSP, or related disorder - left side; Dystonia consistent with CBS, PSP, or related disorder - right side; Myoclonus consistent with CBS - left side; Myoclonus consistent with CBS - right side; Findings suggesting ALS (e.g., muscle wasting, fasciculations, upper motor and/or lower motor neuron signs); Normal pressure hydrocephalus - gait apraxia; Other findings (e.g., cerebella ataxia, chorea, myoclonus) |

**Table S5: Features available for NACC Training.**

| **Category** | **Features** |
|---|---|

| | |
|---|---|
| Patient History 1/3 | Patient Sex; Hispanic/Latino ethnicity; Primary language; Years of education; Marital status; Type of residence; Is the subject left- or right-handed?; Derived NIH race definitions; Subject's age at visit; Indicator of mother with cognitive impairment; Indicator of father with cognitive impairment; Subject's height (inches); Subject's weight (lbs); Subject blood pressure (sitting), systolic; Subject blood pressure (sitting), diastolic; Subject resting heart rate (pulse); Primary reason for coming to ADC; Principal referral source; Hispanic origins; Living situation; Level of independence; Indicator of first-degree family member with cognitive impairment; In this family, is there evidence of a dominantly inherited AD mutation?; In this family, is there evidence for an AD mutation (from list of specific mutations)?; Source of evidence for AD mutation; In this family, is there evidence for an FTLD mutation?; In this family, is there evidence for a mutation other than an AD or FTLD mutation?; Smoked cigarettes in last 30 days; Smoked more than 100 cigarettes in life; Total years smoked cigarettes; Average number of packs smoked per day; If the subject quit smoking, age at which he/she last smoked (i.e., quit) |

| | |
|---|---|
| Patient History (cont.) 2/3 | In the past three months, has the subject consumed any alcohol?; During the past three months, how often did the subject have at least one drink of any alcoholic beverage such as wine, beer, malt liquor, or spirits?; Heart attack/cardiac arrest; More than one heart attack/cardiac arrest?; Year of most recent heart attack; Atrial fibrillation; Angioplasty/endarterectomy/stent; Cardiac bypass procedure; Pacemaker and/or defibrillator; Congestive heart failure; Angina; Heart valve replacement or repair; Other cardiovascular disease; Stroke; More than one stroke reported as of the Initial Visit; Most recently reported year of stroke as of the Initial Visit; Transient ischemic attack (TIA); More than one TIA reported as of the Initial Visit; Most recently reported year of TIA as of the Initial Visit; Parkinson's disease (PD); Year of PD diagnosis; Other parkinsonian disorder; Year of parkinsonian disorder diagnosis; Seizures; Traumatic brain injury (TBI); Traumatic brain injury (TBI) with brief loss of consciousness; TBI with extended loss of consciousness - 5 minutes of longer; TBI without loss of consciousness - as might result from military detonations or sports injury; Year of most recent TBI; Diabetes; If Recent/active or Remote/inactive diabetes, which type?; Hypertension |

| | |
|---|---|
| Patient History (cont.) 3/3 | Hypercholesterolemia; Vitamin b12 deficiency; Thyroid disease; Arthritis; Type of arthritis; Arthritis, region affected - upper extremity; Arthritis, region affected - lower extremity; Arthritis, region affected - spine; Region affected - unknown; Incontinence - urinary; Incontinence - bowel; Sleep apnea history reported at Initial Visit; REM sleep behavior disorder (RBD) history reported at Initial Visit; Hyposomnia/insomnia history reported at Initial Visit; Other sleep disorder history reported at Initial Visit; Alcohol abuse - clinically significant occurring over a 12-month period manifested in one of the following areas: work, driving, legal, or social; Other abused substances - clinically significant impairment occurring over a 12-month period manifested in one of the following areas: work, driving, legal, or social; Post-traumatic stress disorder (PTSD); bipolar disorder; Schizophrenia; Active depression in the last two years; Depression episodes more than two years ago; Anxiety; Obsessive-compulsive disorder (OCD); Developmental neuropsychiatric disorders (e.g., autism spectrum disorder [ASD], attention-deficit hyperactivity disorder [ADHD], dyslexia); Other psychiatric disorder; History of traumatic brain injury (TBI); body mass index (BMI); Without corrective lenses, is the subject's vision functionally normal?; Does the subject usually wear corrective lenses?; If the subject usually wears corrective lenses, is the subject's vision functionally normal with corrective lenses?; Without a hearing aid(s), is the subject's hearing functionally normal?; Does the subject usually wear a hearing aid(s)?; If the subject usually wears a hearing aid(s), is the subject's hearing functionally normal with a hearing aid(s)? |

| | |
|---|---|
| Medication | Subject taking any medications; Total number of medications reported at each visit; Reported current use of any type of antihypertensive or blood pressure medication; Reported current use of an antihypertensive combination therapy; Reported current use of an angiotensin converting enzyme (ACE) inhibitor; Reported current use of an antiadrenergic agent; Reported current use of a beta-adrenergic blocking agent (beta-blocker); Reported current use of a calcium channel blocking agent; Reported current use of a diuretic; Reported current use of a vasodilator; Reported current use of an angiotensin II inhibitor; Reported current use of lipid lowering medication; Reported current use of nonsteroidal anti-inflammatory medication; Reported current use of an anticoagulant or antiplatelet agent; Reported current use of an antidepressant; Reported current use of an antipsychotic agent; Reported current use of an anxiolytic, sedative, or hypnotic agent; Reported current use of a FDA-approved medication for Alzheimer's disease symptoms; Reported current use of an antiparkinson agent; Reported current use of estrogen hormone therapy; Reported current use of estrogen + progestin hormone therapy; Reported current use of a diabetes medication |
| Genetics | Number of APOE e4 alleles |
| Clinical Dementia Rating (CDR) | Memory; Orientation; Judgment and problem-solving; Community affairs; Home and hobbies; Personal care; CDR sum of boxes; Global CDR |
| Geriatric Depression Scale (GDS) | Are you basically satisfied with your life?; Have you dropped many of your activities and interests?; Do you feel that your life is empty?; Do you often get bored?; Are you in good spirits most of the time?; Are you afraid that something bad is going to happen to you?; Do you feel happy most of the time?; Do you often feel helpless?; Do you prefer to stay at home, rather than going out and doing new things?; Do you feel you have more problems with memory than most?; Do you think it is wonderful to be alive now?; Do you feel pretty worthless the way you are now?; Do you feel full of energy?; Do you feel that your situation is hopeless?; Do you think that most people are better off than you are?; Total GDS Score; Is the subject able to complete the GDS, based on the clinician's best judgment? |

| | |
|---|---|
| Neuropsychological Battery 1/3 | Orientation subscale score - Time; Orientation subscale score - Place; Total MMSE score (using D-L-R-O-W); boston Naming Test (30) - Total score; Logical Memory IIA - Delayed - Total number of story units recalled; Total number of story units recalled from this current test administration; Animals - Total number of animals named in 60 seconds; Trail Making Test Part A - Total number of seconds to complete; Trail Making Test Part b - Total number of seconds to complete; Multilingual Naming Test (MINT) - Total score; MoCA Total Score - corrected for education; MoCA: Visuospatial/executive - Trails; MoCA: Visuospatial/executive - Cube; MoCA: Visuospatial/executive - Clock contour; MoCA: Visuospatial/executive - Clock numbers; MoCA: Visuospatial/executive - Clock hands; MoCA: Language - Naming; MoCA: Attention - Digits; MoCA: Attention - Letter A; MoCA: Attention - Serial 7s; MoCA: Language - Repetition; MoCA: Language - Fluency; MoCA: Abstraction; MoCA: Delayed recall - No cue; MoCA: Orientation - Date; MoCA: Orientation - Month; MoCA: Orientation - Year; MoCA: Orientation - Day; MoCA: Orientation - Place; MoCA: Orientation - City; Digit span forward trials correct; Digit span backward trials correct; Number of correct F-words generated in 1 minute; Was any part of the MMSE completed?; Administration of the MMSE was:; Language of MMSE administration; Subject was unable to complete one or more sections due to visual impairment |

| | |
|---|---|
| Neuropsychological Battery (cont.) 2/3 | Subject was unable to complete one or more sections due to hearing impairment; Intersecting pentagon subscale score; The remainder of the battery was administered:; Language of test administration; Logical Memory IIA - Delayed - Time elapsed since Logical Memory IA - Immediate; Total score for copy of benson figure; Total score for 10- to 15-minute delayed drawing of benson figure; Recognized original stimulus from among four options; Digit span forward length; Digit span backward length; Vegetable - Total number of vegetables named in 60 seconds; Part A - Number of commission errors; Part A - Number of correct lines; Part b - Number of commission errors; Part b - Number of correct lines; Number of F-words repeated in 1 minute; Number of non-F-words and rule violation errors in 1 minute; Number of correct L-words generated in 1 minute; Number of L-words repeated in 1 minute; Number of non-L-words and rule violation errors in 1 minute; Total number of correct F-words and L-words; Total number of F-word and L-word repetition errors; Total number of non-F/L-words and rule violation errors; Per clinician, based on the neuropsychological examination, the subject's cognitive status is deemed; Modality of communication used to administer neuropsychological battery; Was any part of MoCA administered?; If no part of MoCA administered, reason code; Where was MoCA administered?; Language of MoCA administration; MoCA: Memory - Registration (two trials); MoCA: Delayed recall - Category cue; MoCA: Delayed recall - Recognition; Craft Story 21 Recall (Immediate) - Total story units recalled, verbatim scoring; Craft Story 21 Recall (Immediate) - Total story units recalled, paraphrase scoring; Number Span Test: Forward - Number of correct trials; Number Span Test: Forward - Longest span forward; Number Span Test: backward - Number of correct trials |

| | |
|---|---|
| Neuropsychological Battery (cont.) 3/3 | Number Span Test: backward - Longest span backward; Craft Story 21 Recall (Delayed) - Total story units recalled, verbatim scoring; Craft Story 21 Recall (Delayed) - Total story units recalled, paraphrase scoring; Craft Story 21 Recall (Delayed) - Delay time; Craft Story 21 Recall (Delayed) - Cue (boy) needed; Multilingual Naming Test (MINT) - Total correct without semantic cue; Multilingual Naming Test (MINT) - Semantic cues: Number given; Multilingual Naming Test (MINT) - Semantic cues: Number correct with cue; Multilingual Naming Test (MINT) - Phonemic cues: Number given; Multilingual Naming Test (MINT) - Phonemic cues: Number correct with cue; MoCA blind Total raw score - uncorrected; MoCA-blind Total Score - corrected for education; Rey Auditory Verbal Learning: Trial 1 total recall; Rey Auditory Verbal Learning: Trial 1 intrusions; Rey Auditory Verbal Learning: Trial 2 total recall; Rey Auditory Verbal Learning: Trial 2 intrusions; Rey Auditory Verbal Learning: Trial 3 total recall; Rey Auditory Verbal Learning: Trial 3 intrusions; Rey Auditory Verbal Learning: Trial 4 total recall; Rey Auditory Verbal Learning: Trial 4 intrusions; Rey Auditory Verbal Learning: Trial 5 total recall; Rey Auditory Verbal Learning: Trial 5 intrusions; Rey Auditory Verbal Learning: Trial 6 total recall; Rey Auditory Verbal Learning: Trial 6 intrusions; Oral Trail Making Test - Part A: Total number of seconds to complete; Oral Trail Making Test - Part A: Number of commission errors; Oral Trail Making Test - Part A: Number of correct lines; Oral Trail Making Test Part b: Total number of seconds to complete; Oral Trail Making Test Part b: Number of commission errors; Oral Trail Making Test Part b: Number of correct lines; Rey Auditory Verbal Learning: total delayed recall; Rey Auditory Verbal Learning: delayed intrusions; Rey Auditory Verbal Learning: recognition total correct; Rey Auditory Verbal Learning: recognition total false positives; Verbal naming test: total correct without a cue; Verbal naming test: total correct with a phonemic cue; How valid do you think the participant's responses are? |

| | |
|---|---|
| Neuropsychiatric Inventory (NPI) Questionnaire | Delusions in the last month; Hallucinations in the last month; Agitation or aggression in the last month; Depression or dysphoria in the last month; Anxiety in the last month; Elation or euphoria in the last month; Apathy or indifference in the last month; Disinhibition in the last month; Irritability or lability in the last month; Motor disturbance in the last month; Nighttime behaviors in the last month; Appetite and eating problems in the last month; NPI-Q co-participant |
| Functional Activities Questionnaire (FAQ) | In the past four weeks, did the subject have any difficulty or need help with: Writing checks, paying bills, or balancing a checkbook; In the past four weeks, did the subject have any difficulty or need help with: Assembling tax records, business affairs, or other paper; In the past four weeks, did the subject have any difficulty or need help with: Shopping alone for clothes, household necessities, or groceries; In the past four weeks, did the subject have any difficulty or need help with: Playing a game of skill such as bridge or chess, working on a hobby; In the past four weeks, did the subject have any difficulty or need help with: Heating water, making a cup of coffee, turning off the stove; In the past four weeks, did the subject have any difficulty or need help with: Preparing a balanced meal; In the past four weeks, did the subject have any difficulty or need help with: Keeping track of current events; In the past four weeks, did the subject have any difficulty or need help with: Paying attention to and understanding a TV program, book, or magazine; In the past four weeks, did the subject have any difficulty or need help with: Remembering appointments, family occasions, holidays, medications; In the past four weeks, did the subject have any difficulty or need help with: Traveling out of the neighborhood, driving, or arranging to take public transportation |
| CSF Tests | Amyloid Beta in CSF; Abnormally elevated CSF Tau or pTau |

| | |
|---|---|
| Physical Exam 1/2 | Were there abnormal neurological exam findings?; Parkinsonian signs; Resting tremor - left arm; Resting tremor - right arm; Slowing of fine motor movements - left side; Slowing of fine motor movements - right side; Rigidity - left arm; Rigidity - right arm; bradykinesia; Parkinsonian gait disorder; Postural instability; Neurological sign considered by examiner to be most likely consistent with cerebrovascular disease; Cortical cognitive deficit (e.g., aphasia, apraxia, neglect); Focal or other neurological findings consistent with SIVD (subcortical ischemic vascular dementia); Motor (may include weakness of combination of face, arm, and leg; reflex changes, etc.) - left side; Motor (may include weakness of combination of face, arm, and leg; reflex changes, etc.) - right side; Cortical visual field loss - left side; Cortical visual field loss - right side; Somatosensory loss - left side; Somatosensory loss - right side; Higher cortical visual problem suggesting posterior cortical atrophy (e.g., prosopagnosia, simultagnosia, balint's syndrome) or apraxia of gaze |
| Physical Exam (cont.) 2/2 | Findings suggestive of progressive supranuclear palsy (PSP), corticobasal syndrome (CBS), or other related disorders; Eye movement changes consistent with PSP; Dysarthria consistent with PSP; Axial rigidity consistent with PSP; Gait disorder consistent with PSP; Apraxia of speech; Apraxia consistent with CBS - left side; Apraxia consistent with CBS - right side; Cortical sensory deficits consistent with CBS - left side; Cortical sensory deficits consistent with CBS - right side; Ataxia consistent with CBS - left side; Ataxia consistent with CBS - right side; Alien limb consistent with CBS - left side; Alien limb consistent with CBS - right side; Dystonia consistent with CBS, PSP, or related disorder - left side; Dystonia consistent with CBS, PSP, or related disorder - right side; Myoclonus consistent with CBS - left side; Myoclonus consistent with CBS - right side; Findings suggesting ALS (e.g., muscle wasting, fasciculations, upper motor and/or lower motor neuron signs); Normal pressure hydrocephalus - gait apraxia; Other findings (e.g., cerebella ataxia, chorea, myoclonus) |

**Table S6: Features available for NACC Testing.**

| Category | Features |
|---|---|

| | |
|---|---|
| Patient History | Patient Sex; Hispanic/Latino ethnicity; Years of education; Derived NIH race definitions; Subject's age at visit |
| Genetics | Number of APOE e4 alleles |
| Clinical Dementia Rating (CDR) | Memory; CDR sum of boxes; Global CDR |
| Geriatric Depression Scale (GDS) | Total GDS Score |
| Neuropsychological Battery | Total MMSE score (using D-L-R-O-W); boston Naming Test (30) - Total score; Logical Memory IIA - Delayed - Total number of story units recalled; Total number of story units recalled from this current test administration; Animals - Total number of animals named in 60 seconds; Trail Making Test Part A - Total number of seconds to complete; Trail Making Test Part b - Total number of seconds to complete; Digit span forward trials correct; Digit span backward trials correct; Number of correct F-words generated in 1 minute |

**Table S7: Features available for HABS.**

| **Category** | **Features** |
|---|---|
| Patient History | Patient Sex; Hispanic/Latino ethnicity; Primary language; Years of education; Marital status; Type of residence; Is the subject left- or right-handed?; Derived NIH race definitions; Subject's age at visit; Indicator of mother with cognitive impairment; Indicator of father with cognitive impairment; Subject's height (inches); Subject's weight (lbs); Subject blood pressure (sitting), systolic; Subject blood pressure (sitting), diastolic; Subject resting heart rate (pulse) |
| Genetics | Number of APOE e4 alleles |
| Clinical Dementia Rating (CDR) | Memory; Orientation; Judgment and problem-solving; Community affairs; Home and hobbies; Personal care; CDR sum of boxes; Global CDR |

| | |
|---|---|
| Geriatric Depression Scale (GDS) | Are you basically satisfied with your life?; Have you dropped many of your activities and interests?; Do you feel that your life is empty?; Do you often get bored?; Are you in good spirits most of the time?; Are you afraid that something bad is going to happen to you?; Do you feel happy most of the time?; Do you often feel helpless?; Do you prefer to stay at home, rather than going out and doing new things?; Do you feel you have more problems with memory than most?; Do you think it is wonderful to be alive now?; Do you feel pretty worthless the way you are now?; Do you feel full of energy?; Do you feel that your situation is hopeless?; Do you think that most people are better off than you are?; Total GDS Score |
| Neuropsychological Battery | Orientation subscale score - Time; Orientation subscale score - Place; Total MMSE score (using D-L-R-O-W); boston Naming Test (30) - Total score; Logical Memory IIA - Delayed - Total number of story units recalled; Total number of story units recalled from this current test administration; Animals - Total number of animals named in 60 seconds; Trail Making Test Part A - Total number of seconds to complete; Trail Making Test Part b - Total number of seconds to complete; Multilingual Naming Test (MINT) - Total score; MoCA Total Score - corrected for education; MoCA: Visuospatial/executive - Trails; MoCA: Visuospatial/executive - Cube; MoCA: Visuospatial/executive - Clock contour; MoCA: Visuospatial/executive - Clock numbers; MoCA: Visuospatial/executive - Clock hands; MoCA: Language - Naming; MoCA: Attention - Digits; MoCA: Attention - Letter A; MoCA: Attention - Serial 7s; MoCA: Language - Repetition; MoCA: Language - Fluency; MoCA: Abstraction; MoCA: Delayed recall - No cue; MoCA: Orientation - Date; MoCA: Orientation - Month; MoCA: Orientation - Year; MoCA: Orientation - Day; MoCA: Orientation - Place; MoCA: Orientation - City |
| Neuropsychiatric Inventory (NPI) Questionnaire | Delusions in the last month; Hallucinations in the last month; Agitation or aggression in the last month; Depression or dysphoria in the last month; Anxiety in the last month; Elation or euphoria in the last month; Apathy or indifference in the last month; Disinhibition in the last month; Irritability or lability in the last month; Motor disturbance in the last month; Nighttime behaviors in the last month; Appetite and eating problems in the last month |

| | |
|---|---|
| Functional Activities Questionnaire (FAQ) | In the past four weeks, did the subject have any difficulty or need help with: Writing checks, paying bills, or balancing a checkbook; In the past four weeks, did the subject have any difficulty or need help with: Assembling tax records, business affairs, or other paper; In the past four weeks, did the subject have any difficulty or need help with: Shopping alone for clothes, household necessities, or groceries; In the past four weeks, did the subject have any difficulty or need help with: Playing a game of skill such as bridge or chess, working on a hobby; In the past four weeks, did the subject have any difficulty or need help with: Heating water, making a cup of coffee, turning off the stove; In the past four weeks, did the subject have any difficulty or need help with: Preparing a balanced meal; In the past four weeks, did the subject have any difficulty or need help with: Keeping track of current events; In the past four weeks, did the subject have any difficulty or need help with: Paying attention to and understanding a TV program, book, or magazine; In the past four weeks, did the subject have any difficulty or need help with: Remembering appointments, family occasions, holidays, medications; In the past four weeks, did the subject have any difficulty or need help with: Traveling out of the neighborhood, driving, or arranging to take public transportation |
| Blood Tests | Plasma Tau 181 Measurements; AB42/AB40 Ratio |
| CSF Tests | Amyloid Beta in CSF; Abnormally elevated CSF Tau or pTau |

**Table S8: Features available for ADNI.**

| **Dataset** (Label) | **Age** (mean ± std) | **Male gender** (n, %) | **Education in years** (mean ± std) | **Race** (White; Black; Asian; American Indian; Pacific; Multi-race) | **CDR** (0.0, 0.5, 1.0, 2.0) | **MRIs available** (T1, T2*, FLAIR) |
|---|---|---|---|---|---|---|
| | | | **Training** | | | |
| **NACC training** [n = 4193] | | | | | | |
| Frontal Lobe Tau-Negative [n=283, 84.98%] | 71.16±8.10 | 139, 49.12% | 16.45±2.49 | 237; 15; 2; 12; 0; 15 | 160; 103; 18; 1 | 0, 0, 0 |
| Frontal Lobe Tau-Positive [n=50, 15.02%] | 70.44±8.53 | 22, 44.00% | 15.92±2.47 | 46; 2; 0; 0; 0; 2 | 4; 27; 12; 7 | 0, 0, 0 |
| Occipital Lobe Tau-Negative [n=275, 82.58%] | 71.09±8.09 | 141, 51.27% | 16.43±2.46 | 229; 15; 2; 12; 0; 15 | 155; 98; 17; 4 | 0, 0, 0 |
| Occipital Lobe Tau-Positive [n=58, 17.42%] | 70.86±8.58 | 20, 34.48% | 16.12±2.63 | 54; 2; 0; 0; 0; 2 | 9; 32; 13; 4 | 0, 0, 0 |
| Medial Temporal Lobe Tau-Negative [n=225, 67.57%] | 70.52±8.39 | 108, 48.00% | 16.35±2.51 | 183; 14; 1; 12; 0; 13 | 146; 68; 9; 1 | 0, 0, 0 |
| Medial Temporal Lobe Tau-Positive [n=108, 32.43%] | 72.16±7.58 | 53, 49.07% | 16.43±2.47 | 100; 3; 1; 0; 0; 4 | 18; 62; 21; 7 | 0, 0, 0 |
| Lateral Temporal Lobe Tau-Negative [n=264, 79.28%] | 70.98±8.21 | 129, 48.86% | 16.45±2.47 | 219; 16; 1; 12; 0; 14 | 157; 90; 15; 1 | 0, 0, 0 |
| Lateral Temporal Lobe Tau-Positive [n=69, 20.72%] | 71.32±8.00 | 32, 46.38% | 16.07±2.57 | 64; 1; 1; 0; 0; 3 | 7; 40; 15; 7 | 0, 0, 0 |
| Medial Parietal Lobe Tau-Negative [n=296, 88.89%] | 71.45±8.02 | 146, 49.32% | 16.44±2.46 | 248; 16; 2; 12; 0; 16 | 164; 110; 19; 2 | 0, 0, 0 |
| Medial Parietal Lobe Tau-Positive [n=37, 11.11%] | 67.86±8.66 | 15, 40.54% | 15.84±2.69 | 35; 1; 0; 0; 0; 1 | 0; 20; 11; 6 | 0, 0, 0 |
| Lateral Parietal Lobe Tau-Negative [n=277, 83.18%] | 71.38±8.03 | 138, 49.82% | 16.47±2.46 | 232; 15; 2; 11; 0; 15 | 159; 99; 17; 1 | 0, 0, 0 |
| Lateral Parietal Lobe Tau-Positive [n=56, 16.82%] | 69.43±8.68 | 23, 41.07% | 15.91±2.62 | 51; 2; 0; 1; 0; 2 | 5; 31; 13; 7 | 0, 0, 0 |
| p-values | 3.31e-01 | 6.71e-01 | 6.92e-01 | N.A. | 2.61e-66 | |
| **OASIS** [n = 962] | | | | | | |
| Frontal Lobe Tau-Negative [n=312, 83.87%] | 68.98±8.48 | 140, 44.87% | 16.35±2.37 | 276; 32; 2; 1; 0; 1 | 280; 27; 5; 0 | 311, 308, 308 |
| Frontal Lobe Tau-Positive [n=60, 16.13%] | 72.99±6.78 | 19, 31.67% | 16.15±2.23 | 49; 11; 0; 0; 0; 0 | 38; 12; 8; 2 | 60, 60, 60 |
| Occipital Lobe Tau-Negative [n=231, 62.10%] | 68.37±8.64 | 119, 51.52% | 16.53±2.33 | 204; 24; 2; 1; 0; 0 | 210; 18; 2; 1 | 230, 230, 229 |
| Occipital Lobe Tau-Positive [n=141, 37.90%] | 71.69±7.44 | 40, 28.37% | 15.96±2.33 | 121; 19; 0; 0; 0; 1 | 108; 21; 11; 1 | 141, 138, 139 |
| Medial Temporal Lobe Tau-Negative [n=300, 80.65%] | 68.81±8.41 | 133, 44.33% | 16.40±2.29 | 261; 36; 2; 1; 0; 0 | 281; 18; 1; 0 | 299, 298, 297 |
| Medial Temporal Lobe Tau-Positive [n=72, 19.35%] | 73.02±7.24 | 26, 36.11% | 15.99±2.53 | 64; 7; 0; 0; 0; 1 | 37; 21; 12; 2 | 72, 70, 71 |
| Lateral Temporal Lobe Tau-Negative [n=285, 76.61%] | 68.70±8.65 | 131, 45.96% | 16.39±2.35 | 252; 30; 2; 1; 0; 0 | 263; 20; 2; 0 | 284, 281, 281 |
| Lateral Temporal Lobe Tau-Positive [n=87, 23.39%] | 72.66±6.48 | 28, 32.18% | 16.07±2.31 | 73; 13; 0; 0; 0; 1 | 55; 19; 11; 2 | 87, 87, 87 |
| Medial Parietal Lobe Tau-Negative [n=294, 79.03%] | 68.67±8.55 | 125, 42.52% | 16.38±2.29 | 256; 34; 2; 1; 0; 1 | 271; 21; 2; 0 | 293, 290, 290 |
| Medial Parietal Lobe Tau-Positive [n=78, 20.97%] | 73.22±6.45 | 34, 43.59% | 16.06±2.53 | 69; 9; 0; 0; 0; 0 | 47; 18; 11; 2 | 78, 78, 78 |
| Lateral Parietal Lobe Tau-Negative [n=266, 71.51%] | 68.87±8.31 | 126, 47.37% | 16.41±2.32 | 234; 29; 2; 1; 0; 0 | 245; 19; 2; 0 | 265, 264, 263 |
| Lateral Parietal Lobe Tau-Positive [n=106, 28.49%] | 71.52±8.18 | 33, 31.13% | 16.08±2.39 | 91; 14; 0; 0; 0; 1 | 73; 20; 11; 2 | 106, 104, 105 |
| p-values | 4.93e-13 | 8.71e-05 | 4.27e-01 | N.A. | 1.29e-46 | |
| **A4** [n = 4475] | | | | | | |
| Frontal Lobe Tau-Negative [n=441, 99.32%] | 71.85±4.86 | 190, 43.08% | 16.21±2.84 | 404; 10; 20; 1; 0; 4 | 441; 0; 0; 0 | 441, 441, 80 |
| Frontal Lobe Tau-Positive [n=3, 0.68%] | 70.70±4.24 | 0, 0.00% | 18.67±1.15 | 3; 0; 0; 0; 0; 0 | 3; 0; 0; 0 | 3, 3, 0 |
| Occipital Lobe Tau-Negative [n=437, 98.42%] | 71.89±4.87 | 188, 43.02% | 16.24±2.84 | 400; 10; 20; 1; 0; 4 | 437; 0; 0; 0 | 437, 437, 79 |
| Occipital Lobe Tau-Positive [n=7, 1.58%] | 68.94±2.10 | 2, 28.57% | 15.57±2.82 | 7; 0; 0; 0; 0; 0 | 7; 0; 0; 0 | 7, 7, 1 |
| Medial Temporal Lobe Tau-Negative [n=382, 86.04%] | 71.66±4.85 | 165, 43.19% | 16.16±2.90 | 351; 8; 17; 1; 0; 3 | 382; 0; 0; 0 | 382, 382, 69 |
| Medial Temporal Lobe Tau-Positive [n=62, 13.96%] | 72.99±4.70 | 25, 40.32% | 16.68±2.37 | 56; 2; 3; 0; 0; 1 | 62; 0; 0; 0 | 62, 62, 11 |
| Lateral Temporal Lobe Tau-Negative [n=427, 96.17%] | 71.80±4.81 | 184, 43.09% | 16.22±2.86 | 391; 9; 20; 1; 0; 4 | 427; 0; 0; 0 | 427, 427, 79 |
| Lateral Temporal Lobe Tau-Positive [n=17, 3.83%] | 72.88±5.85 | 6, 35.29% | 16.35±2.26 | 16; 1; 0; 0; 0; 0 | 17; 0; 0; 0 | 17, 17, 1 |
| Medial Parietal Lobe Tau-Negative [n=439, 98.87%] | 71.88±4.86 | 189, 43.05% | 16.22±2.83 | 402; 10; 20; 1; 0; 4 | 439; 0; 0; 0 | 439, 439, 79 |
| Medial Parietal Lobe Tau-Positive [n=5, 1.13%] | 68.69±2.85 | 1, 20.00% | 17.00±3.32 | 5; 0; 0; 0; 0; 0 | 5; 0; 0; 0 | 5, 5, 1 |
| Lateral Parietal Lobe Tau-Negative [n=436, 98.20%] | 71.92±4.84 | 189, 43.35% | 16.22±2.84 | 399; 10; 20; 1; 0; 4 | 436; 0; 0; 0 | 436, 436, 79 |
| Lateral Parietal Lobe Tau-Positive [n=8, 1.80%] | 67.50±3.33 | 1, 12.50% | 16.75±2.87 | 8; 0; 0; 0; 0; 0 | 8; 0; 0; 0 | 8, 8, 1 |
| p-values | 1.33e-01 | 7.52e-01 | 9.26e-01 | N.A. | N.A. | |
| **FHS** [n = 255] | | | | | | |
| Frontal Lobe Tau-Negative [n=216, 100.00%] | 54.82±8.46 | 112, 51.85% | 15.45±2.05 | 216; 0; 0; 0; 0; 0 | 216; 0; 0; 0 | 216, 0, 0 |
| Occipital Lobe Tau-Negative [n=216, 100.00%] | 54.82±8.46 | 112, 51.85% | 15.45±2.05 | 216; 0; 0; 0; 0; 0 | 216; 0; 0; 0 | 216, 0, 0 |
| Medial Temporal Lobe Tau-Negative [n=216, 100.00%] | 54.82±8.46 | 112, 51.85% | 15.45±2.05 | 216; 0; 0; 0; 0; 0 | 216; 0; 0; 0 | 216, 0, 0 |
| Lateral Temporal Lobe Tau-Negative [n=216, 100.00%] | 54.82±8.46 | 112, 51.85% | 15.45±2.05 | 216; 0; 0; 0; 0; 0 | 216; 0; 0; 0 | 216, 0, 0 |
| Medial Parietal Lobe Tau-Negative [n=216, 100.00%] | 54.82±8.46 | 112, 51.85% | 15.45±2.05 | 216; 0; 0; 0; 0; 0 | 216; 0; 0; 0 | 216, 0, 0 |
| Lateral Parietal Lobe Tau-Negative [n=216, 100.00%] | 54.82±8.46 | 112, 51.85% | 15.45±2.05 | 216; 0; 0; 0; 0; 0 | 216; 0; 0; 0 | 216, 0, 0 |
| p-values | N.A. | 1.00e+00 | 1.00e+00 | N.A. | N.A. | |
| | | | **Testing** | | | |
| **ADNI** [n = 1404] | | | | | | |
| Frontal Lobe Tau-Negative [n=576, 92.01%] | 73.42±7.69 | 266, 46.18% | 16.42±2.45 | 489; 56; 15; 2; 0; 10 | 348; 191; 33; 3 | 576, 267, 306 |
| Frontal Lobe Tau-Positive [n=50, 7.99%] | 71.91±7.82 | 25, 50.00% | 15.80±2.03 | 43; 6; 0; 0; 0; 0 | 5; 32; 11; 2 | 50, 26, 21 |
| Occipital Lobe Tau-Negative [n=579, 92.49%] | 73.33±7.70 | 270, 46.63% | 16.44±2.43 | 490; 57; 15; 2; 0; 10 | 348; 198; 31; 2 | 579, 266, 306 |
| Occipital Lobe Tau-Positive [n=47, 7.51%] | 72.95±7.76 | 21, 44.68% | 15.53±2.29 | 42; 5; 0; 0; 0; 0 | 5; 25; 13; 3 | 47, 27, 21 |
| Medial Temporal Lobe Tau-Negative [n=472, 75.40%] | 72.71±7.57 | 219, 46.40% | 16.48±2.42 | 400; 48; 12; 2; 0; 7 | 321; 134; 15; 2 | 472, 224, 253 |
| Medial Temporal Lobe Tau-Positive [n=154, 24.60%] | 75.11±7.85 | 72, 46.75% | 16.03±2.42 | 132; 14; 3; 0; 0; 3 | 32; 89; 29; 3 | 154, 69, 74 |
| Lateral Temporal Lobe Tau-Negative [n=531, 84.82%] | 73.09±7.63 | 243, 45.76% | 16.47±2.43 | 446; 56; 14; 2; 0; 9 | 339; 171; 20; 1 | 531, 243, 282 |
| Lateral Temporal Lobe Tau-Positive [n=95, 15.18%] | 74.51±8.05 | 48, 50.53% | 15.83±2.36 | 86; 6; 1; 0; 0; 1 | 14; 52; 24; 4 | 95, 50, 45 |
| Medial Parietal Lobe Tau-Negative [n=588, 93.93%] | 73.50±7.66 | 276, 46.94% | 16.42±2.43 | 496; 60; 15; 2; 0; 10 | 349; 204; 30; 4 | 588, 272, 310 |
| Medial Parietal Lobe Tau-Positive [n=38, 6.07%] | 70.27±7.81 | 15, 39.47% | 15.66±2.29 | 36; 2; 0; 0; 0; 0 | 4; 19; 14; 1 | 38, 21, 17 |
| Lateral Parietal Lobe Tau-Negative [n=570, 91.05%] | 73.42±7.71 | 267, 46.84% | 16.45±2.43 | 485; 56; 14; 2; 0; 8 | 345; 194; 28; 3 | 570, 265, 304 |
| Lateral Parietal Lobe Tau-Positive [n=56, 8.95%] | 72.11±7.61 | 24, 42.86% | 15.57±2.26 | 47; 6; 1; 0; 0; 2 | 8; 29; 16; 2 | 56, 28, 23 |
| p-values | 1.39e-02 | 9.98e-01 | 2.41e-03 | N.A. | 1.49e-86 | |
| **HABS** [n = 282] | | | | | | |
| Frontal Lobe Tau-Negative [n=171, 98.84%] | 77.96±6.12 | 69, 40.35% | 16.09±3.08 | 148; 21; 1; 0; 0; 0 | 148; 20; 0; 0 | 171, 0, 170 |
| Frontal Lobe Tau-Positive [n=2, 1.16%] | 75.88±6.89 | 1, 50.00% | 16.00±0.00 | 2; 0; 0; 0; 0; 0 | 2; 0; 0; 0 | 2, 0, 2 |
| Occipital Lobe Tau-Negative [n=171, 98.84%] | 77.90±6.13 | 70, 40.94% | 16.10±3.06 | 149; 20; 1; 0; 0; 0 | 149; 19; 0; 0 | 171, 0, 170 |
| Occipital Lobe Tau-Positive [n=2, 1.16%] | 80.75±4.60 | 0, 0.00% | 15.00±4.24 | 1; 1; 0; 0; 0; 0 | 1; 1; 0; 0 | 2, 0, 2 |
| Medial Temporal Lobe Tau-Negative [n=141, 81.50%] | 77.46±6.19 | 59, 41.84% | 16.08±3.15 | 126; 14; 1; 0; 0; 0 | 125; 13; 0; 0 | 141, 0, 140 |
| Medial Temporal Lobe Tau-Positive [n=32, 18.50%] | 80.01±5.40 | 11, 34.38% | 16.12±2.67 | 24; 7; 0; 0; 0; 0 | 25; 7; 0; 0 | 32, 0, 32 |
| Lateral Temporal Lobe Tau-Negative [n=169, 97.69%] | 77.93±6.14 | 69, 40.83% | 16.04±3.07 | 148; 19; 1; 0; 0; 0 | 148; 18; 0; 0 | 169, 0, 168 |
| Lateral Temporal Lobe Tau-Positive [n=4, 2.31%] | 78.31±5.44 | 1, 25.00% | 18.00±1.63 | 2; 2; 0; 0; 0; 0 | 2; 2; 0; 0 | 4, 0, 4 |
| Medial Parietal Lobe Tau-Negative [n=173, 100.00%] | 77.93±6.11 | 70, 40.46% | 16.09±3.06 | 150; 21; 1; 0; 0; 0 | 150; 20; 0; 0 | 173, 0, 172 |
| Lateral Parietal Lobe Tau-Negative [n=173, 100.00%] | 77.93±6.11 | 70, 40.46% | 16.09±3.06 | 150; 21; 1; 0; 0; 0 | 150; 20; 0; 0 | 173, 0, 172 |
| p-values | 8.17e-01 | 9.82e-01 | 9.93e-01 | N.A. | N.A. | |

**Table S9: Study population characteristics broken down by regional tau label status** Demographic and clinical attributes for the training and testing cohorts used in the regional tau model. "N.A." entries in the table signify scenarios when statistical tests could not be concluded and when data were not available.

| Metric | $A\beta$ label | Meta-$\tau$ |
|---|---|---|
| Accuracy | 0.77 | 0.80 |
| Balanced Accuracy | 0.76 | 0.83 |
| Precision | 0.80 | 0.65 |
| Sensitivity/Recall | 0.80 | 0.94 |
| Specificity | 0.72 | 0.72 |
| F1 score | 0.80 | 0.77 |
| MCC | 0.52 | 0.63 |
| AUC (ROC) | 0.84 | 0.93 |
| AUC (PR) | 0.84 | 0.85 |
| NPV | 0.72 | 0.95 |

**Table S10: Performance metrics for amyloid and tau labels for the NACC test set.**

| Metric | $A\beta$ label | Meta-$\tau$ label |
|---|---|---|
| Accuracy | 0.81 | 0.86 |
| Balanced Accuracy | 0.79 | 0.80 |
| Precision | 0.85 | 0.48 |
| Sensitivity/Recall | 0.69 | 0.72 |
| Specificity | 0.90 | 0.88 |
| F1 score | 0.76 | 0.57 |
| MCC | 0.61 | 0.51 |
| AUC (ROC) | 0.87 | 0.88 |
| AUC (PR) | 0.86 | 0.57 |
| NPV | 0.78 | 0.95 |

**(a)**

| Metric | $\tau_{\text{medtemp}}$ | $\tau_{\text{lattemp}}$ | $\tau_{\text{medpar}}$ | $\tau_{\text{latpar}}$ | $\tau_{\text{front}}$ | $\tau_{\text{occ}}$ |
|---|---|---|---|---|---|---|
| Accuracy | 0.78 | 0.85 | 0.89 | 0.86 | 0.89 | 0.72 |
| Balanced Accuracy | 0.68 | 0.78 | 0.78 | 0.77 | 0.65 | 0.77 |
| Precision | 0.53 | 0.43 | 0.25 | 0.28 | 0.26 | 0.16 |
| Sensitivity/Recall | 0.51 | 0.70 | 0.66 | 0.68 | 0.38 | 0.82 |
| Specificity | 0.86 | 0.87 | 0.90 | 0.87 | 0.92 | 0.72 |
| F1 score | 0.52 | 0.53 | 0.36 | 0.40 | 0.31 | 0.26 |
| MCC | 0.37 | 0.47 | 0.36 | 0.37 | 0.26 | 0.27 |
| AUC (ROC) | 0.77 | 0.86 | 0.88 | 0.86 | 0.81 | 0.86 |
| AUC (PR) | 0.55 | 0.57 | 0.37 | 0.40 | 0.27 | 0.33 |
| NPV | 0.85 | 0.95 | 0.98 | 0.97 | 0.96 | 0.98 |

**(b)**

**Table S11: Overall performance metrics for all labels on external testing cohorts (ADNI and HABS).** Subtable (a) shows the performance metrics for $A\beta$ and meta-$\tau$ labels and Subtable (b) details the metrics on the regional $\tau$ labels.

| **Metric** | $A\beta$ **label** | **Meta-$\tau$ label** |
|---|---|---|
| Accuracy | 0.73 | 0.86 |
| Balanced Accuracy | 0.74 | 0.72 |
| Precision | 0.65 | 0.44 |
| Sensitivity/Recall | 0.89 | 0.52 |
| Specificity | 0.60 | 0.91 |
| F1 score | 0.75 | 0.48 |
| MCC | 0.50 | 0.40 |
| AUC (ROC) | 0.85 | 0.86 |
| AUC (PR) | 0.85 | 0.41 |

**(a)**

| **Metric** | $\tau_{\textbf{medtemp}}$ | $\tau_{\textbf{lattemp}}$ | $\tau_{\textbf{medpar}}$ | $\tau_{\textbf{latpar}}$ | $\tau_{\textbf{front}}$ | $\tau_{\textbf{occ}}$ |
|---|---|---|---|---|---|---|
| Accuracy | 0.80 | 0.88 | 0.93 | 0.91 | 0.94 | 0.94 |
| Balanced Accuracy | 0.69 | 0.71 | 0.71 | 0.72 | 0.59 | 0.62 |
| Precision | 0.58 | 0.52 | 0.34 | 0.41 | 0.53 | 0.46 |
| Sensitivity/Recall | 0.50 | 0.49 | 0.46 | 0.49 | 0.19 | 0.26 |
| Specificity | 0.89 | 0.93 | 0.95 | 0.94 | 0.99 | 0.98 |
| F1 score | 0.54 | 0.50 | 0.39 | 0.44 | 0.28 | 0.33 |
| MCC | 0.41 | 0.43 | 0.36 | 0.40 | 0.29 | 0.32 |
| AUC (ROC) | 0.77 | 0.82 | 0.88 | 0.81 | 0.74 | 0.77 |
| AUC (PR) | 0.58 | 0.50 | 0.35 | 0.43 | 0.30 | 0.36 |

**(b)**

**Table S12: Performance metrics for all labels with Catboost on the combined ADNI, HABS, and NACC* test set.** Table (a) shows Catboost's performance metrics for $A\beta$ and meta-$\tau$ labels. Table (b) details performance metrics for regional $\tau$ labels.

| Groups | Measure | Kruskal H | p-value |
|---|---|---|---|
| Meta-$\tau$ Tertiles | $P(A\beta)$ | 87.31 | $1.10 \times 10^{-19}$ |
| | CL | 65.22 | $6.86 \times 10^{-15}$ |
| CL Tertiles | $P(\tau)$ | 35.94 | $1.57 \times 10^{-8}$ |
| | Meta-$\tau$ SUVR | 50.93 | $8.73 \times 10^{-12}$ |

**(a)**

| Measure | Low vs Medium $\tau$ | Medium vs High $\tau$ | Low vs High $\tau$ |
|---|---|---|---|
| $P(A\beta)$ | $7.37 \times 10^{-2}$ | $2.89 \times 10^{-12}$ | $2.74 \times 10^{-18}$ |
| CL | $5.88 \times 10^{-2}$ | $7.48 \times 10^{-9}$ | $2.73 \times 10^{-14}$ |
| **Measure** | **Low vs Medium CL** | **Medium vs High CL** | **Low vs High CL** |
| $P(\tau)$ | $3.41 \times 10^{-3}$ | $4.45 \times 10^{-3}$ | $6.20 \times 10^{-9}$ |
| Meta-$\tau$ SUVR | $4.51 \times 10^{-4}$ | $4.51 \times 10^{-4}$ | $2.88 \times 10^{-12}$ |

**(b)**

**Table S13: Statistical analysis comparing low, medium, and high PET groups using Kruskal-Wallis tests followed by post hoc Dunn's tests with FDR correction on the combined ADNI and HABS external test cohorts.** **(a)** Kruskal-Wallis H test results comparing model predicted probabilities against PET-estimated tertiles in $A\beta+$ cases. Model predicted probabilities for amyloid, $P(A\beta)$, were compared across meta-$\tau$ PET tertile groups, and a similar analysis was run comparing centiloid scores across the $\tau$ tertiles. Conversely, model predicted probabilities for tau, $P(\tau)$, and meta-temporal $\tau$ SUVR, were compared across centiloid (CL) tertiles. **(b)** Post-Hoc Dunn's test p-values after FDR correction, comparing model predicted probabilities and PET derived measures across low, medium and high PET groups. $P(A\beta)$ and centiloids, CL, were each compared across $\tau$ PET tertile pairs, with significant results found between the medium vs high, and low vs high groups in both analyses. $P(\tau)$ and meta-$\tau$ SUVRs were each compared across tertile pairs, and we found significant differences across all pairs in both analyses. Refer to Figure 3 for the graphical representation of the data

| Comparison | Test | Coefficient | P-value |
|---|---|---|---|
| $P(A\beta)$ vs CL | Spearman's | 0.68 | $9.15 \times 10^{-188}$ |
| | Pearson's | 0.69 | $9.29 \times 10^{-194}$ |
| $P(\tau)$ vs meta-$\tau$ SUVR | Spearman's | 0.43 | $1.42 \times 10^{-29}$ |
| | Pearson's | 0.51 | $1.53 \times 10^{-42}$ |

**Table S14: Correlation coefficients and significance levels for model predicted probabilities against true biomarker values.** Correlation tests indicate significant associations between model predicted probabilities for both $A\beta$ and $\tau$ against continuous centiloid values (CL) and SUVRs, respectively. Refer to Figure 4a for the graphical representation of the data

| Measure | Kruskal H | p-value |
|---|---|---|
| $P(A\beta)$ | 188.06 | $1.38 \times 10^{-39}$ |
| $P(\tau)$ | 51.22 | $2.01 \times 10^{-10}$ |

**(a)**

| Measure | 5 vs 4 | 4 vs 3 | 3 vs 2 | 2 vs 1 | 5 vs 3 |
|---|---|---|---|---|---|
| $P(A\beta)$ | $6.14 \times 10^{-12}$ | $2.44 \times 10^{-2}$ | $1.03 \times 10^{-1}$ | $7.70 \times 10^{-3}$ | $6.76 \times 10^{-12}$ |
| $P(\tau)$ | $4.80 \times 10^{-4}$ | $1.80 \times 10^{-1}$ | $6.79 \times 10^{-1}$ | $9.58 \times 10^{-2}$ | $5.19 \times 10^{-4}$ |
| **Measure** | **5 vs 2** | **5 vs 1** | **4 vs 2** | **4 vs 1** | **3 vs 1** |
| $P(A\beta)$ | $6.57 \times 10^{-13}$ | $7.47 \times 10^{-20}$ | $2.94 \times 10^{-4}$ | $4.63 \times 10^{-10}$ | $1.91 \times 10^{-5}$ |
| $P(\tau)$ | $4.80 \times 10^{-4}$ | $6.05 \times 10^{-5}$ | $9.58 \times 10^{-2}$ | $2.70 \times 10^{-3}$ | $4.99 \times 10^{-2}$ |

**(b)**

**Table S15: Statistical analysis comparing model-predicted probabilities against scores on the Clock Drawing Task in the ADNI cohort. (a)** Kruskal-Wallis H tests reveal significant differences in model probabilities for each biomarker across scoring categories of the Clock Drawing Task, suggesting a meaningful alignment between model outputs and a cognitive assessment not used during model training. **(b)** Subsequent Post-Hoc Dunn's tests with FDR correction provide detailed p-values for each pairwise comparison, illustrating the specific differences between scoring categories for $P(A\beta)$ and $P(\tau)$. Refer to Figure 4b for the graphical representation of the data

| | SUVr community 1 | SUVr community 2 | SUVr community 3 |
|---|---|---|---|
| **Model community 1** | 10 | 5 | 18 |
| **Model community 2** | 24 | 7 | 12 |

**(a) Medial temporal,** $p = 0.0228$

| | SUVr community 1 | SUVr community 2 | SUVr community 3 |
|---|---|---|---|
| **Model community 1** | 10 | 15 | 7 |
| **Model community 2** | 3 | 9 | 4 |
| **Model community 3** | 7 | 4 | 17 |

**(b) Lateral temporal,** $p = 0.0048$

| | SUVr community 1 | SUVr community 2 | SUVr community 3 |
|---|---|---|---|
| **Model community 1** | 8 | 13 | 14 |
| **Model community 2** | 5 | 4 | 8 |
| **Model community 3** | 13 | 0 | 11 |

**(c) Medial parietal,** $p = 0.0488$

| | SUVr community 1 | SUVr community 2 | SUVr community 3 |
|---|---|---|---|
| **Model community 1** | 8 | 11 | 5 |
| **Model community 2** | 18 | 1 | 2 |
| **Model community 3** | 0 | 0 | 9 |
| **Model community 4** | 9 | 8 | 5 |

**(d) Lateral parietal,** $p < 0.0001$

| | SUVr community 1 | SUVr community 2 | SUVr community 3 |
|---|---|---|---|
| **Model community 1** | 17 | 13 | 7 |
| **Model community 2** | 3 | 1 | 10 |
| **Model community 3** | 14 | 6 | 5 |

**(e) Frontal,** $p = 0.0226$

| | SUVr community 1 | SUVr community 2 | SUVr community 3 |
|---|---|---|---|
| **Model community 1** | 8 | 1 | 2 |
| **Model community 2** | 2 | 1 | 6 |
| **Model community 3** | 12 | 8 | 36 |

**(f) Occipital,** $p = 0.0278$

**Table S16: Breakdown of model-based and SUVr-derived communities.** The contingent tables show the breakdown of model-based and SUVr-derived communities for each regional label in spatial analysis. Each cell denotes the number of brain regions in the intersection of a model-based community and an SUVr-derived community. $p$ values indicate statistical results from permutation $t$-tests on the adjusted Rand index.

| Group | Median | IQR |
|---|---|---|
| $P(A\beta)$ vs **Cerebral amyloid angiopathy** | | |
| None | 0.37 | [0.32, 0.50] |
| Mild | 0.63 | [0.58, 0.66] |
| Moderate | 0.64 | [0.53, 0.69] |
| Severe | 0.67 | [0.61, 0.68] |
| $P(\tau)$ vs **Cerebral amyloid angiopathy** | | |
| None | 0.41 | [0.33, 0.59] |
| Mild | 0.70 | [0.63, 0.76] |
| Moderate | 0.72 | [0.55, 0.77] |
| Severe | 0.76 | [0.72, 0.79] |
| $P(A\beta)$ vs **Thal phase for amyloid plaques** | | |
| P0 (A0) | 0.35 | [0.30, 0.46] |
| P1 (A1) | 0.35 | [0.28, 0.37] |
| P2 (A1) | 0.34 | [0.30, 0.51] |
| P3 (A2) | 0.59 | [0.37, 0.67] |
| P4 (A3) | 0.65 | [0.59, 0.69] |
| P5 (A3) | 0.63 | [0.57, 0.68] |
| $P(\tau)$ vs **Thal phase for amyloid plaques** | | |
| P0 (A0) | 0.37 | [0.29, 0.49] |
| P1 (A1) | 0.37 | [0.32, 0.39] |
| P2 (A1) | 0.38 | [0.32, 0.61] |
| P3 (A2) | 0.69 | [0.44, 0.74] |
| P4 (A3) | 0.74 | [0.63, 0.78] |
| P5 (A3) | 0.72 | [0.63, 0.77] |
| $P(A\beta)$ vs **Braak stage for neurofibrillary degeneration (NFD)** | | |
| S0 (B0) | 0.34 | [0.29, 0.46] |
| S1 (B1) | 0.36 | [0.28, 0.47] |
| S2 (B1) | 0.34 | [0.32, 0.46] |
| S3 (B2) | 0.61 | [0.58, 0.64] |
| S4 (B2) | 0.40 | [0.31, 0.57] |
| S5 (B3) | 0.63 | [0.59, 0.67] |
| S6 (B3) | 0.65 | [0.59, 0.69] |
| $P(\tau)$ vs **Braak stage for neurofibrillary degeneration (NFD)** | | |
| S0 (B0) | 0.39 | [0.25, 0.57] |
| S1 (B1) | 0.38 | [0.31, 0.46] |
| S2 (B1) | 0.35 | [0.31, 0.49] |
| S3 (B2) | 0.68 | [0.68, 0.69] |
| S4 (B2) | 0.46 | [0.31, 0.57] |
| S5 (B3) | 0.72 | [0.64, 0.74] |
| S6 (B3) | 0.74 | [0.66, 0.79] |
| $P(A\beta)$ vs **CERAD score for density of neocortical neuritic plaque** | | |
| C0 | 0.33 | [0.29, 0.48] |
| C1 | 0.37 | [0.35, 0.60] |

| | | |
|---|---|---|
| C2 | 0.57 | [0.39, 0.65] |
| C3 | 0.64 | [0.59, 0.69] |
| $P(\tau)$ vs **CERAD score for density of neocortical neuritic plaque** | | |
| C0 | 0.35 | [0.29, 0.52] |
| C1 | 0.44 | [0.36, 0.64] |
| C2 | 0.61 | [0.46, 0.73] |
| C3 | 0.73 | [0.66, 0.78] |
| $P(A\beta)$ vs **CERAD score for density of diffuse plaques** | | |
| No diffuse plaques | 0.35 | [0.30, 0.46] |
| Sparse diffuse plaques | 0.37 | [0.28, 0.42] |
| Moderate diffuse plaques | 0.58 | [0.37, 0.63] |
| Frequent diffuse plaques | 0.64 | [0.57, 0.69] |
| $P(\tau)$ vs **CERAD score for density of diffuse plaques** | | |
| No diffuse plaques | 0.39 | [0.30, 0.50] |
| Sparse diffuse plaques | 0.37 | [0.32, 0.50] |
| Moderate diffuse plaques | 0.68 | [0.41, 0.75] |
| Frequent diffuse plaques | 0.72 | [0.63, 0.77] |

**Table S17: Median & interquartile range (IQR) for model probabilities (P(A$\beta$) & P($\tau$)) against various neuropathological grades.** Refer to Figure 6 for the graphical representation of the data and to Table S18 for detailed test statistics and associated p-values.

| Neuropathology Variable | Model Probability | Test | Statistic | P-value |
|---|---|---|---|---|
| Cerebral amyloid angiopathy | $P(A\beta)$ | Spearman correlation | $\rho = 0.43$ | $1.69 \times 10^{-7}$ |
| | | Kruskal-Wallis | $H = 31.74$ | $5.94 \times 10^{-7}$ |
| | $P(\tau)$ | Spearman correlation | $\rho = 0.45$ | $7.76 \times 10^{-8}$ |
| | | Kruskal-Wallis | $H = 33.30$ | $2.78 \times 10^{-7}$ |
| Thal phase for amyloid plaques | $P(A\beta)$ | Spearman correlation | $\rho = 0.50$ | $1.00 \times 10^{-9}$ |
| | | Kruskal-Wallis | $H = 46.85$ | $6.09 \times 10^{-9}$ |
| | $P(\tau)$ | Spearman correlation | $\rho = 0.47$ | $1.71 \times 10^{-8}$ |
| | | Kruskal-Wallis | $H = 42.80$ | $4.05 \times 10^{-8}$ |
| Braak stage for neurofibrillary degeneration (NFD) | $P(A\beta)$ | Spearman correlation | $\rho = 0.54$ | $2.95 \times 10^{-11}$ |
| | | Kruskal-Wallis | $H = 46.74$ | $2.11 \times 10^{-8}$ |
| | $P(\tau)$ | Spearman correlation | $\rho = 0.59$ | $2.31 \times 10^{-13}$ |
| | | Kruskal-Wallis | $H = 50.25$ | $4.19 \times 10^{-9}$ |
| CERAD score for density of neocortical neuritic plaque | $P(A\beta)$ | Spearman correlation | $\rho = 0.60$ | $2.04 \times 10^{-14}$ |
| | | Kruskal-Wallis | $H = 49.87$ | $8.50 \times 10^{-11}$ |
| | $P(\tau)$ | Spearman correlation | $\rho = 0.63$ | $7.87 \times 10^{-16}$ |
| | | Kruskal-Wallis | $H = 53.27$ | $1.61 \times 10^{-11}$ |
| CERAD score for density of diffuse plaques | $P(A\beta)$ | Spearman correlation | $\rho = 0.56$ | $5.34 \times 10^{-12}$ |
| | | Kruskal-Wallis | $H = 41.10$ | $6.22 \times 10^{-9}$ |
| | $P(\tau)$ | Spearman correlation | $\rho = 0.50$ | $1.80 \times 10^{-9}$ |
| | | Kruskal-Wallis | $H = 33.98$ | $2.00 \times 10^{-7}$ |

**Table S18: Statistical results for neuropathological validation of model probabilities, as shown in Figure 6 and described in detail in Table S17.** For each neuropathology variable and model probability label ($P(A\beta)$ & $P(\tau)$), both the Spearman correlation coefficient $\rho$ and the Kruskal Wallis $H$ statistic are presented along with their associated p-values.
